# Explaining variation in antibiotic prescribing for common infections: a three-way variance decomposition using UK primary care data

**DOI:** 10.64898/2026.05.04.26352315

**Authors:** Nam Vinh Nguyen, Julie V. Robotham, A. Sarah Walker, David W. Eyre, Russell Hope, Christopher C. Butler, Mike Sharland, Koen B. Pouwels

**Affiliations:** Nuffield Department of Primary Care Health Sciences, University of Oxford, UK; NIHR Health Protection Research Unit in Healthcare Associated Infections and AMR at University of Oxford in partnership with the UK Health Security Agency; Antibiotic Policy Group, City St George’s University London, London, UK; AMR Modelling and Evaluation, UK Health Security Agency, London, UK; Modernising Medical Microbiology Unit, Nuffield Department of Medicine, University of Oxford, UK; Antimicrobial Resistance and Healthcare Associated Infections Division, UK Health Security Agency, London, UK

## Abstract

**Background:** Benchmarking has been used to target clinically unwarranted variation in antimicrobial prescribing in UK primary care. However, variation in antibiotic prescribing between general practices may be partly explained by differences in case-mix. We aimed to quantify how much variation in antibiotic prescribing for common infections was attributable to differences in prescribing between practices after accounting for case mix.

**Methods:** We used the UK Clinical Practice Research Datalink (CPRD) Aurum database to identify GP consultations for 11 common infections. Three-way variance decomposition quantified the proportions of total variance attributable to patient case-mix, between practice variation, and residual unexplained variance using variables recorded in electronic medical records, across three models (no, minimal (age/sex), and full case mix adjustment). For lower respiratory tract infection (LRTI) and sore throat, external data to impute illness severity were used to estimate the potential effect of unmeasured infection severity.

**Findings:** We identified 3,820,806 consultations in 2019. There was clear variability in antibiotic prescribing across practices for most conditions. In fully adjusted models, between practice variation explained 5.8–32.6% of total variance, exceeding variation attributed to case-mix in 9 out of 11 infections. Compared to no adjustment, full case-mix adjustment reduced between-practice differences, lowering their contribution to total explained variation by more than 20% in 6 of 11 infections and by 10–20% in 4 others. Minimal age–sex adjustment had little impact, with changes below 5% in 8 of 11 infections. Imputing infection severity in addition to full case-mix adjustment further reduced contribution of between-practice variance to the total variance (by 25.9% for LRTI and 8.5% for sore throat).

**Interpretation:** Differences in practice-level prescribing, beyond patient case-mix, call for targeted interventions and highlight the value of providing feedback at the practice level. Comprehensive case-mix adjustment, including imputed infection severity, improves the assessment of prescribing variation and supports fairer performance comparisons.

**Funding:** Wellcome Trust; NIHR

**RESEARCH IN CONTEXT:** *Evidence before this study:* We searched MEDLINE for articles published between 1 January 2005 and 31 July 2025, using a combination of key terms including “antibiotic” (or “antimicrobial” or “antibacterial”), “prescribing” (or “prescription” or “use” or “utilisation” or “utilization”), “primary health care” (or “primary care” or “general practice” or “general practitioner” or “GP”), and “United Kingdom” (or “UK” or “England”). We focused on studies using patient-level data to compare antibiotic prescribing between general practices (GP practices). Most studies assessed overall prescribing or focused on a small subset of infections. Only a few examined condition-specific measures across a broader range of infections. We found no studies that decomposed variance into that caused by patient case-mix versus practice performance adjusting for case-mix across a wide range of infections.

*Added value of this study:* Using individual-level data from 3.8 million consultations for eleven common infections in the UK Clinical Practice Research Datalink (CPRD) Aurum, we applied three-way variance decomposition to quantify the proportions of total variance attributable to patient case-mix, between-practice differences after adjusting for case-mix, and residual variation under three adjustment strategies (non, age-sex only, and full case-mix adjustment). There was clear variability in antibiotic prescribing across practices for most conditions. The total variance attributable to between-practice differences exceeded that attributed to case-mix in 9 out of 11 infections according, according to the fully adjusted case-mix models. Fully adjusting for case-mix based on routinely collected data substantially reduced between-practice differences, lowering their contribution to explained variance (the sum of the patient case-mix variance and between-practice variance) by more than 20% in 6 of 11 infections and by 10–20% in 4 others, whereas minimal age–sex adjustment had little impact. Between-practice differences were reduced further incorporating external information to simulate unmeasured infection severity.

*Implications of all the available evidence:* Differences in practice-level prescribing, beyond patient case-mix, call for targeted interventions and highlight the value of providing feedback at the practice level. Full case-mix adjustment substantially reduces the risk of overstating between-practice differences, performing far better than adjusting for age/sex alone. Condition-specific indicators with sufficient case-mix adjustment may be more effective benchmarks of practice performance than aggregated total antibiotic use levels as general practitioners (GPs) are more likely to respond positively to comparisons they perceive as fair. In particular, acute otitis media and upper respiratory tract infection, conditions with substantial variability in antibiotic prescribing across GP practices and the highest variance attributable to adjusted between-practice differences (12.6% and 10.3%, respectively), are promising candidates for fair prescribing indicators.

## INTRODUCTION

Antimicrobial resistance (AMR) is a major challenge which affects all health systems.^1^ Because antibiotic use drives AMR, promoting responsible use of current antibiotics has become a central goal of antibiotic stewardship strategies.^2^ In the UK, antibiotic use declined steadily between 2014 and 2020, with a sharp drop in 2020 during the COVID-19 pandemic. However, recent data indicate a reversal of this trend, with antibiotic use increasing in 2022, and increases continuing into 2023 across all primary and secondary care settings, except dentistry. Prescribing levels are now comparable to those observed in 2019, accompanied by a rise in resistant cases and deaths from severe infections.^3,4^

Approximately 70% of antibiotics in the UK are prescribed in primary care, and at least 20% of these prescriptions are estimated to be inappropriate.^5–7^ Reducing such overuse requires a better understanding why antibiotic prescribing varies between general practitioner practices (GP practices). Differences may reflect patient characteristics, infection severity, or prescriber’s threshold for prescribing given diagnostic uncertainty.^8^

Benchmarking interventions, which provide GP practices with feedback comparing their antibiotic prescribing rates with peers, have been moderately effective at reducing prescribing.^9,10^ However, a persistent challenge is ensuring that such comparisons are fair. Practices differ in their case-mix (e.g. age, sex, comorbidity, and socioeconomic deprivation) and existing UK monitoring metrics, such as the Specific Therapeutic group Age-sex Related Prescribing Unit (STAR-PU) adjustment, account only for age and sex. Consequently, observed differences in prescribing rates may reflect variations in patient populations rather than true differences in prescriber behaviour.

Previous work using aggregated practice-level data concluded that age-sex weighted comparisons are unfair for some practices, with 21% of practices being no longer classed as high prescribers once comorbidity, smoking prevalence, and deprivation were accounted for.^11^ However, the use of aggregated data meant that individual-level patient characteristics could not be included in adjustments, and patient-level and practice-level effects could not be disentangled. Consequently, associations observed at the practice level may not reflect relationships at the individual level, making the study susceptible to the ecological fallacy. For example, a higher prevalence of diabetes at the practice level may be associated with higher overall antibiotic prescribing, even if the excess prescribing arises from consultations among healthy adults without diabetes.

Analyses using individual patient-level data allow for more robust adjustment for case-mix, direct estimation of patient and practice contributions to prescribing variation, and evaluation of condition-specific patterns that may be more salient for action for GP practices. Robust adjustment for case-mix is essential for fair comparisons but also for likely effectiveness of benchmarking, as general practitioners (GPs) are more likely to respond positively to comparisons they perceive as fair and reflective of differences in case-mix.^12^

To address these issues, we used individual-level data from the UK Clinical Practice Datalink (CPRD) Aurum and applied three-way variance decomposition models ^13^ to quantify how much of the observed variation in antibiotic prescribing for 11 common infections reflected patient case-mix versus practice performance. By comparing three levels of adjustment, none, age-sex only, and full case-mix adjustment, and conducting sensitivity analyses using a previous study that measured infection severity,^14^ we aimed to identify which models provide the fairest and most accurate basis for interpreting variation in antibiotic prescribing across primary care in England. In addition, by examining the extent of between-practice variation in prescribing across multiple infection types after case-mix adjustment, we aimed to identify conditions that may serve as robust indicators of antibiotic prescribing quality. Rather than relying solely on expert consensus, which have generated over 700 proposed indicators of antibiotic prescribing quality,^15^ our approach uses observed prescribing variation and case-mix adjustment to provide an evidence-based framework for selecting indicators that are more likely to capture true between-practice differences in prescribing propensity after accounting for patient case-mix.

## METHODS

### Study design and participants

We conducted a cross-sectional study using the CPRD Aurum database linked to Hospital Episode Statistics (HES) Admitted Patient Care and Office for National Statistics (ONS) death registration data. CPRD Aurum is a large UK primary care database that is broadly representative of the UK population.^16^ The database contains information on patients’ GP practice, demographic characteristics, diagnoses and symptoms, prescriptions, vaccination history, laboratory tests, and specialist referrals.

We identified GP consultations between 1 January and 31 December 2019 for new episodes of 11 common infections: acute cough, acute sore throat, acute rhinosinusitis, acute bronchitis, any upper respiratory tract infection (URTI), any lower respiratory tract infection (LRTI), acute otitis media (in patients under 18 years), asthma exacerbation, chronic obstructive pulmonary disease (COPD) exacerbation (in patients above 40 years), acute lower urinary tract infection (lower UTI), or impetigo *(Appendix 1)*. An infection episode was considered new if, in the preceding 30 days, the patient had not received any antibiotic prescription and had not consulted a GP for the same or a related diagnosis of infection within the same body system (e.g. acute sinusitis and URTI or acute bronchitis and LRTI). A new episode also required no hospital admission, emergency department attendance, or outpatient visit for the same or a related infection diagnosis either within the preceding 30 days or on the index consultation date, identified using corresponding ICD-10 codes in Hospital Episode Statistics (HES) *(Appendix 2)*. These were excluded because they may represent cases where a GP refers the patient to hospital rather than prescribing an antibiotic. Consultations involving pregnant women were excluded due to distinct prescribing patterns during pregnancy and too few infection-related consultations for a pregnancy-specific analysis.

To minimise bias due to including practices that often do not use informative Read codes–the clinical coding system used in UK primary care to record diagnoses and symptoms^17^–we applied infection-specific thresholds to exclude practices.^5^ For infections with >500,000 consultations in 2019 (URTI, acute cough, LRTI and lower UTI), practices reporting <100 consultations in 2019 were excluded; for those with fewer consultations (acute rhinosinusitis, acute sore throat, acute otitis media, acute bronchitis, asthma exacerbation, and COPD exacerbation), practices reporting <50 consultations in 2019 were excluded.

### Statistical analysis

We focused on systemic antibiotics *(Appendix 3)* prescribed on the same day as the infection diagnosis. We first calculated and, summarised across practices, the percentage of antibiotic prescriptions for each infection, overall and by Access, Watch, and Reserve (AWaRe) categories following the World Health Organization (WHO) and UK classification systems.^18,19^

To assess the extent to which variation in antibiotic prescribing could be causally attributed to different sources, we applied the three-way variance decomposition method of Chen et al.^13^ This method decomposes the total variance in prescribing into: (i) variance explained by patient case-mix, (ii) variance explained by between-practice differences after adjusting for case-mix, and (iii) residual (unexplained) variance (details in *Appendix 4*).

The case-mix variance captures the portion of total variance attributable to the measured patient characteristics, including sociodemographic factors (age, sex, ethnicity, and deprivation status^18^), smoking status, comorbidities (Elixhauser comorbidity score^19^, diabetes, chronic kidney disease, and chronic lung disease), previous use of immunosuppressants, history of hospitalisation and/or outpatient visits in the preceding year, and vaccination status (influenza and pneumococcal vaccines). Covariates were selected for each condition based on clinical relevance. For example, chronic lung disease, and influenza and pneumococcal vaccination, were included only in models for respiratory infections and not for UTIs or impetigo. Associations between these covariates and the antibiotic prescribing decision are shown in *Appendix 5*.

After adjusting for measured case-mix, the between-practice variance captures systematic differences across practices, such as practice-level prescribing norms or local policy. The residual variance, at the patient level, is primarily attributable to unmeasured patient characteristics, differences between prescribers within an individual practice, both across and within professional groups, and random noise. However, it may also include practice-level influences that vary across patient subgroups that cannot be modelled as interactions due to large numbers of practices.

Decomposition analyses were conducted separately for each infection type, using a 20% random sample of included GP practices to ensure computational feasibility. For each model, we reported the proportion of variance explained by each component, along with 95% confidence intervals using 100 posterior simulations based on the estimated coefficients and variance-covariance matrices.^22^

To evaluate whether a more limited set of case-mix adjustments may be sufficient, we fitted two alternative models in addition to the fully adjusted main model (Model 1). Model 2 included only patient-level age and sex as covariates akin the current STAR-PU methodology, and Model 3 included only practice identifiers without adjustment for case-mix.

Because infection severity is not captured in routine electronic health record databases such as CPRD, we performed a sensitivity analysis using simulation-based imputation. Severity distributions for adults (≥16 years) with sore throat or LRTI (FeverPAIN score for sore throat and Pneumonia risk score for LRTI) were obtained from Stuart et al.^14^ That study reported the severity distribution by (i) GP practice antibiotic-prescribing proportion (in quartiles) and (ii) prescribing decision (antibiotics prescribed vs. not prescribed) *(Appendix 6)*.

To estimate the joint probability distribution of infection severity conditional on both GP practice prescribing quartile and prescribing decision, we combined these two marginal distributions using iterative proportional fitting (IPF). This procedure generates a three-way contingency table matching the observed margins from Stuart et al.^14^ while preserving the total number of consultations for adults (≥16 years) in our CPRD practices. For each practice, the numbers of consultations with and without antibiotics were then allocated across severity levels according to this fitted joint distribution.

To account for uncertainty in the estimated proportions, we simulated 100 draws from a Dirichlet distribution parameterised by the fitted cell counts within each practice and prescribing group. The three-way variance decomposition was subsequently performed in every replicate and variance components were summarised across replicates.

## RESULTS

From the CPRD Aurum database, 3,820,806 records of GP consultations for new episodes of common infections in 2019 were identified. Acute cough (968,611), URTI (840,955), UTI (677,540), and LRTI (531,470) accounted for most consultations. Median ages varied by infection, ranging between 20 and 60 years, and females outnumbered males for all infections, except for otitis media. Patients from the two most deprived quintiles accounted for the largest share of consultations. Comorbidities and recent antibiotics use (within six months) were most common among those with lower respiratory tract-related conditions (COPD or asthma exacerbation, LRTI, and bronchitis) and UTI (Table 1).

**Table 1.** Patient characteristics for GP consultations for new episodes of one of 11 common infections in 2019 (n = 3,820,806)

|  | URTI | Acute cough | Acute rhinosinusitis | Acute sore throat | LRTI | Acute bronchitis | Acute otitis media<br>[age <18 years] | Asthma exacerbation | COPD exacerbation<br>[age ≥40 years] | Lower UTI | Impetigo |
| --- | --- | --- | --- | --- | --- | --- | --- | --- | --- | --- | --- |
| Number of visits, n (%) | 840,955 | 968,611 | 201,577 | 432,490 | 531,470 | 11,944 | 98,484 | 33,042 | 17,969 | 677,540 | 16,724 |
| Age (median [IQR]) | 12 [2-40] | 45 [13-66] | 42 [28-58] | 24 [12-40] | 54 [29-72] | 55 [36-70] | 3 [1-6] | 46 [28-60] | 71 [63-78] | 52 [29-73] | 9 [4-26] |
| Age group: ≤ 5 | 337,062 (40.1) | 179,343 (18.5) | 10,999 (5.5) | 54,839 (12.7) | 68,349 (12.9) | 613 (5.1) | 70,503 (71.6) | 776 (2.3) | 0 (0) | 22,793 (3.4) | 5,802 (34.7) |
| Age group: 6-9 | 56,656 (6.7) | 42,287 (4.4) | 3,802 (1.9) | 34,771 (8) | 11,460 (2.2) | 154 (1.3) | 15,645 (15.9) | 1,349 (4.1) | 0 (0) | 15,590 (2.3) | 2,598 (15.5) |
| Age group: 10-17 | 67,749 (8.1) | 47,853 (4.9) | 9,163 (4.5) | 62,926 (14.5) | 15,108 (2.8) | 316 (2.6) | 12,336 (12.5) | 2,452 (7.4) | 0 (0) | 22,691 (3.3) | 2,324 (13.9) |
| Age group: 18-24 | 52,997 (6.3) | 49,197 (5.1) | 15,871 (7.9) | 68,291 (15.8) | 21,802 (4.1) | 585 (4.9) | 0 (0) | 2,478 (7.5) | 0 (0) | 68,694 (10.1) | 1,615 (9.7) |
| Age group: 25-39 | 110,903 (13.2) | 118,026 (12.2) | 52,723 (26.2) | 100,777 (23.3) | 64,720 (12.2) | 1,799 (15.1) | 0 (0) | 6,356 (19.2) | 0 (0) | 123,245 (18.2) | 1,991 (11.9) |
| Age group: 40-59 | 116,693 (13.9) | 207,961 (21.5) | 64,071 (31.8) | 70,281 (16.3) | 122,430 (23) | 3,545 (29.7) | 0 (0) | 11,038 (33.4) | 3,215 (17.9) | 144,645 (21.3) | 1,482 (8.9) |
| Age group: 60-74 | 62,722 (7.5) | 184,094 (19) | 33,119 (16.4) | 28,352 (6.6) | 110,669 (20.8) | 2,967 (24.8) | 0 (0) | 5,871 (17.8) | 8,146 (45.3) | 122,712 (18.1) | 557 (3.3) |
| Age group: ≥ 75 | 36,173 (4.3) | 139,850 (14.4) | 11,829 (5.9) | 12,253 (2.8) | 116,932 (22) | 1,965 (16.5) | 0 (0) | 2,722 (8.2) | 6,608 (36.8) | 157,170 (23.2) | 355 (2.1) |
| Female gender, n (%) | 462,112 (55) | 525,739 (54.3) | 126,366 (62.7) | 266,590 (61.6) | 295,830 (55.7) | 6,921 (57.9) | 47,156 (47.9) | 20,782 (62.9) | 9,844 (54.8) | 577,065 (85.2) | 9,216 (55.1) |
| Ethnicity, n (%) |  |  |  |  |  |  |  |  |  |  |  |
| Asian | 133,199 (15.9) | 95,357 (9.9) | 19,995 (9.9) | 52,321 (12.1) | 42,475 (8) | 833 (7) | 9,962 (10.1) | 3,050 (9.2) | 148 (0.8) | 42,962 (6.3) | 634 (3.8) |
| Black | 42,620 (5.1) | 34,306 (3.5) | 6,585 (3.3) | 17,052 (3.9) | 14,708 (2.8) | 210 (1.8) | 2,258 (2.3) | 1,152 (3.5) | 56 (0.3) | 16,376 (2.4) | 146 (0.9) |
| Mix or multiple | 30,310 (3.6) | 21,031 (2.2) | 3,661 (1.8) | 11,571 (2.7) | 8,403 (1.6) | 146 (1.2) | 3,962 (4) | 610 (1.8) | 42 (0.2) | 9,024 (1.3) | 398 (2.4) |
| White | 594,194 (70.7) | 780,168 (80.6) | 164,171 (81.5) | 331,516 (76.7) | 447,325 (84.2) | 10,375 (87) | 76,845 (78.1) | 27,435 (83.1) | 17,330 (96.5) | 587,329 (86.8) | 14,594 (87.3) |
| Other | 7,700 (0.9) | 4,434 (0.5) | 1,191 (0.6) | 2,859 (0.7) | 1,873 (0.4) | 39 (0.3) | 699 (0.7) | 77 (0.2) | 6 (0) | 2,226 (0.3) | 51 (0.3) |
| Unknown | 32,294 (3.8) | 32,578 (3.4) | 5,776 (2.9) | 16,738 (3.9) | 16,300 (3.1) | 326 (2.7) | 4,692 (4.8) | 692 (2.1) | 371 (2.1) | 18,993 (2.8) | 885 (5.3) |
| IMD quintile, n (%) |  |  |  |  |  |  |  |  |  |  |  |
| 1 (least deprived) | 108,991 (17.3) | 143,083 (19.2) | 32,869 (21.2) | 60,528 (18.6) | 72,555 (18.4) | 1,827 (21) | 16,837 (22.2) | 4,198 (16.5) | 1,234 (9.2) | 110,638 (21.5) | 3,169 (25.4) |
| 2 | 110,217 (17.4) | 147,570 (19.8) | 32,618 (21.1) | 61,461 (18.9) | 75,730 (19.2) | 1,844 (21.2) | 15,414 (20.4) | 4,854 (19.1) | 2,087 (15.5) | 108,886 (21.1) | 2,750 (22.1) |
| 3 | 85,382 (13.5) | 105,607 (14.2) | 22,404 (14.5) | 44,942 (13.8) | 52,010 (13.2) | 1,164 (13.4) | 10,189 (13.5) | 3,362 (13.2) | 1,516 (11.2) | 74,323 (14.4) | 1,617 (13) |
| 4 | 171,793 (27.2) | 189,298 (25.4) | 38,406 (24.8) | 84,712 (26.1) | 98,750 (25) | 2,092 (24.1) | 17,487 (23.1) | 6,588 (25.9) | 3,557 (26.4) | 125,542 (24.4) | 2,510 (20.2) |
| 5 (most deprived) | 155,368 (24.6) | 159,494 (21.4) | 28,641 (18.5) | 73,521 (22.6) | 95,405 (24.2) | 1,755 (20.2) | 15,748 (20.8) | 6,436 (25.3) | 5,085 (37.7) | 95,469 (18.5) | 2,409 (19.3) |
| Ever smoked, n (%) | 223,627 (26.6) | 431,417 (44.5) | 88,803 (44.1) | 152,688 (35.3) | 249,334 (46.9) | 6,209 (52) | 5,160 (5.2) | 22,439 (67.9) | 12,411 (69.1) | 302,378 (44.6) | 3,417 (20.4) |
| Elixhauser score, n (%) |  |  |  |  |  |  |  |  |  |  |  |
| ≤ 0 | 745,293 (88.6) | 707,195 (73) | 158,527 (78.6) | 375,998 (86.9) | 350,241 (65.9) | 8,745 (73.2) | 97,095 (98.6) | 24,473 (74.1) | 1,746 (9.7) | 445,093 (65.7) | 15,256 (91.2) |
| 1–5 | 59,784 (7.1) | 151,437 (15.6) | 29,626 (14.7) | 38,778 (9) | 95,729 (18) | 1,972 (16.5) | 683 (0.7) | 5,951 (18) | 7,583 (42.2) | 121,837 (18) | 911 (5.4) |
| 5–9 | 16,944 (2) | 55,494 (5.7) | 5,523 (2.7) | 7,527 (1.7) | 39,627 (7.5) | 567 (4.7) | 582 (0.6) | 1,181 (3.6) | 5,767 (32.1) | 47,170 (7) | 269 (1.6) |
| ≥ 10 | 18,934 (2.3) | 54,485 (5.6) | 7,901 (3.9) | 10,187 (2.4) | 45,873 (8.6) | 660 (5.5) | 124 (0.1) | 1,437 (4.3) | 2,873 (16) | 63,440 (9.4) | 288 (1.7) |
| Diabetes, n (%) | 40,918 (4.9) | 104,912 (10.8) | 15,167 (7.5) | 20,298 (4.7) | 74,178 (14) | 1,604 (13.4) | 292 (0.3) | 3,807 (11.5) | 4,256 (23.7) | 83,794 (12.4) | 341 (2) |
| Chronic renal disease, n (%) | 63,891 (7.6) | 161,044 (16.6) | 31,570 (15.7) | 40,928 (9.5) | 113,555 (21.4) | 1,994 (16.7) | 559 (0.6) | 6,385 (19.3) | 7,372 (41) | 153,734 (22.7) | 1,058 (6.3) |
| Prescribed at least one immunosuppressant within the past 6 months, n (%) | 35,478 (4.2) | 115,251 (11.9) | 11,305 (5.6) | 15,121 (3.5) | 93,538 (17.6) | 2,366 (19.8) | 2,389 (2.4) | 25,673 (77.7) | 14,894 (82.9) | 39,465 (5.8) | 430 (2.6) |
| Received flu vaccine within the past year, n (%) | 129,740 (15.4) | 231,207 (23.9) | 31,066 (15.4) | 61,548 (14.2) | 132,947 (25) | 2,516 (21.1) | 22,431 (22.8) | 11,156 (33.8) | 7,547 (42) | 157,981 (23.3) | 3,820 (22.8) |
| Received pneumococcal vaccines within the past 10 years, n (%) | 404,553 (48.1) | 309,832 (32) | 27,448 (13.6) | 104,587 (24.2) | 136,380 (25.7) | 2,151 (18) | 85,061 (86.4) | 5,787 (17.5) | 5,142 (28.6) | 106,959 (15.8) | 8,722 (52.2) |
| Hospital, emergency department, or outpatient admission for any cause in the past year, n (%) | 366,641 (43.5) | 451,382 (46.6) | 86,447 (42.9) | 164,735 (38.1) | 258,544 (48.6) | 5,243 (100) | 43,390 (43.9) | 16,036 (16.3) | 10,334 (31.3) | 331,874 (49.0) | 5,839 (34.9) |

### Antibiotic prescribing (%) by infection

The percentage of patients receiving an antibiotic differed by infection diagnosis, exceeding 70% for five infections (lower UTI: 81.8%, LRTI: 79.7%, COPD exacerbation: 77.1%, otitis media: 76.3% and bronchitis: 75.4%). For four infections, this percentage ranged between 40% and approximately 50% (rhinosinusitis: 50.7%, asthma exacerbation: 46.7%, impetigo: 41.6% and sore throat: 40.6%). For cough, it was 33.5%: URTI (15.8%) was the only condition with a percentage below 20%. For all infections, Access-group antibiotics accounted for over 85% of prescriptions. Antibiotic choice was aligned with UK guidelines, with narrow-spectrum agents commonly prioritised for the conditions studied (i.e. amoxicillin, phenoxymethylpenicillin, and doxycycline for respiratory infections; nitrofurantoin and trimethoprim for UTIs; and flucloxacillin for skin infections) (Table 2).

**Table 2.**
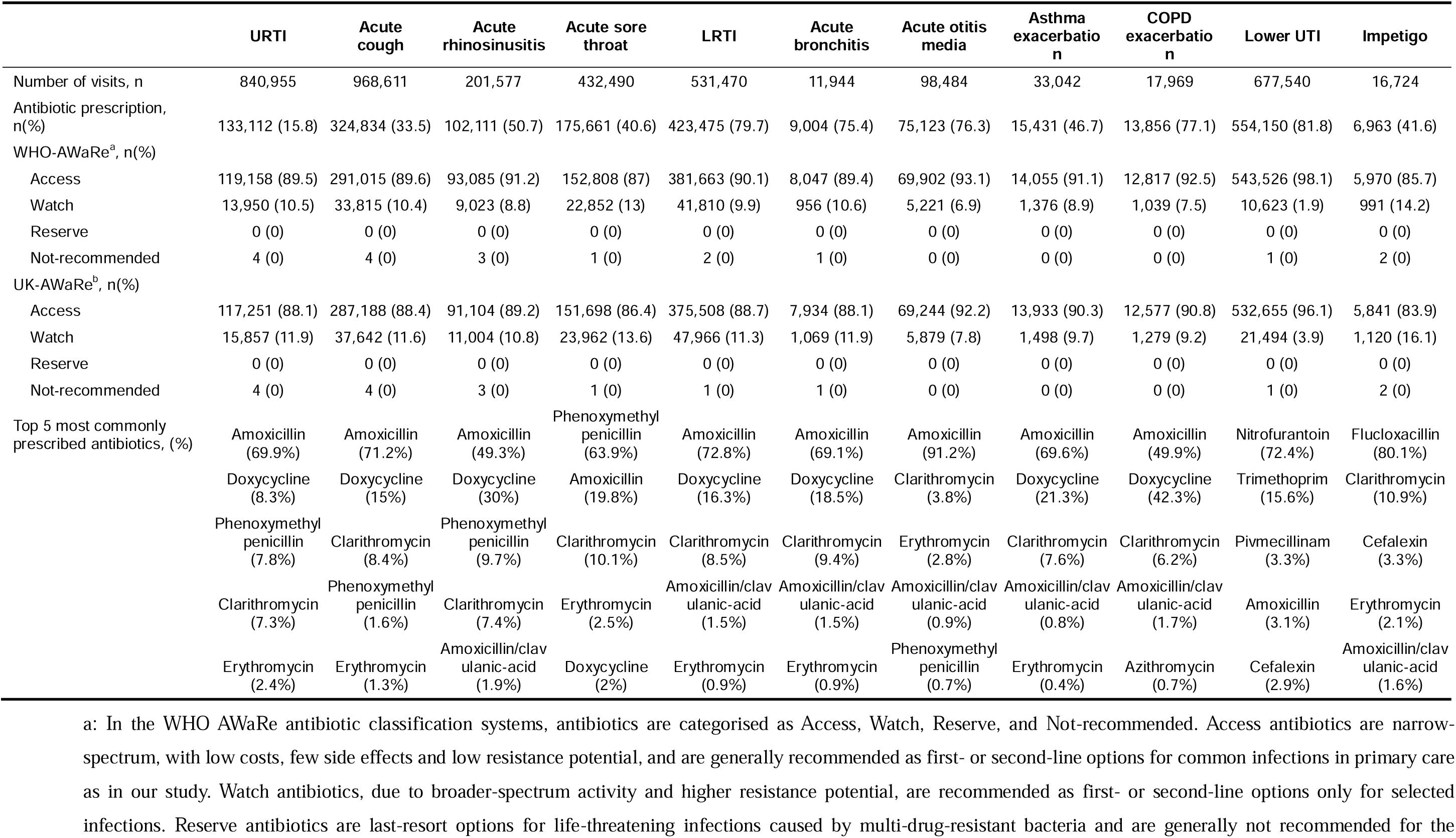
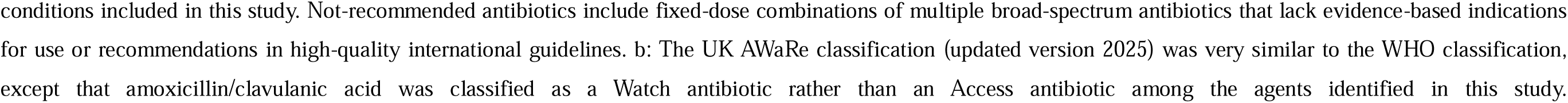
Antibiotic prescribing patterns by infection.

### Variability in practice-level antibiotic prescribing across GP practices

There was clear variability in practice-level antibiotic prescribing percentages across GP practices, with the width of the interquartile range (IQR) above 10 percentage points (pp) in all infections. IQRs exceeded 20 percentage points for asthma exacerbation (21.2pp) and acute bronchitis (20.1pp), and were between 14–20 percentage points for acute rhinosinusitis (19.1 pp), COPD exacerbation (16.1pp), acute sore throat (15.2pp), acute cough (14.6pp), acute otitis media (14.5pp) and impetigo (14.5pp). For acute cough, although its IQR was lower (10.6pp), given its relatively low median antibiotic prescribing percentages (15.8%), the between-practice variability was still substantial. Lower UTIs and LRTIs showed the lowest variability, with IQRs of 10.6pp and 12.2pp, respectively, but also had high median antibiotic prescribing percentages (81.7% and 79.5%) (Figure 1).

**Figure 1.**
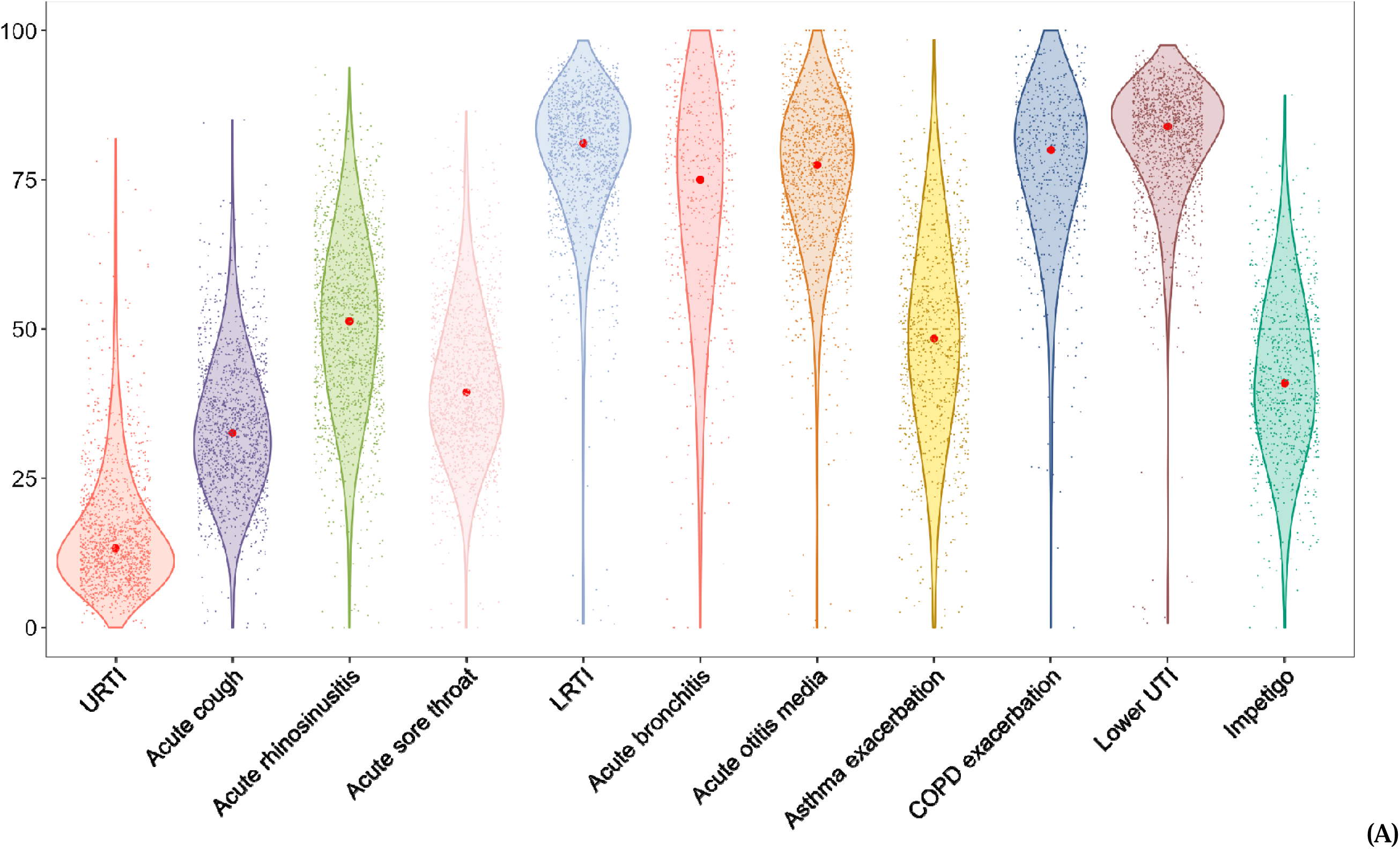
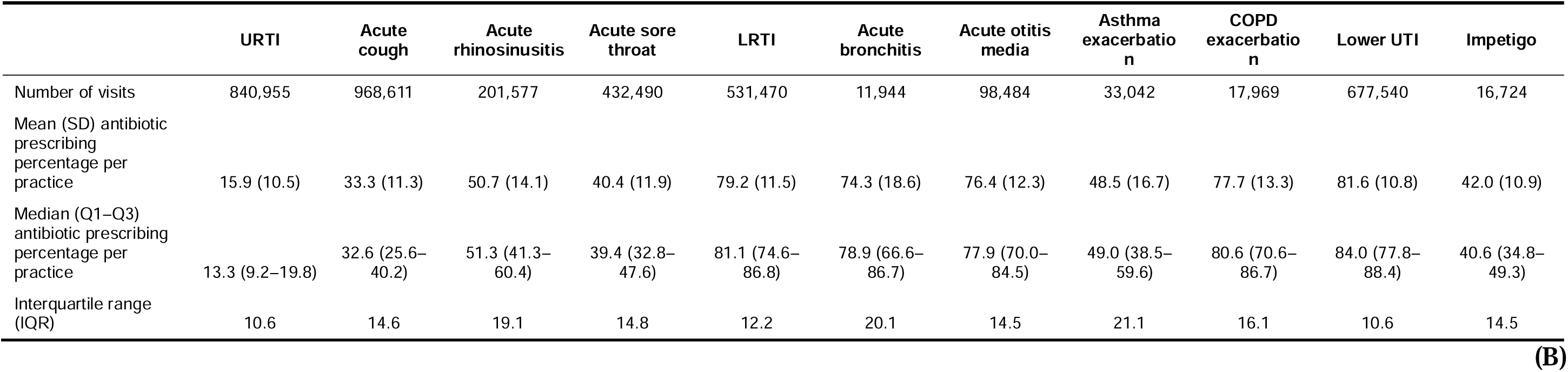
Variability in antibiotic prescribing across GP practices by infection. Only practices with at least 100 consultations in 2019 were included for infections with more than 500,000 consultations (URTI, acute cough, LRTI, lower UTI), and those with at least 50 consultations for less frequent infections (acute rhinosinusitis, acute sore throat, bronchitis, asthma exacerbation, COPD exacerbation, and impetigo). (A) Violin chart illustrating the variability in antibiotic prescribing proportions among GP practices across different infections. The x-axis shows infections, and the y-axis shows the proportion of antibiotic prescriptions. Each dot corresponds to an individual GP’s prescribing proportion. The vertical span of each violin represents the distribution of prescribing proportions, with the thickest (widest) parts indicating where values are most concentrated among GPs. The lines extend from the minimum to the maximum observed proportions. The red dots mark the median prescribing proportion for each infection type. (B) Summary table reporting measures of variability for each infection, including the mean and standard deviation (SD), median, first and third quartiles (Q1, Q3), and interquartile range (IQR).

### Variance decomposition analyses

Across infections, between-practice differences explained 5.8-32.6% of total variance in antibiotic prescribing in the fully adjusted models, whereas patient case-mix accounted for 0.8-11.4%. In 9 of the 11 infections, the between-practice component of the variance, after adjusting for patient case-mix, was larger than the case-mix component (Table 3). After case-mix adjustment, the conditions with the highest proportion of total variance attributable to between-practice variation, a pre-requisite for a useful prescribing quality indicator, were acute bronchitis (32.6%), acute otitis media (12.6%), LRTI (10.5%), URTI (10.3%) and UTI (10.0%) (Table 3).

**Table 3.**
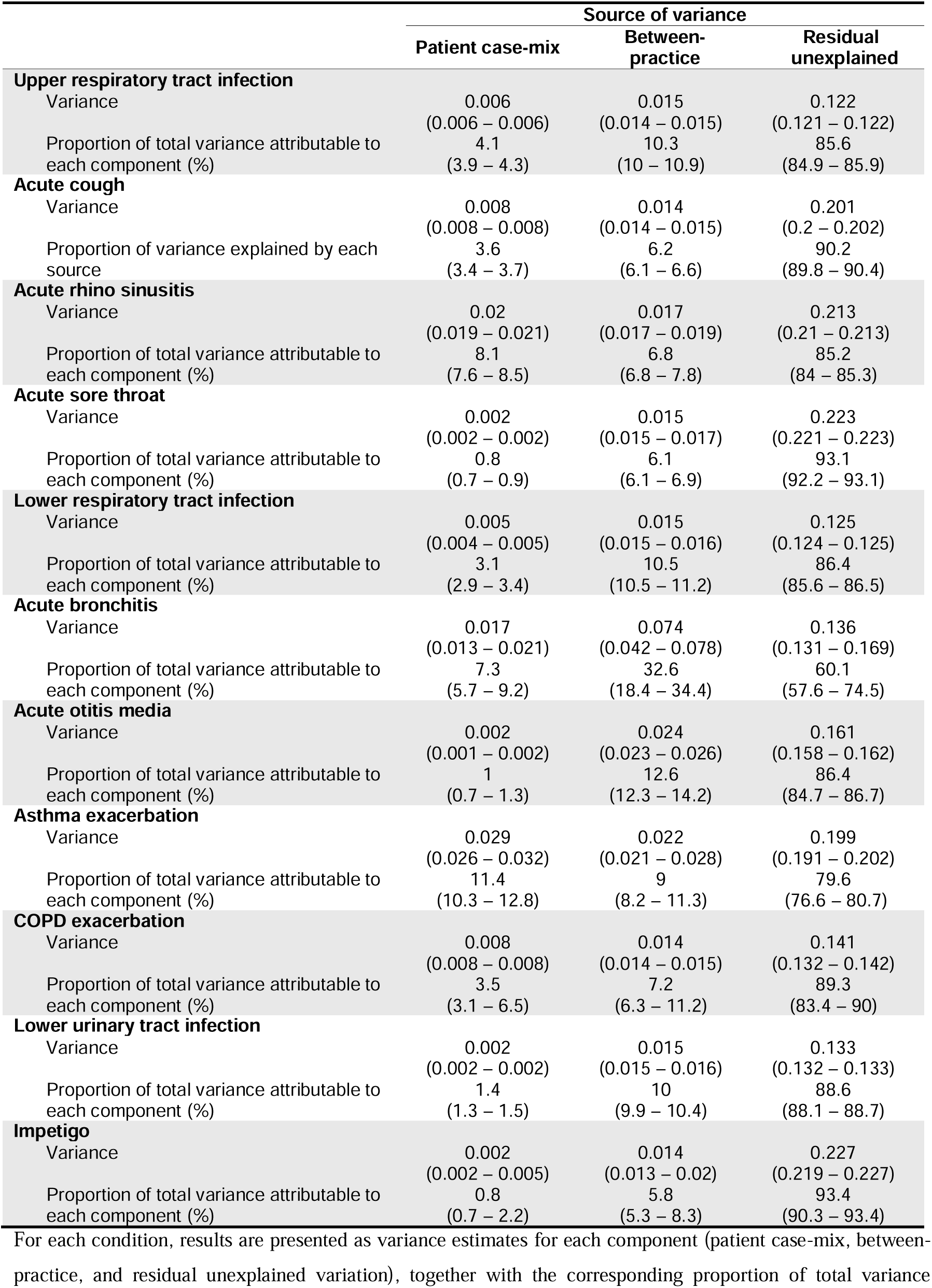
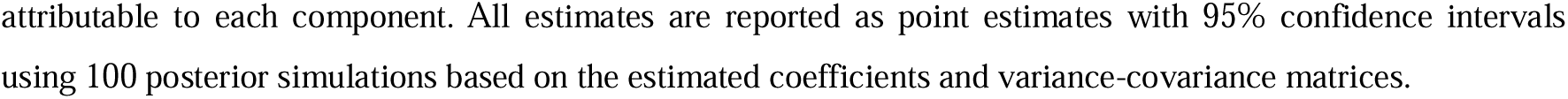
Three-way variance decomposition analysis estimating how different sources of variance contribute to the overall variance in antibiotic prescribing for each infection using a fully adjusted model (Model 1)

|  | Source of variance |  |  |
| --- | --- | --- | --- |
|  | Patient case-mix | Between-practice | Residual unexplained |
| **Upper respiratory tract infection** |  |  |  |
| Variance | 0.006<br>(0.006 – 0.006) | 0.015<br>(0.014 – 0.015) | 0.122<br>(0.121 – 0.122) |
| Proportion of total variance attributable to each component (%) | 4.1<br>(3.9 – 4.3) | 10.3<br>(10 – 10.9) | 85.6<br>(84.9 – 85.9) |
| **Acute cough** |  |  |  |
| Variance | 0.008<br>(0.008 – 0.008) | 0.014<br>(0.014 – 0.015) | 0.201<br>(0.2 – 0.202) |
| Proportion of variance explained by each source | 3.6<br>(3.4 – 3.7) | 6.2<br>(6.1 – 6.6) | 90.2<br>(89.8 – 90.4) |
| **Acute rhino sinusitis** |  |  |  |
| Variance | 0.02<br>(0.019 – 0.021) | 0.017<br>(0.017 – 0.019) | 0.213<br>(0.21 – 0.213) |
| Proportion of total variance attributable to each component (%) | 8.1<br>(7.6 – 8.5) | 6.8<br>(6.8 – 7.8) | 85.2<br>(84 – 85.3) |
| **Acute sore throat** |  |  |  |
| Variance | 0.002<br>(0.002 – 0.002) | 0.015<br>(0.015 – 0.017) | 0.223<br>(0.221 – 0.223) |
| Proportion of total variance attributable to each component (%) | 0.8<br>(0.7 – 0.9) | 6.1<br>(6.1 – 6.9) | 93.1<br>(92.2 – 93.1) |
| **Lower respiratory tract infection** |  |  |  |
| Variance | 0.005<br>(0.004 – 0.005) | 0.015<br>(0.015 – 0.016) | 0.125<br>(0.124 – 0.125) |
| Proportion of total variance attributable to each component (%) | 3.1<br>(2.9 – 3.4) | 10.5<br>(10.5 – 11.2) | 86.4<br>(85.6 – 86.5) |
| **Acute bronchitis** |  |  |  |
| Variance | 0.017<br>(0.013 – 0.021) | 0.074<br>(0.042 – 0.078) | 0.136<br>(0.131 – 0.169) |
| Proportion of total variance attributable to each component (%) | 7.3<br>(5.7 – 9.2) | 32.6<br>(18.4 – 34.4) | 60.1<br>(57.6 – 74.5) |
| **Acute otitis media** |  |  |  |
| Variance | 0.002<br>(0.001 – 0.002) | 0.024<br>(0.023 – 0.026) | 0.161<br>(0.158 – 0.162) |
| Proportion of total variance attributable to each component (%) | 1<br>(0.7 – 1.3) | 12.6<br>(12.3 – 14.2) | 86.4<br>(84.7 – 86.7) |
| **Asthma exacerbation** |  |  |  |
| Variance | 0.029<br>(0.026 – 0.032) | 0.022<br>(0.021 – 0.028) | 0.199<br>(0.191 – 0.202) |
| Proportion of total variance attributable to each component (%) | 11.4<br>(10.3 – 12.8) | 9<br>(8.2 – 11.3) | 79.6<br>(76.6 – 80.7) |
| **COPD exacerbation** |  |  |  |
| Variance | 0.008<br>(0.008 – 0.008) | 0.014<br>(0.014 – 0.015) | 0.141<br>(0.132 – 0.142) |
| Proportion of total variance attributable to each component (%) | 3.5<br>(3.1 – 6.5) | 7.2<br>(6.3 – 11.2) | 89.3<br>(83.4 – 90) |
| **Lower urinary tract infection** |  |  |  |
| Variance | 0.002<br>(0.002 – 0.002) | 0.015<br>(0.015 – 0.016) | 0.133<br>(0.132 – 0.133) |
| Proportion of total variance attributable to each component (%) | 1.4<br>(1.3 – 1.5) | 10<br>(9.9 – 10.4) | 88.6<br>(88.1 – 88.7) |
| **Impetigo** |  |  |  |
| Variance | 0.002<br>(0.002 – 0.005) | 0.014<br>(0.013 – 0.02) | 0.227<br>(0.219 – 0.227) |
| Proportion of total variance attributable to each component (%) | 0.8<br>(0.7 – 2.2) | 5.8<br>(5.3 – 8.3) | 93.4<br>(90.3 – 93.4) |

For each condition, results are presented as variance estimates for each component (patient case-mix, between-practice, and residual unexplained variation), together with the corresponding proportion of total variance
attributable to each component. All estimates are reported as point estimates with 95% confidence intervals using 100 posterior simulations based on the estimated coefficients and variance-covariance matrices.

When comparing results from Model 1 (full case-mix adjustment) and Model 3 (no case-mix adjustment), we found that not adjusting for patient case-mix overstated the contribution of between-GP differences. While the absolute contribution of between-practice variance to total variance changed only modestly after case-mix adjustment (Figure 2A and *Appendix 7*), its contribution to the explained variance (considering only patient case-mix and between-practice variance components) changed substantially (Figure 2B and *Appendix 7*). In Model 3, between-practice differences constituted the sole explained variance component. After including patient case-mix in Model 1, the contribution of between-practice differences was reduced by more than 20pp for six conditions, falling below 50% for rhinitis (45.7%) and asthma exacerbation (43.9%), and to between 50% and 80% for URTI (71.5%), cough (63.6%), LRTI (77.1%), and COPD (67.1%). Meanwhile, minimal age-sex adjustment, akin the STAR-PU approach (Model 2), did not make a noticeable difference compared with no adjustment (Model 3). For 8 of the 11 infections, the total explained variance attributable to between-practice differences changed by less than 5pp relative to the unadjusted models when adjusting only for age and sex (Figure 2B and *Appendix 7*).

**Figure 2.**
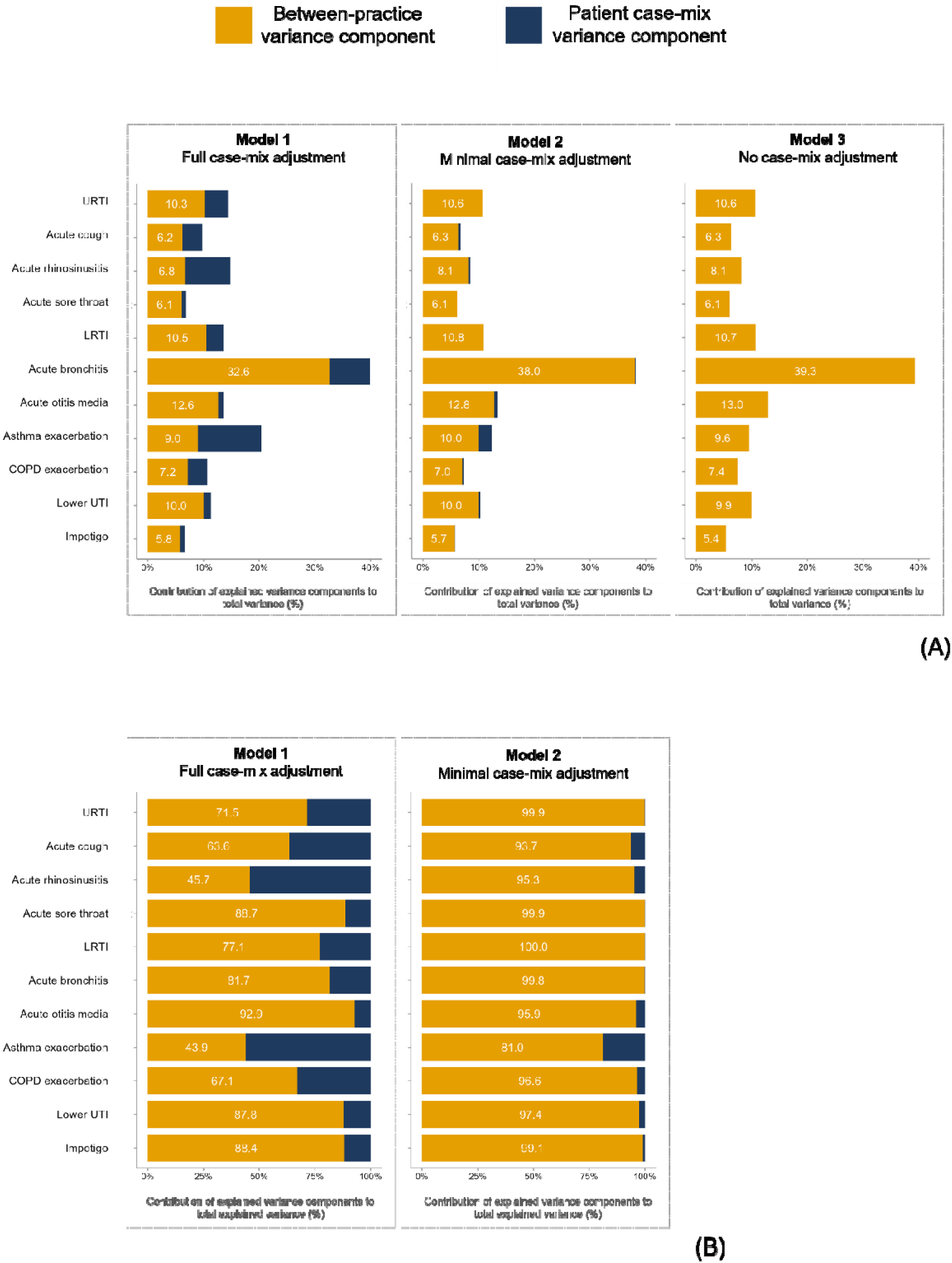
Contributions of explained variance components (between-practice variance and patient case-mix) across three models with different levels of case-mix adjustment. Panel **A** shows contributions to total variance (the sum of patient case-mix variance, between-practice variance, and residual unexplained variance). Panel **B** shows relative contributions to total **explained** variance (the sum of patient case-mix variance and between-practice variance). In both panels, percentages are shown only for the between-practice variance component in each bar to highlight how limited case-mix adjustment could overstate the contribution of between-practice differences, and how this was reduced as additional case-mix factors were included. In Panel B, Results from Model 3 are not shown because, in the absence of case-mix adjustment, all explained variance is attributable to between-practice differences (100%). (See *Appendix 7* for detailed results, including point estimates and 95% confidence intervals for each variance component.)

However, a high proportion of total variance remained unexplained across all infections studied, ranging from 60.1% for acute bronchitis to 93.4% for impetigo (Table 3).

### Sensitivity analysis with infection severity

Simulating the effect of unmeasured infection severity of sore throat and LRTI in adults substantially shifted variance components (Table 4). For sore throat, contribution of case-mix variance component to total variance increased from 0.9% to 11.3% and the between-practice component attenuated by 14%, decreasing from 5.9% to 5.4%. For LRTI, accounting for infection severity reduced the contribution of the between-practice component even further, with a relative reduction of 25.9%, from 11.2% to 8.3%. These simulations indicated that unmeasured severity might explain a substantial fraction of what otherwise appeared to be practice-level variability.

**Table 4.** Sensitivity analysis of variance components for antibiotic prescribing for patients aged ≥16 years with sore throat and LRTI, before and after severity imputation.

|  | Model without<br>severity imputation (baseline) <sup>a</sup> |  |  | Model with<br>severity imputation <sup>b</sup> |  |  |
| --- | --- | --- | --- | --- | --- | --- |
|  | Source of variance |  |  | Source of variance |  |  |
|  | Patient case-mix | Between-GP | Residual | Patient case-mix | Between-GP | Residual |
| **Acute sore throat** |  |  |  |  |  |  |
| Variance | 0.002<br>(0.002 – 0.003) | 0.015<br>(0.015 – 0.017) | 0.223<br>(0.22 – 0.223) | 0.027<br>(0.026; 0.028) | 0.013<br>(0.013; 0.013) | 0.200<br>(0.199; 0.201) |
| Proportion of total variance<br>attributable to each component<br>(%) | 0.9<br>(0.8 – 1.1) | 5.9<br>(5.9 – 7) | 93.2<br>(92 – 93.2) | 11.3<br>(10.8; 11.7) | 5.4<br>(5.3; 5.5) | 83.4<br>(82.9; 83.8) |
| **LRTI** |  |  |  |  |  |  |
| Variance | 0.002<br>(0.002 – 0.002) | 0.015<br>(0.015 – 0.016) | 0.116<br>(0.115 – 0.117) | 0.022<br>(0.021; 0.022) | 0.011<br>(0.011; 0.011) | 0.101 (0.100;<br>0.101) |
| Proportion of total variance<br>attributable to each component<br>(%) | 1.6<br>(1.4 – 1.8) | 11.2<br>(11 – 12) | 87.2<br>(86.4 – 87.4) | 16.3<br>(15.8; 16.7) | 8.3<br>(8.1; 8.4) | 75.5<br>(75.0; 75.9) |

For each infection, results are presented as variance estimates for each component (patient case-mix, between-practice, and residual unexplained variation), together with the corresponding proportion of total variance attributable to each component (%). All estimates are reported as point estimates with 95% confidence intervals. a: Models without severity imputation (baseline), where patient case-mix factors were included as in the full case-mix adjustment models (Model 1). b: Models incorporating severity imputation, where consultation-level infection severity was included in addition to the full case-mix adjustment patient case-mix factors. Severity was imputed using a simulation-based approach using the joint probability distribution of infection severity conditional on both GP practice prescribing quartile and prescribing decision (*Appendix 6*). 100 imputations were generated from a Dirichlet distribution parameterised by fitted cell counts within each practice-prescribing group. Variance decomposition was performed in each replicate; point estimates are the mean across imputations, with 95% uncertainty intervals derived from the empirical distribution of simulated estimates.

## DISCUSSION

Using large population-based patient-level data, we found that antibiotic prescribing for common infections in UK primary care settings remained very high. For many conditions, such as cough, sinusitis, sore throat, bronchitis, and otitis media, where rates of antibiotic prescribing ideally remain below 20%, we observed actual proportions ranging from 30% to 70%.^5,23,24^ We also observed substantial variation in antibiotic prescribing between practices across most infections studied. This highlights the importance of identifying the factors underlying prescribers’ antibiotic prescribing decisions, which ultimately drove differences between practices.

Across infections, between-practice differences explained more of the total prescribing variance than patient case-mix, although the largest proportion of variance remained unexplained. Compared with no case-mix adjustment, the fully case-mix adjusted model reduced between-practice differences, providing a fairer basis for comparing prescribing performance. In contrast, minimal adjustment, reflecting the age-sex weighting used in STAR-PU, had almost no effect relative to no case-mix adjustment. Our sensitivity analyses suggested that accounting for infection severity would markedly change the contribution of the between-GP component, with the largest change for LRTI where apparent between-GP variation was approximately halved.

Our findings are consistent with previous studies showing large and persistent variability of antibiotic prescribing in primary care in England. Using CPRD data from 2000 to 2015, Palin et al. reported considerable variation in prescribing between practices, which increased over time, e.g. ranging from 10% to 70% across practices for URTI.^25^ Three large prospective cohort studies conducted by Stuart et al. confirmed this pattern, reporting a wide range in antibiotic prescribing for adults with sore throat (0–97%) and LRTIs (7–100%), and for children with respiratory infections (0–75%).^14^ Pouwels et al. found that comorbidity prevalences explained little practice-level variation in antibiotic prescribing.^26^ Several qualitative studies have highlighted non-clinical factors, such as perceived patient expectations and local practice culture, but their quantitative contribution remains unclear.^27^

Our study shows the importance of condition-specific analysis of prescribing in primary care, as decomposition analyses revealed that the contribution of variance components differed substantially between conditions. Comprehensive case-mix adjustment is important for fair comparisons of GP prescribing performance,^11^ particularly for conditions such as LRTI where diagnostic uncertainty is high and patients vary widely in their risk of adverse outcomes. In these contexts, a substantial proportion of patients are unlikely to benefit from antibiotics, while a smaller group may require treatment to prevent complications such as hospital admission. Without adequate adjustment, variation in prescribing may be incorrectly attributed to practice norms or prescriber behaviour, instead of differences in patient populations. In contrast, for more discrete and guideline-driven conditions, such as otitis media, case-mix adjustment had limited impact on between-practice variation, suggesting that residual variation may more strongly reflect differences in prescribing behaviour. Robust adjustment for case-mix is not only essential for fair comparisons but also for effectiveness of benchmarking, as GPs are more likely to respond positively to comparisons they perceive as fair and reflective of differences in case-mix.^12^

Our sensitivity analyses illustrate that not accounting for infection severity may lead to substantial overestimation of the influence of prescriber behaviour, particularly for LRTI, a condition marked by diagnostic uncertainty. In primary care, LRTI may represent either acute bronchitis, which rarely requires antibiotics, or pneumonia, which always does. Consequently, a pneumonia risk index represents a particularly important case-mix factor influencing prescribing decisions. However, similarly strong severity indices are not available for most common infections because clinical signs and symptoms do not clearly distinguish mild from severe disease. This is commonly the case for infections that rarely require antibiotic treatment, such as rhinosinusitis, for which severity assessment relies largely on individual prescriber judgement, making adjustment for infection severity indices less feasible.

Beyond explaining variability, our findings help identify infections that may serve as practical indicators of antibiotic prescribing quality. Acute otitis media and URTI showed substantial between-practice variation after accounting for case-mix, making them promising conditions for benchmarking prescribing behaviour. By contrast, although acute bronchitis showed the greatest between-practice variation, diagnostic uncertainty and coding practices, where pneumonia or more severe cases are often coded as LRTI or pneumonia when antibiotics are prescribed, limit its suitability as a prescribing quality indicator. For LRTI and UTI, the more limited variation in antibiotic prescribing percentages between GP practices offered limited ability to distinguish between higher- and lower-prescribing practices and therefore reduced their usefulness as indicators of prescribing quality.

Our study has important implications for antibiotic stewardship in primary care. It shows that practice behaviour and prescribing norm, independent of case-mix variations, play an important role and highlight the potential value of targeted feedback at the practice level. The observed variation between conditions highlights the value of our condition-specific approach. By comparing different case-mix adjustment strategies, we provide an empirical basis for developing fairer benchmarks of GP prescribing performance. Although data on severity were unavailable, our imputation-based sensitivity analyses using external data indicate the likely influence of this factor.

Our study has some limitations. Case-mix will still not have been fully adjusted for, as the severity of current infection remains unmeasured, as well as other unmeasured factors. However, we quantified the potential impact of severity of infection in sensitivity analyses for the two conditions for which external evidence on the distribution of severity of illness exists. We assumed constant effects of patient covariates across practices. Given that measured case-mix explained only a small share of total variance, large, widespread patient-practice interactions are unlikely to play an important role. We did not quantify variation between individual clinicians, as this data was not available: this may partly explain the substantial residual variance. However, although prescribing decisions are made by individual GPs, benchmarking is usually conducted at the practice level, and some practices consistenly exhibit higher overall prescribing levels than others. In this context, estimating practice-level prescribing propensity aligns with how performance is monitored and acted upon in routine stewardship programmes. Furthermore, for binary outcomes with moderate prevalence, residual variance is necessarily large even in the absence of presciber heterogeneity. Unless prescribing decisions for most patients can be predicted deterministically from measured characteristics, substantial within-practice variability will remain. Therefore, any additional contribution from differences between individual prescribers is likely to be limited relative to total variance.

In conclusion, our study shows that antibiotic prescribing for common infections in primary care remains high and varies considerably between general practices. The extent to which this variation reflects differences in case-mix varies by condition. Comprehensive case-mix adjustment is therefore essential to avoid overestimating the role of prescribing behaviour and to enable fair benchmarking of prescribing performance.

Our condition-specific analyses highlight acute otitis media and URTI as conditions with substantial prescribing variation after case-mix adjustment, suggesting that prescribing differences are more likely to be modifiable and making them promising indicators of antibiotic prescribing quality.

## Data Availability

Electronic health records are classified as sensitive data in the UK under the Data Protection Act and therefore cannot be publicly shared due to information governance requirements designed to protect patient confidentiality. Access to Clinical Practice Research Datalink (CPRD) data requires protocol approval through its research data governance process (see https://cprd.com/data-access for details). Linked datasets, including secondary care data from Hospital Episodes Statistics, mortality data from Office for National Statistics, and Index of Multiple Deprivation data, can also be requested via CPRD.

## Acknowledgements

This study was funded by the Wellcome grant ADILA (Antibiotic Data to Inform Local Action) [222051/Z/20/Z] and the National Institute for Health and Care Research (NIHR) Health Protection Research Unit in Healthcare Associated Infections and Antimicrobial Resistance (NIHR207397), a partnership between the UK Health Security Agency (UKHSA) and the University of Oxford. A.S.W. is an NIHR Senior Investigator and is also supported by the NIHR Oxford Biomedical Research Centre. K.B.P. is also supported by the Medical Research Foundation (MRF-160-0017-ELP-POUW-C0909). The views expressed are those of the authors and not necessarily those of the NIHR, UKHSA or the Department of Health and Social Care.

## Contributors

KBP conceptualised the study. KBP and NVN designed the study, with input from ASW and DWE. NVN and KBP had access to the raw data. NVN prepared the data for analysis, conducted the analyses, and drafted the first version of the manuscript under the supervision of KBP. KBP supervised all stages of the study. MS, SW, JVR, and KBP acquired funding. All authors critically reviewed and edited the manuscript and approved the final version. NVN and KBP act as guarantors.

## Ethics

This study was approved by the Clinical Practice Research Datalink’s (CPRD) independent scientific advisory committee (protocol 23_003072).

