## Supplementary for "Explaining variation in antibiotic prescribing for common infections: a three-way variance decomposition using UK primary care data"

**Appendix 1.** Medcodeid for infections of interest

| **Medcodeid** | **Term** | **Infections** |
| --- | --- | --- |
| 18268014 | Acute bronchitis | acute bronchitis |
| 58909014 | Acute tracheobronchitis | acute bronchitis |
| 142425010 | Laryngotracheobronchitis | acute bronchitis |
| 301095014 | Acute bronchitis and bronchiolitis | acute bronchitis |
| 301100011 | Acute fibrinous bronchitis | acute bronchitis |
| 301101010 | Acute membranous bronchitis | acute bronchitis |
| 301103013 | Acute purulent bronchitis | acute bronchitis |
| 301105018 | Acute pneumococcal bronchitis | acute bronchitis |
| 301106017 | Acute streptococcal bronchitis | acute bronchitis |
| 301108016 | Acute haemophilus influenzae bronchitis | acute bronchitis |
| 301109012 | Acute Neisseria catarrhalis bronchitis | acute bronchitis |
| 301116013 | Acute bronchitis due to rhinovirus | acute bronchitis |
| 301119018 | Subacute bronchitis | acute bronchitis |
| 301120012 | Acute viral bronchitis | acute bronchitis |
| 301121011 | Acute bacterial bronchitis | acute bronchitis |
| 301122016 | Acute bronchitis NOS | acute bronchitis |
| 301132011 | Acute bronchitis or bronchiolitis NOS | acute bronchitis |
| 301437010 | Tracheobronchitis | acute bronchitis |
| 301441014 | Bronchitis NOS | acute bronchitis |
| 301820014 | Acute infective bronchitis | acute bronchitis |
| 350041018 | Chest infection - unspecified bronchitis | acute bronchitis |
| 350044014 | Acute bronchitis due to mycoplasma pneumoniae | acute bronchitis |
| 396107011 | Bronchitis | acute bronchitis |
| 411490016 | Acute wheezy bronchitis | acute bronchitis |
| 2163183015 | Wheezy bronchitis | acute bronchitis |
| 2475601016 | Acute pseudomembranous bronchitis | acute bronchitis |
| 2475602011 | Acute croupous bronchitis | acute bronchitis |
| 3514606010 | Protracted bacterial bronchitis | acute bronchitis |
| 3514603019 | Acute noninfective bronchitis | acute bronchitis |
| 455991000006112 | Acute bronchitis due to coxsackievirus | acute bronchitis |
| 456001000006113 | Acute bronchitis due to echovirus | acute bronchitis |
| 456021000006115 | Acute bronchitis due to parainfluenza virus | acute bronchitis |
| 456031000006117 | Acute bronchitis due to respiratory syncytial virus | acute bronchitis |
| 885041000006119 | Acute bronchitis/bronchiolitis | acute bronchitis |
| 907481000006118 | [RFC] Bronchitis | acute bronchitis |
| 1174251000000120 | Aspergillus bronchitis | acute bronchitis |
| 1709191000006110 | Aspergillus bronchitis | acute bronchitis |
| 2756071000006110 | Viral bronchitis | acute bronchitis |
| 3896411000006110 | LTB - Laryngotracheobronchitis | acute bronchitis |
| 3955621000006110 | Catarrhal bronchitis | acute bronchitis |
| 4780421000006110 | Acute Moraxella catarrhalis bronchitis | acute bronchitis |
| 4780481000006120 | Acute parainfluenza virus bronchitis | acute bronchitis |
| 4780491000006120 | Acute respiratory syncytial virus bronchitis | acute bronchitis |
| 5053331000006120 | Acute mycoplasmal bronchitis | acute bronchitis |
| 5573791000006110 | Acute fibrinous laryngotracheobronchitis | acute bronchitis |
| 6015831000006120 | Acute infective tracheobronchitis | acute bronchitis |
| 6720721000006110 | Asthmatic bronchitis | acute bronchitis |
| 7715861000006110 | Acute bronchitis co-occurrent with bronchiectasis | acute bronchitis |
| 12717061000006100 | Acute bronchitis and/or bronchiolitis | acute bronchitis |
| 13486511000006100 | Acute bronchitis caused by SARS-CoV-2 (severe acute respiratory syndrome coronavirus 2) | acute bronchitis |
| 13486521000006100 | COVID-19 acute bronchitis | acute bronchitis |
| 13651301000006100 | Bronchitis co-occurrent with wheeze | acute bronchitis |
| 13651611000006100 | Acute bronchitis co-occurrent with wheeze | acute bronchitis |
| 317403015 | [D]Cough | acute cough |
| 317408012 | [D]Cough with haemorrhage | acute cough |
| 2168391000000120 | [D]Episodic dry cough | acute cough |
| 30371011 | Barking cough | acute cough |
| 3193601000006110 | Bout of coughing | acute cough |
| 2828791000006110 | Brassy cough | acute cough |
| 252360013 | Bronchial cough | acute cough |
| 13768751000006100 | Bronchial cough | acute cough |
| 407081018 | C/O - cough | acute cough |
| 5259521000006120 | Character of cough | acute cough |
| 252359015 | Chesty cough | acute cough |
| 13768761000006100 | Chesty cough | acute cough |
| 5533011000006110 | Complaining of cough | acute cough |
| 1970531000006110 | COPD assessment test score - cough | acute cough |
| 82824016 | Cough | acute cough |
| 961781000006119 | Cough | acute cough |
| 479311018 | Cough on exercise | acute cough |
| 1709331000006110 | Cough on exercise | acute cough |
| 252366019 | Cough symptom NOS | acute cough |
| 317404014 | Cough syncope | acute cough |
| 6782071000006110 | Cough variant asthma | acute cough |
| 5259531000006110 | Cough when swallowing | acute cough |
| 216653013 | Cough with fever | acute cough |
| 5468241000006110 | Coughing | acute cough |
| 6357671000006110 | Coughing and deep breathing | acute cough |
| 3587071000006110 | Coughing up blood | acute cough |
| 598431000006113 | Coughing up phlegm | acute cough |
| 5259541000006110 | Coughs when swallowing | acute cough |
| 1780331000006120 | Daytime cough | acute cough |
| 5754161000006110 | Croupy cough | acute cough |
| 5741451000006120 | Does cough | acute cough |
| 5890991000006120 | Does cough up sputum | acute cough |
| 20419011 | Dry cough | acute cough |
| 5890901000006110 | Effective cough | acute cough |
| 8260311000006110 | Episodic dry cough | acute cough |
| 252364016 | Evening cough | acute cough |
| 3474321000006120 | Hacking cough | acute cough |
| 3520051000006110 | Increasing frequency of cough | acute cough |
| 2961941000006110 | Loose cough | acute cough |
| 2961951000006110 | Moist cough | acute cough |
| 252363010 | Morning cough | acute cough |
| 252357018 | Night cough present | acute cough |
| 3516981000006110 | Nocturnal cough | acute cough |
| 252406018 | Nocturnal cough / wheeze | acute cough |
| 6781071000006110 | Non-productive cough | acute cough |
| 11823221000006100 | Non-productive cough | acute cough |
| 3304991000006120 | Observation of cough | acute cough |
| 5243651000006110 | Painful cough | acute cough |
| 3193591000006110 | Paroxysmal cough | acute cough |
| 423230012 | Persistent cough | acute cough |
| 7305051000006120 | Persistent cough after viral respiratory infection | acute cough |
| 7305041000006120 | Postviral cough | acute cough |
| 397882011 | Productive cough | acute cough |
| 252351017 | Productive cough -clear sputum | acute cough |
| 252352012 | Productive cough -green sputum | acute cough |
| 252353019 | Productive cough-yellow sputum | acute cough |
| 3524231000006120 | Spasmodic cough | acute cough |
| 459738011 | Unexplained cough | acute cough |
| 411654018 | Sputum evidence of infection | acute cough |
| 414646016 | Yellow sputum | acute cough |
| 414647013 | Green sputum | acute cough |
| 131721000006111 | Sputum appears infected | acute cough |
| 1834111000006110 | Dark green sputum | acute cough |
| 1834121000006120 | Pale green sputum | acute cough |
| 2226161000000120 | Dark green sputum | acute cough |
| 2226201000000110 | Pale green sputum | acute cough |
| 3180091000006120 | Purulent sputum | acute cough |
| 4609991000006110 | Identification of organism on gram stain of sputum | acute cough |
| 5562961000006110 | Pus in sputum O/E | acute cough |
| 9528331000006120 | Sputum culture positive for Pseudomonas | acute cough |
| 9610391000006120 | Sputum culture positive for Aspergillus | acute cough |
| 9613381000006110 | Respiratory syncytial virus positive sputum culture | acute cough |
| 14738014 | Acute allergic mucoid otitis media | acute otitis media |
| 22934019 | Postmeasles otitis media | acute otitis media |
| 30177017 | Acute allergic sanguinous otitis media | acute otitis media |
| 58707018 | Acute otitis media with effusion | acute otitis media |
| 87124011 | Acute non-suppurative otitis media - mucoid | acute otitis media |
| 128622017 | Acute sanguinous otitis media | acute otitis media |
| 299014013 | Acute non-suppurative otitis media - serous | acute otitis media |
| 299040019 | Serous otitis media | acute otitis media |
| 299041015 | Catarrhal otitis media | acute otitis media |
| 299042010 | Mucoid otitis media | acute otitis media |
| 299061014 | Acute suppurative otitis media | acute otitis media |
| 299065017 | Acute suppurative otitis media due to disease EC | acute otitis media |
| 299066016 | Acute suppurative otitis media NOS | acute otitis media |
| 299068015 | Purulent otitis media NOS | acute otitis media |
| 299069011 | Bilateral suppurative otitis media | acute otitis media |
| 299071011 | Acute left otitis media | acute otitis media |
| 299072016 | Acute right otitis media | acute otitis media |
| 299073014 | Acute bilateral otitis media | acute otitis media |
| 299529015 | [X]Other chronic suppurative otitis media | acute otitis media |
| 299530013 | [X]Otitis media in bacterial diseases classified elsewhere | acute otitis media |
| 299532017 | [X]Otitis media in other diseases classified elsewhere | acute otitis media |
| 473798017 | Acute secretory otitis media | acute otitis media |
| 77591000006116 | Non-suppurative otitis media | acute otitis media |
| 116801000006110 | Purulent otitis media | acute otitis media |
| 297571000006118 | Non-suppurative otitis media with eustachian tube disorders | acute otitis media |
| 297581000006115 | Nonsuppurative otitis media NOS | acute otitis media |
| 406091000006110 | [X]Other acute nonsuppurative otitis media | acute otitis media |
| 458531000006114 | Acute non-suppurative otitis media | acute otitis media |
| 458581000006110 | Acute nonsuppurative otitis media NOS | acute otitis media |
| 460541000006114 | Acute suppurative otitis media - tympanic membrane intact | acute otitis media |
| 460551000006111 | Acute suppurative otitis media - tympanic membrane ruptured | acute otitis media |
| 555791000006118 | Otitis media with effusion - serous | acute otitis media |
| 883681000006115 | Acute nonsupp. otitis media | acute otitis media |
| 906681000006117 | [RFC] Otitis media | acute otitis media |
| 1806411000006120 | Acute otitis media | acute otitis media |
| 1991671000006110 | At risk of otitis media complication | acute otitis media |
| 1991681000006110 | High risk of otitis media complication | acute otitis media |
| 2548641000006110 | AOM - Acute otitis media | acute otitis media |
| 2712661000006120 | Post measles otitis media | acute otitis media |
| 2736971000006110 | Acute suppurative otitis media without spontaneous rupture of ear drum | acute otitis media |
| 3118471000006110 | Otitis media with effusion - purulent | acute otitis media |
| 3132041000006120 | Suppurative otitis media | acute otitis media |
| 3348911000006110 | Acute mucoid otitis media | acute otitis media |
| 3562731000006110 | OM - Otitis media | acute otitis media |
| 3760591000006120 | Acute sanguinous otitis media | acute otitis media |
| 3760601000006110 | Acute non-suppurative otitis media with haemotympanum | acute otitis media |
| 3760621000006120 | Acute non-suppurative otitis media - sanguinous | acute otitis media |
| 3760631000006120 | Acute non-suppurative otitis media - bloody | acute otitis media |
| 3782571000006120 | OME - Otitis media with effusion | acute otitis media |
| 3782601000006110 | Otitis media with effusion | acute otitis media |
| 3782621000006120 | Secretory otitis media | acute otitis media |
| 3902951000006110 | Acute suppurative otitis media with spontaneous rupture of ear drum | acute otitis media |
| 3902971000006110 | Acute suppurative otitis media with discharge | acute otitis media |
| 4774551000006110 | Acute serous otitis media | acute otitis media |
| 4774691000006120 | ASOM - Acute suppurative otitis media | acute otitis media |
| 5033831000006110 | Otitis media with effusion - sanguinous | acute otitis media |
| 6013861000006110 | Infective otitis media | acute otitis media |
| 6204261000006120 | Acute nonsuppurative otitis media | acute otitis media |
| 7987351000006120 | Acute bilateral otitis media with effusion | acute otitis media |
| 7987391000006110 | Perforation of tympanic membrane due to otitis media | acute otitis media |
| 8025131000006110 | Acute persistent otitis media | acute otitis media |
| 13012241000006100 | Otitis media caused by SARS-CoV-2 (severe acute respiratory syndrome coronavirus 2) | acute otitis media |
| 13483771000006100 | Otitis media due to disease caused by SARS-CoV-2 (severe acute respiratory syndrome coronavirus 2) | acute otitis media |
| 13483791000006100 | Otitis media due to COVID-19 | acute otitis media |
| 14175821000006100 | Acute mucoid otitis media of right middle ear | acute otitis media |
| 130848015 | Glue ear | acute otitis media |
| 3826261000006120 | Glue ear - serous | acute otitis media |
| 780361000006114 | Ear infection | acute otitis media |
| 255651016 | O/E - tympanic membrane pink | acute otitis media |
| 255652011 | O/E - tympanic membrane red | acute otitis media |
| 255653018 | O/E -tympanic membrane bulging | acute otitis media |
| 399840017 | Catarrh - eustachian | acute otitis media |
| 82051019 | Aero-otitis media | acute otitis media |
| 3296881000006110 | Aero-otitis media | acute otitis media |
| 301801012 | [X]Other acute sinusitis | acute rhinosinusitis |
| 488101000006116 | Acute antritis | acute rhinosinusitis |
| 3727741000006110 | Acute bacterial sinusitis | acute rhinosinusitis |
| 112674013 | Acute ethmoidal sinusitis | acute rhinosinusitis |
| 150861013 | Acute frontal sinusitis | acute rhinosinusitis |
| 113403018 | Acute maxillary sinusitis | acute rhinosinusitis |
| 9421011 | Acute pansinusitis | acute rhinosinusitis |
| 567061000000117 | Acute rhinosinusitis | acute rhinosinusitis |
| 26785019 | Acute sinusitis | acute rhinosinusitis |
| 301015010 | Acute sinusitis NOS | acute rhinosinusitis |
| 129311019 | Acute sphenoidal sinusitis | acute rhinosinusitis |
| 146816010 | Aerosinusitis | acute rhinosinusitis |
| 7576131000006110 | Bacterial sinusitis | acute rhinosinusitis |
| 253249011 | Blocked sinuses | upper respiratory tract infection |
| 2795871000006120 | Ethmoidal sinusitis | acute rhinosinusitis |
| 301209018 | Fistula of nasal sinus | acute rhinosinusitis |
| 130650014 | Frontal sinusitis | acute rhinosinusitis |
| 8032501000006120 | Fungal sinusitis | acute rhinosinusitis |
| 146480018 | Maxillary sinusitis | acute rhinosinusitis |
| 301013015 | Other acute sinusitis | acute rhinosinusitis |
| 301014014 | Other acute sinusitis NOS | acute rhinosinusitis |
| 301212015 | Pansinusitis | acute rhinosinusitis |
| 14149851000006100 | Rhinosinusitis | acute rhinosinusitis |
| 61668014 | Sinusitis | acute rhinosinusitis |
| 8016801000006110 | Sinusitis co-occurrent with nasal polyps | acute rhinosinusitis |
| 2710431000006110 | Sphenoidal sinusitis | acute rhinosinusitis |
| 5035671000006110 | Suppurative sinusitis with complications | acute rhinosinusitis |
| 7298311000006110 | Viral sinusitis | acute rhinosinusitis |
| 3838761000006110 | Congestion of nasal sinus | upper respiratory tract infection |
| 136509019 | Sinus congestion | upper respiratory tract infection |
| 407076010 | C/O nasal congestion | upper respiratory tract infection |
| 113345017 | Nasal congestion | upper respiratory tract infection |
| 317324014 | [D]Nasal obstruction | acute rhinosinusitis |
| 6349471000006110 | Nasal sinus obstruction | acute rhinosinusitis |
| 5498541000006110 | Nasal obstruction present | acute rhinosinusitis |
| 397997010 | Nasal obstruction present | acute rhinosinusitis |
| 681631000006110 | Nasal obstruction | acute rhinosinusitis |
| 517051000006111 | Nasal obstruction | acute rhinosinusitis |
| 347922010 | Nasal obstruction | acute rhinosinusitis |
| 5033361000006110 | Nasal airway obstruction | acute rhinosinusitis |
| 5033341000006110 | NO - Nasal obstruction | acute rhinosinusitis |
| 317269017 | [D]Throat pain | acute sore throat |
| 301831018 | Abscess of pharynx | acute sore throat |
| 4780151000006110 | Acute exudative tonsillitis | acute sore throat |
| 5035771000006110 | Acute herpes simplex pharyngitis | acute sore throat |
| 5036091000006110 | Acute infection of tonsillar remnant | acute sore throat |
| 5912901000006120 | Acute lingual tonsillitis | acute sore throat |
| 136470019 | Acute nasopharyngitis | acute sore throat |
| 4075191000006110 | Acute peritonsillitis | acute sore throat |
| 5036301000006120 | Adult acute epiglottitis and supraglottitis | upper respiratory tract infection |
| 7576111000006120 | Bacterial tonsillitis | acute sore throat |
| 7267891000006120 | Bilateral tonsillar swelling | acute sore throat |
| 5035831000006110 | Chlamydial pharyngitis | acute sore throat |
| 2525451000006120 | Drainage of abscess of tonsil | acute sore throat |
| 1486215014 | Drainage of peritonsillar abscess | acute sore throat |
| 267812011 | Drainage of retropharyngeal abscess | acute sore throat |
| 5894161000006110 | Enlarged tonsil | acute sore throat |
| 5271161000006120 | Enlarged tonsils | acute sore throat |
| 14165541000006100 | Enlargement of left tonsil | acute sore throat |
| 348208015 | Enlargement of tonsil or adenoid | acute sore throat |
| 5035801000006120 | Glandular fever pharyngitis | acute sore throat |
| 3708981000006110 | Gonococcal pharyngitis | acute sore throat |
| 253225013 | Has a sore throat | acute sore throat |
| 7819881000006110 | Herpes simplex pharyngotonsillitis | acute sore throat |
| 6530801000006110 | Incision and drainage of peritonsillar abscess | acute sore throat |
| 6016171000006110 | Infective pharyngitis | acute sore throat |
| 5655881000006120 | Inflamed tonsils | acute sore throat |
| 5894171000006110 | Large tonsil | acute sore throat |
| 5271151000006120 | Large tonsils | acute sore throat |
| 1984341000006110 | Manchester triage - Sore throat | acute sore throat |
| 402541013 | O/E - pharynx hyperaemic | acute sore throat |
| 255674017 | O/E - tonsils - quinsy present | acute sore throat |
| 411841010 | O/E - tonsils enlarged | acute sore throat |
| 255673011 | O/E - tonsils grossly enlarged | acute sore throat |
| 255671013 | O/E - tonsils hyperaemic | acute sore throat |
| 4577581000006110 | On examination - tonsils - quinsy present | acute sore throat |
| 255672018 | On examination - tonsils moderately enlarged | acute sore throat |
| 253240010 | Pain in throat | acute sore throat |
| 140751018 | Parapharyngeal abscess | acute sore throat |
| 229081000006112 | Peritonsillar abscess | acute sore throat |
| 862301000006114 | Peritonsillar abscess drainage | acute sore throat |
| 6028741000006110 | Peritonsillar abscess drained | acute sore throat |
| 885111000006119 | Peritonsillar abscess -quinsey | acute sore throat |
| 411477011 | Persistent sore throat | acute sore throat |
| 14130751000006100 | Pharyngotonsillitis | acute sore throat |
| 301300017 | Pharynx or nasopharynx abscess | acute sore throat |
| 4780231000006120 | RAT - Recurrent acute tonsillitis | acute sore throat |
| 1805641000006110 | Red throat | acute sore throat |
| 30574019 | Retropharyngeal abscess | acute sore throat |
| 3206351000006110 | Septic sore throat | acute sore throat |
| 139761000006116 | Sore throat | acute sore throat |
| 4548281000006110 | Sore throat present | acute sore throat |
| 398001015 | Sore throat symptom | acute sore throat |
| 253227017 | Sore throat symptom NOS | acute sore throat |
| 3206381000006110 | Strep throat | acute sore throat |
| 879051000006113 | Strep.throat + scarlatina NOS | acute sore throat |
| 3206371000006120 | Strept throat | acute sore throat |
| 73157016 | Streptococcal sore throat | acute sore throat |
| 286457014 | Streptococcal sore throat NOS | acute sore throat |
| 121591000006115 | Streptococcal sore throat with scarlatina | acute sore throat |
| 286458016 | Streptococcal sore throat with scarlatina NOS | acute sore throat |
| 7266681000006120 | Swelling of right tonsil | acute sore throat |
| 6974361000006120 | Swelling of throat | acute sore throat |
| 7263731000006120 | Swelling of tonsil | acute sore throat |
| 100101000006117 | Throat infection - pharyngitis | acute sore throat |
| 100111000006119 | Throat infection - tonsillitis | acute sore throat |
| 253247013 | Throat irritation | acute sore throat |
| 253237010 | Throat pain | acute sore throat |
| 2164212016 | Throat soreness | acute sore throat |
| 5886851000006120 | Tonsillar adenitis | acute sore throat |
| 3254181000006120 | Tonsillar enlargement | acute sore throat |
| 12717051000006100 | Tracheopharyngitis | acute sore throat |
| 93954019 | Tuberculosis of nasopharynx | acute sore throat |
| 5036081000006110 | Vincent tonsillitis | acute sore throat |
| 4743831000006120 | Vincent's angina - pharyngitis | acute sore throat |
| 348206016 | Vincent's tonsillitis | acute sore throat |
| 3329901000006120 | Viral tonsillitis | acute sore throat |
| 1761951000006120 | [SHHAPT] Gonorrhoea - pharyngeal infection | acute sore throat |
| 1762031000006120 | [SHHAPT] Chlamydial infection - pharyngeal | acute sore throat |
| 1762091000006120 | [SHHAPT] Chlamydial infection - pharyngeal - medication given | acute sore throat |
| 1762011000006110 | [SHHAPT] Gonorrhoea - pharyngeal infection - medication given | acute sore throat |
| 1762081000006120 | [SHHAPT] Chlamydial infection-pharyngeal-diag. prev. elsewhere | acute sore throat |
| 1762001000006110 | [SHHAPT] Gonorrhoea - pharyngeal - diagnosed prev. elsewhere | acute sore throat |
| 1762111000006110 | [SHHAPT] Lymphogranuloma venereum - pharyngeal infection | acute sore throat |
| 301802017 | [X]Acute pharyngitis due to other specified organisms | acute sore throat |
| 3880611000006110 | Abscess of parapharyngeal space | acute sore throat |
| 3880621000006120 | Abscess of lateral pharyngeal space | acute sore throat |
| 301024018 | Acute bacterial pharyngitis | acute sore throat |
| 301027013 | Acute bacterial pharyngitis NOS | acute sore throat |
| 301021014 | Acute gangrenous pharyngitis | acute sore throat |
| 92029013 | Acute laryngopharyngitis | acute sore throat |
| 5035851000006120 | Acute pharyngeal candidiasis | acute sore throat |
| 486416017 | Acute pharyngitis | acute sore throat |
| 459581000006119 | Acute pharyngitis NOS | acute sore throat |
| 301022019 | Acute phlegmonous pharyngitis | acute sore throat |
| 301025017 | Acute pneumococcal pharyngitis | acute sore throat |
| 301026016 | Acute staphylococcal pharyngitis | acute sore throat |
| 855671000006111 | Acute tonsilitis/pharyngitis | acute sore throat |
| 301023012 | Acute ulcerative pharyngitis | acute sore throat |
| 301028015 | Acute viral pharyngitis | acute sore throat |
| 301029011 | Allergic pharyngitis | acute sore throat |
| 106138013 | Atrophic pharyngitis | acute sore throat |
| 348182011 | Chlamydial infection of pharynx | acute sore throat |
| 5037761000006110 | Drainage of parapharyngeal abscess | acute sore throat |
| 805501000006113 | Gonococcal pharynx infection | acute sore throat |
| 3708961000006120 | Gonorrhoea of pharynx | acute sore throat |
| 481673013 | Hypertrophic pharyngitis | acute sore throat |
| 301417011 | Influenza with pharyngitis | acute sore throat |
| 4548341000006110 | Pain in the pharynx | acute sore throat |
| 360534017 | Pharyngeal candidiasis | acute sore throat |
| 4548371000006110 | Pharyngeal pain | acute sore throat |
| 779281000006114 | Pharyngitis | acute sore throat |
| 885011000006118 | Pharyngitis | acute sore throat |
| 301087010 | Pharyngotracheitis | acute sore throat |
| 230901000006114 | Pharynx or nasopharynx cellulitis | acute sore throat |
| 861041000006119 | Retropharyngeal absces.drained | acute sore throat |
| 3334061000006110 | Rhinopharyngitis | acute sore throat |
| 73158014 | Streptococcal pharyngitis | acute sore throat |
| 301086018 | Tracheopharyngitis | acute sore throat |
| 60991000006118 | Vincent's pharyngitis | acute sore throat |
| 2522971000006110 | Viral pharyngitis | acute sore throat |
| 286934013 | Viral pharyngoconjunctivitis | acute sore throat |
| 442131015 | Uvulitis | acute sore throat |
| 301089013 | Pharyngolaryngitis | upper respiratory tract infection |
| 1675541000006110 | Strep pharyngitis probability (Modified Centor) score | acute sore throat |
| 2436021000000110 | Centor criteria score | acute sore throat |
| 50596017 | Swallowing painful | acute sore throat |
| 2986821000006120 | Painful swallowing | acute sore throat |
| 2986831000006120 | Pain on swallowing | acute sore throat |
| 25522012 | Quinsy | acute sore throat |
| 255675016 | O/E - quinsy present | acute sore throat |
| 1493925016 | Drainage of quinsy | acute sore throat |
| 6028751000006120 | Quinsy drained | acute sore throat |
| 853251000006117 | Recurrent tonsillitis | acute sore throat |
| 301047017 | Recurrent acute tonsillitis | acute sore throat |
| 2724571000000110 | FeverPAIN (Fever in last 24 hours, Purulence, Attend rapidly under 3 days, Inflamed tonsils, No cough and/or coryza) Clinical Score | acute sore throat |
| 3382661000006110 | AURTI - Acute upper respiratory tract infection | upper respiratory tract infection |
| 3378371000006120 | URTI - Infection of the upper respiratory tract | upper respiratory tract infection |
| 5655871000006120 | URTI - Viral upper respiratory tract infection | upper respiratory tract infection |
| 362471000006114 | [X]Acute upper respiratory infections | upper respiratory tract infection |
| 460751000006119 | Acute upper respiratory infection | upper respiratory tract infection |
| 406151000006117 | Acute upper respiratory infection of multiple sites | upper respiratory tract infection |
| 3382671000006120 | Acute upper respiratory tract infection | upper respiratory tract infection |
| 6013001000006120 | Bacterial upper respiratory infection | upper respiratory tract infection |
| 13483871000006100 | Infection of upper respiratory tract caused by 2019 novel coronavirus | upper respiratory tract infection |
| 13483881000006100 | Infection of upper respiratory tract caused by Severe acute respiratory syndrome coronavirus 2 | upper respiratory tract infection |
| 301083014 | Other acute upper respiratory infections | upper respiratory tract infection |
| 301090016 | Other upper respiratory infections of multiple sites | upper respiratory tract infection |
| 301088017 | Recurrent upper respiratory tract infection | upper respiratory tract infection |
| 7739191000006110 | Severe acute respiratory syndrome of upper respiratory tract | upper respiratory tract infection |
| 6013011000006120 | Upper respiratory bacterial infection | upper respiratory tract infection |
| 73091000006118 | Upper respiratory infection | upper respiratory tract infection |
| 396089010 | Upper respiratory infection NOS | upper respiratory tract infection |
| 3378381000006120 | Upper respiratory tract infection | upper respiratory tract infection |
| 12990621000006100 | Upper respiratory tract infection caused by 2019-nCoV (novel coronavirus) | upper respiratory tract infection |
| 13012261000006100 | Upper respiratory tract infection caused by SARS-CoV-2 (severe acute respiratory syndrome coronavirus 2) | upper respiratory tract infection |
| 3378391000006110 | URI - Upper respiratory infection | upper respiratory tract infection |
| 350040017 | Viral upper respiratory tract infection | upper respiratory tract infection |
| 136463019 | Common cold | upper respiratory tract infection |
| 195731000006118 | Pyrexial cold | upper respiratory tract infection |
| 598191000006116 | Acute coryza | upper respiratory tract infection |
| 885001000006116 | Coryza | upper respiratory tract infection |
| 681561000006119 | Nasal catarrh - acute | upper respiratory tract infection |
| 3549411000006110 | Nasal catarrh | upper respiratory tract infection |
| 12028011 | Acute laryngitis | upper respiratory tract infection |
| 98755014 | Laryngitis sicca | upper respiratory tract infection |
| 116837013 | Tuberculous laryngitis | upper respiratory tract infection |
| 141055011 | Streptococcal laryngitis | upper respiratory tract infection |
| 301052010 | Acute oedematous laryngitis | upper respiratory tract infection |
| 301053017 | Acute ulcerative laryngitis | upper respiratory tract infection |
| 301054011 | Acute catarrhal laryngitis | upper respiratory tract infection |
| 301055012 | Acute phlegmonous laryngitis | upper respiratory tract infection |
| 301057016 | Acute haemophilus influenzae laryngitis | upper respiratory tract infection |
| 301058014 | Acute pneumococcal laryngitis | upper respiratory tract infection |
| 301059018 | Acute suppurative laryngitis | upper respiratory tract infection |
| 301063013 | Viral laryngitis | upper respiratory tract infection |
| 301064019 | Acute bacterial laryngitis | upper respiratory tract infection |
| 301065018 | Acute laryngitis NOS | upper respiratory tract infection |
| 301082016 | Acute laryngitis and/or tracheitis | upper respiratory tract infection |
| 301416019 | Influenza with laryngitis | upper respiratory tract infection |
| 348220011 | Vincent's laryngitis | upper respiratory tract infection |
| 412576019 | Acute laryngitis and tracheitis | upper respiratory tract infection |
| 412577011 | Acute laryngitis/tracheitis | upper respiratory tract infection |
| 989171000006116 | Acute laryngitis/tracheitis | upper respiratory tract infection |
| 1001611000006110 | Acute laryngitis with obstruction | upper respiratory tract infection |
| 3241481000006110 | Laryngitis | upper respiratory tract infection |
| 5036201000006110 | Acute simple laryngitis | upper respiratory tract infection |
| 5036251000006110 | Acute subglottic laryngitis | upper respiratory tract infection |
| 6016181000006120 | Infective laryngitis | upper respiratory tract infection |
| 7078651000006110 | Laryngitis due to gastro-oesophageal reflux | upper respiratory tract infection |
| 15053015 | Acute tracheitis with obstruction | upper respiratory tract infection |
| 44623012 | Acute tracheitis | upper respiratory tract infection |
| 63301015 | Acute laryngotracheitis without obstruction | upper respiratory tract infection |
| 91647017 | Laryngotracheitis | upper respiratory tract infection |
| 99622016 | Acute laryngotracheitis with obstruction | upper respiratory tract infection |
| 106996011 | Acute tracheitis without obstruction | upper respiratory tract infection |
| 107009017 | Acute laryngotracheitis | upper respiratory tract infection |
| 301068016 | Acute tracheitis NOS | upper respiratory tract infection |
| 301074016 | Acute laryngotracheitis NOS | upper respiratory tract infection |
| 3524071000006110 | Tracheitis | upper respiratory tract infection |
| 3573331000006110 | Viral tracheitis | upper respiratory tract infection |
| 5899261000006110 | Acute viral laryngotracheitis | upper respiratory tract infection |
| 48067018 | Cellulitis of larynx | upper respiratory tract infection |
| 40845016 | Cellulitis of vocal cords | upper respiratory tract infection |
| 8040911000006110 | Cellulitis of nasal mucous membrane | upper respiratory tract infection |
| 25070012 | Parotitis | upper respiratory tract infection |
| 44466012 | Toxic parotitis | upper respiratory tract infection |
| 80417018 | Allergic parotitis | upper respiratory tract infection |
| 302196010 | Parotitis NOS | upper respiratory tract infection |
| 360291011 | Mumps parotitis | upper respiratory tract infection |
| 1223202016 | Infectious parotitis | upper respiratory tract infection |
| 1223203014 | Epidemic parotitis | upper respiratory tract infection |
| 244501000006113 | Parotitis - epidemic | upper respiratory tract infection |
| 2214191000000110 | Infective parotitis | upper respiratory tract infection |
| 5075531000006110 | Recurrent parotitis | upper respiratory tract infection |
| 5560191000006110 | Parotitis - non-mumps | upper respiratory tract infection |
| 301144014 | Other specified acute respiratory infections | upper respiratory tract infection |
| 1467013 | Acute epiglottitis with obstruction | upper respiratory tract infection |
| 83125011 | Acute epiglottitis without obstruction | upper respiratory tract infection |
| 117891019 | Viral epiglottitis | upper respiratory tract infection |
| 301080012 | Acute epiglottitis | upper respiratory tract infection |
| 456591000006112 | Acute epiglottitis (non-streptococcal) | upper respiratory tract infection |
| 3807341000006120 | Epiglottitis | upper respiratory tract infection |
| 7654141000006120 | Supraglottitis | upper respiratory tract infection |
| 1001621000006120 | Croup | upper respiratory tract infection |
| 3657821000006120 | Croup syndrome | upper respiratory tract infection |
| 5036271000006120 | Recurrent allergic croup | upper respiratory tract infection |
| 5259301000006120 | Croupy breathing | upper respiratory tract infection |
| 411486018 | Nasal infection | upper respiratory tract infection |
| 142044014 | Tuberculosis of tracheobronchial lymph nodes | upper respiratory tract infection |
| 286232019 | Isolated tracheobronchial tuberculosis | upper respiratory tract infection |
| 39820013 | Nasal vestibulitis | upper respiratory tract infection |
| 397999013 | Nose running | upper respiratory tract infection |
| 961831000006111 | Rhinorrhea/nasal congestion | upper respiratory tract infection |
| 5532911000006110 | Complaining of nasal congestion | upper respiratory tract infection |
| 255579018 | O/E - rhinorrhoea | upper respiratory tract infection |
| 2161421018 | Rhinorrhoea | upper respiratory tract infection |
| 3549401000006110 | Rhinorrhea | upper respiratory tract infection |
| 3732431000006110 | Posterior rhinorrhoea | upper respiratory tract infection |
| 5596441000006120 | Anterior rhinorrhoea | upper respiratory tract infection |
| 5596451000006120 | Anterior rhinorrhea | upper respiratory tract infection |
| 255576013 | O/E - nasal discharge | upper respiratory tract infection |
| 255580015 | O/E-nasal discharge-foul smell | upper respiratory tract infection |
| 255581016 | On examination - nasal discharge - mucopurulent | upper respiratory tract infection |
| 255587017 | O/E - nasal discharge NOS | upper respiratory tract infection |
| 397998017 | Nasal discharge present | upper respiratory tract infection |
| 2726611000006110 | Purulent nasal discharge | upper respiratory tract infection |
| 3549381000006110 | Nasal discharge | upper respiratory tract infection |
| 4576251000006120 | On examination - nasal discharge | upper respiratory tract infection |
| 1221348018 | O/E - nose discharge | upper respiratory tract infection |
| 216652015 | Feverish cold | upper respiratory tract infection |
| 406124015 | Neonatal snuffles | upper respiratory tract infection |
| 1777356018 | Snuffles | upper respiratory tract infection |
| 5523971000006120 | Snuffles in newborn | upper respiratory tract infection |
| 459357017 | Suspected UTI (urinary tract infection) | acute lower urinary tract infection |
| 2732311000000110 | Uncomplicated UTI (urinary tract infection) | acute lower urinary tract infection |
| 5888041000006110 | Proteus UTI (urinary tract infection) | acute lower urinary tract infection |
| 6013071000006110 | Bacterial UTI (urinary tract infection) | acute lower urinary tract infection |
| 7135041000006120 | Acute UTI (urinary tract infection) | acute lower urinary tract infection |
| 4790241000006110 | Recurrent UTI - urinary tract infection | acute lower urinary tract infection |
| 21171016 | Neonatal urinary tract infection | acute lower urinary tract infection |
| 113884018 | Urinary tract infection | acute lower urinary tract infection |
| 304319014 | Recurrent urinary tract infection | acute lower urinary tract infection |
| 304323018 | Urinary tract infection, site not specified NOS | acute lower urinary tract infection |
| 305603019 | Urinary tract infection following abortive pregnancy | acute lower urinary tract infection |
| 305977017 | Urinary tract infection following delivery | acute lower urinary tract infection |
| 308102012 | Urinary tract infection in pregnancy | acute lower urinary tract infection |
| 450823012 | UTI - urinary tract infection in pregnancy | acute lower urinary tract infection |
| 1490641019 | Urinary tract infection complicating pregnancy | acute lower urinary tract infection |
| 74741000006111 | Urinary Tract Infection | acute lower urinary tract infection |
| 74781000006117 | Urinary tract infection, site not specified | acute lower urinary tract infection |
| 183141000006112 | Recurrent urinary tract infection | acute lower urinary tract infection |
| 183161000006111 | Recurrent urinary tract infections | acute lower urinary tract infection |
| 183171000006116 | Recurrent urinary tract infection | acute lower urinary tract infection |
| 216611000006114 | Postoperative urinary tract infection | acute lower urinary tract infection |
| 886811000006115 | Urinary tract infection NOS | acute lower urinary tract infection |
| 1761271000006120 | [SHHAPT] Urinary tract infection | acute lower urinary tract infection |
| 2247821000000120 | On urinary tract infection care pathway | acute lower urinary tract infection |
| 2277891000000120 | Catheter-associated urinary tract infection | acute lower urinary tract infection |
| 2277901000000120 | CAUTI - catheter-associated urinary tract infection | acute lower urinary tract infection |
| 2563091000006110 | Lower urinary tract infection | acute lower urinary tract infection |
| 2563101000006110 | UTI - Lower urinary tract infection | acute lower urinary tract infection |
| 2732301000000110 | Uncomplicated urinary tract infection | acute lower urinary tract infection |
| 3615701000006120 | UTI - Urinary tract infection | acute lower urinary tract infection |
| 5888011000006110 | Coliform urinary tract infection | acute lower urinary tract infection |
| 5888021000006120 | Escherichia coli urinary tract infection | acute lower urinary tract infection |
| 5888031000006110 | Proteus urinary tract infection | acute lower urinary tract infection |
| 5888051000006110 | Pseudomonas urinary tract infection | acute lower urinary tract infection |
| 6051761000006110 | Suspected urinary tract infection | acute lower urinary tract infection |
| 7135021000006110 | Acute urinary tract infection | acute lower urinary tract infection |
| 7144721000006110 | Acute lower urinary tract infection | acute lower urinary tract infection |
| 7534301000006110 | Urinary tract infection associated with catheter | acute lower urinary tract infection |
| 8031341000006120 | Febrile urinary tract infection | acute lower urinary tract infection |
| 8120621000006110 | Urinary tract infection caused by Enterococcus | acute lower urinary tract infection |
| 8120631000006120 | Urinary tract infection caused by Klebsiella | acute lower urinary tract infection |
| 14504521000006100 | Urinary tract infection caused by Escherichia coli | acute lower urinary tract infection |
| 8285011 | Subacute cystitis | acute lower urinary tract infection |
| 65119018 | Cystitis | acute lower urinary tract infection |
| 113331011 | Acute cystitis | acute lower urinary tract infection |
| 107972012 | Follicular cystitis | acute lower urinary tract infection |
| 304205011 | Recurrent cystitis | acute lower urinary tract infection |
| 304209017 | Cystitis NOS | acute lower urinary tract infection |
| 411392011 | Cystitis of pregnancy | acute lower urinary tract infection |
| 3344841000006110 | Emphysematous cystitis | acute lower urinary tract infection |
| 3376381000006120 | Echinococcal cystitis | acute lower urinary tract infection |
| 3925321000006120 | Hemorrhagic cystitis | acute lower urinary tract infection |
| 5096711000006110 | Infective cystitis | acute lower urinary tract infection |
| 5096751000006110 | Acute cystitis - culture-negative | acute lower urinary tract infection |
| 5096761000006110 | Recurrent cystitis - culture-negative | acute lower urinary tract infection |
| 5097681000006110 | Acute culture positive cystitis | acute lower urinary tract infection |
| 5097691000006110 | Recurrent infective cystitis | acute lower urinary tract infection |
| 5560201000006120 | Acute recurrent cystitis | acute lower urinary tract infection |
| 5968141000006120 | Acute infective cystitis | acute lower urinary tract infection |
| 7028541000006110 | Bacterial cystitis | acute lower urinary tract infection |
| 3124871000006110 | Bladder infection | acute lower urinary tract infection |
| 306087011 | Infections of bladder in pregnancy | acute lower urinary tract infection |
| 2580881000006110 | Actinomycotic cystitis | acute lower urinary tract infection |
| 1230922019 | Cystitis in actinomycosis | acute lower urinary tract infection |
| 221191000000117 | Cystitis in bilharziasis | acute lower urinary tract infection |
| 304192013 | Cystitis in diseases EC | acute lower urinary tract infection |
| 1217047011 | Cystitis in amoebiasis | acute lower urinary tract infection |
| 1231277013 | Cystitis in echinococcus infestation | acute lower urinary tract infection |
| 304198012 | Cystitis in moniliasis | acute lower urinary tract infection |
| 304201019 | Cystitis in trichomoniasis | acute lower urinary tract infection |
| 606691000006115 | Cystitis in tuberculosis | acute lower urinary tract infection |
| 4789821000006110 | Monilial cystitis | acute lower urinary tract infection |
| 304208013 | Other cystitis NOS | acute lower urinary tract infection |
| 304206012 | Other specified cystitis | acute lower urinary tract infection |
| 3019561000006120 | TB - Tuberculous cystitis | acute lower urinary tract infection |
| 260296019 | Urine culture - Pseudomonas | acute lower urinary tract infection |
| 260297011 | Urine culture - Bacteria OS | acute lower urinary tract infection |
| 75451000006118 | Urine culture - acid-fast bacilli | acute lower urinary tract infection |
| 75491000006112 | Urine culture - Escherich.coli | acute lower urinary tract infection |
| 75551000006117 | Urine culture - Streptococcus faecalis | acute lower urinary tract infection |
| 260290013 | Urine culture - mixed growth | acute lower urinary tract infection |
| 260291012 | Urine culture - E. coli | acute lower urinary tract infection |
| 260292017 | Urine culture - Proteus | acute lower urinary tract infection |
| 260295015 | Urine culture - Staph. albus | acute lower urinary tract infection |
| 124691012 | Malakoplakia of bladder | acute lower urinary tract infection |
| 6013061000006120 | Bacterial urinary infection | acute lower urinary tract infection |
| 98546013 | Life threatening acute exacerbation of intrinsic asthma | asthma exacerbation |
| 151338014 | Life threatening acute exacerbation of allergic asthma | asthma exacerbation |
| 264556011 | Asthma prophylactic medication used | asthma exacerbation |
| 283550015 | Emergency hospital admission for asthma | asthma exacerbation |
| 350151019 | Acute exacerbation of allergic asthma | asthma exacerbation |
| 350156012 | Acute exacerbation of intrinsic asthma | asthma exacerbation |
| 396118011 | Life threatening acute exacerbation of asthma | asthma exacerbation |
| 396119015 | Asthma attack | asthma exacerbation |
| 409865018 | Asthma attack NOS | asthma exacerbation |
| 419211018 | Acute exacerbation of asthma | asthma exacerbation |
| 3511374015 | Moderate acute exacerbation of asthma | asthma exacerbation |
| 3514925011 | Acute severe exacerbation of asthma co-occurrent and due to allergic asthma | asthma exacerbation |
| 3637387011 | Exacerbation of allergic asthma | asthma exacerbation |
| 145961000006117 | Acute severe exacerbation of asthma | asthma exacerbation |
| 2010031000006120 | Acute infective exacerbation of asthma | asthma exacerbation |
| 2010041000006110 | Acute non-infective exacerbation of asthma | asthma exacerbation |
| 7052221000006120 | Exacerbation of intermittent asthma | asthma exacerbation |
| 7258581000006110 | Acute exacerbation of chronic asthmatic bronchitis | asthma exacerbation |
| 7617611000006110 | Exacerbation of mild persistent asthma | asthma exacerbation |
| 7617621000006120 | Exacerbation of moderate persistent asthma | asthma exacerbation |
| 7617631000006120 | Exacerbation of severe persistent asthma | asthma exacerbation |
| 7626521000006110 | Acute severe exacerbation of severe persistent asthma | asthma exacerbation |
| 7626531000006110 | Acute severe exacerbation of moderate persistent asthma | asthma exacerbation |
| 7626541000006110 | Acute severe exacerbation of mild persistent asthma | asthma exacerbation |
| 7628301000006110 | Acute severe exacerbation of allergic asthma | asthma exacerbation |
| 7628321000006120 | Acute severe exacerbation of immunoglobin E-mediated allergic asthma | asthma exacerbation |
| 7628331000006110 | Acute severe exacerbation of intrinsic asthma | asthma exacerbation |
| 7961271000006110 | Acute severe refractory exacerbation of asthma | asthma exacerbation |
| 7961281000006110 | Acute severe asthma | asthma exacerbation |
| 7970171000006120 | Acute exacerbation of chronic obstructive airways disease with asthma | asthma exacerbation |
| 8031201000006110 | Acute exacerbation of asthma co-occurrent with allergic rhinitis | asthma exacerbation |
| 8042571000006110 | Acute exacerbation of moderate persistent asthma | asthma exacerbation |
| 8042581000006110 | Acute exacerbation of mild persistent asthma | asthma exacerbation |
| 9315391000006120 | Acute severe exacerbation of moderate persistent asthma co-occurrent with allergic rhinitis | asthma exacerbation |
| 9317311000006120 | Chronic obstructive asthma co-occurrent with acute exacerbation of asthma | asthma exacerbation |
| 11922851000006100 | Acute asthma | asthma exacerbation |
| 13619851000006100 | Acute severe exacerbation of allergic asthma | asthma exacerbation |
| 13619961000006100 | Exacerbation of allergic asthma due to infection | asthma exacerbation |
| 14504061000006100 | Acute asthma | asthma exacerbation |
| 14504071000006100 | Asthma attack | asthma exacerbation |
| 8287171000006110 | Acute non-infective exacerbation of COPD (chronic obstructive pulmonary disease) | COPD exacerbaion |
| 4781421000006110 | Acute exacerbation of COPD | COPD exacerbaion |
| 4781431000006110 | Acute exacerbation of chronic obstructive pulmonary disease | COPD exacerbaion |
| 2010061000006110 | Acute non-infective exacerbation of chronic obstructive pulmonary disease | COPD exacerbaion |
| 553211000006119 | Chron obstruct pulmonary dis wth acute exacerbation, unspec | COPD exacerbaion |
| 424365019 | Acute infective exacerbation of chronic obstructive airways disease | COPD exacerbaion |
| 301453013 | Acute exacerbation of chronic obstructive airways disease | COPD exacerbaion |
| 80425016 | Impetigo | impetigo |
| 298959013 | Infective otitis externa due to impetigo | impetigo |
| 298960015 | Impetigo - otitis externa | impetigo |
| 308382017 | Impetigo contagiosa unspecified | impetigo |
| 308385015 | Impetigo contagiosa gyrata | impetigo |
| 308386019 | Circinate impetigo | impetigo |
| 308387011 | Impetigo neonatorum | impetigo |
| 308388018 | Impetigo simplex | impetigo |
| 308389014 | Impetigo follicularis | impetigo |
| 308390017 | Chronic symmetrical impetigo | impetigo |
| 308391018 | Impetigo NOS | impetigo |
| 308748013 | Impetigo herpetiformis | impetigo |
| 1786733013 | Bullous impetigo | impetigo |
| 3365991000006120 | Bockhart impetigo | impetigo |
| 5027641000006110 | Impetigo of eyelid | impetigo |
| 5120891000006110 | Non-bullous impetigo | impetigo |
| 5120911000006110 | Streptococcal impetigo | impetigo |
| 5120921000006120 | Staphylococcal non-bullous impetigo | impetigo |
| 5502121000006120 | Follicular impetigo | impetigo |
| 6617371000006110 | Bullous staphylococcal impetigo | impetigo |
| 6618681000006120 | Impetigo bullosa | impetigo |
| 907511000006114 | [RFC] Chest infection | lower respiratory tract infection |
| 907541000006113 | [RFC] Whooping cough | lower respiratory tract infection |
| 301821013 | [X]Acute bronchiolitis due to other specified organisms | lower respiratory tract infection |
| 316899011 | [X]Congenital pneumonia due to other organisms | lower respiratory tract infection |
| 301819015 | [X]Other acute lower respiratory infections | lower respiratory tract infection |
| 301811017 | [X]Other bacterial pneumonia | lower respiratory tract infection |
| 301818011 | [X]Other pneumonia, organism unspecified | lower respiratory tract infection |
| 288238011 | [X]Other pulmonary aspergillosis | lower respiratory tract infection |
| 301809014 | [X]Other viral pneumonia | lower respiratory tract infection |
| 301810016 | [X]Pneumonia due to other aerobic gram-negative bacteria | lower respiratory tract infection |
| 301812012 | [X]Pneumonia due to other specified infectious organisms | lower respiratory tract infection |
| 301813019 | [X]Pneumonia in bacterial diseases classified elsewhere | lower respiratory tract infection |
| 301815014 | [X]Pneumonia in mycoses classified elsewhere | lower respiratory tract infection |
| 301816010 | [X]Pneumonia in parasitic diseases classified elsewhere | lower respiratory tract infection |
| 301814013 | [X]Pneumonia in viral diseases classified elsewhere | lower respiratory tract infection |
| 288237018 | [X]Pulmonary histoplasmosis capsulati, unspecified | lower respiratory tract infection |
| 288040011 | [X]Whooping cough, unspecified | lower respiratory tract infection |
| 301686019 | Abscess of lung with pneumonia | lower respiratory tract infection |
| 4781261000006120 | Actinomycotic pneumonia | lower respiratory tract infection |
| 8015301000006110 | Acute aspiration pneumonia | lower respiratory tract infection |
| 10187013 | Acute bronchiolitis | lower respiratory tract infection |
| 5053381000006120 | Acute bronchiolitis due to adenovirus | lower respiratory tract infection |
| 1816261000006120 | Acute bronchiolitis due to human metapneumovirus | lower respiratory tract infection |
| 301129013 | Acute bronchiolitis due to other specified organisms | lower respiratory tract infection |
| 301128017 | Acute bronchiolitis due to respiratory syncytial virus | lower respiratory tract infection |
| 301130015 | Acute bronchiolitis NOS | lower respiratory tract infection |
| 25801011 | Acute bronchiolitis with bronchospasm | lower respiratory tract infection |
| 4353381000006110 | Acute bronchopneumonia | lower respiratory tract infection |
| 2478755014 | Acute capillary bronchiolitis | lower respiratory tract infection |
| 301647018 | Acute dry pleurisy | lower respiratory tract infection |
| 3556351000006110 | Acute eosinophilic pneumonia | lower respiratory tract infection |
| 7307141000006110 | Acute exacerbation of bronchiectasis | lower respiratory tract infection |
| 7049011000006110 | Acute exacerbation of chronic bronchitis | lower respiratory tract infection |
| 8090191000006110 | Acute exacerbation of chronic obstructive bronchitis | lower respiratory tract infection |
| 301126018 | Acute exudative bronchiolitis | lower respiratory tract infection |
| 579878017 | Acute lower respiratory tract infection | lower respiratory tract infection |
| 457801000006117 | Acute lower respiratory tract infection | lower respiratory tract infection |
| 546411000006111 | Acute lower respiratory tract infection | lower respiratory tract infection |
| 99508014 | Acute obliterating bronchiolitis | lower respiratory tract infection |
| 287652018 | Acute pulmonary blastomycosis | lower respiratory tract infection |
| 287596012 | Acute pulmonary coccidioidomycosis | lower respiratory tract infection |
| 301145010 | Acute respiratory infection | lower respiratory tract infection |
| 300997012 | Acute respiratory infections | lower respiratory tract infection |
| 5053371000006120 | Acute viral bronchiolitis | lower respiratory tract infection |
| 3163841000006110 | Adenoviral pneumonia | lower respiratory tract infection |
| 6717641000006120 | Allergic bronchitis | lower respiratory tract infection |
| 477291000006114 | Allergic bronchitis NEC | lower respiratory tract infection |
| 63349014 | Allergic bronchopulmonary aspergillosis | lower respiratory tract infection |
| 1709201000006110 | Allergic bronchopulmonary aspergillosis | lower respiratory tract infection |
| 4780011000006110 | ARI - Acute respiratory infections | lower respiratory tract infection |
| 885391000006114 | Aspiration pneumonia | lower respiratory tract infection |
| 1756121000006110 | Aspiration pneumonia | lower respiratory tract infection |
| 3177041000006120 | Aspiration pneumonia caused by inhalation of vomitus | lower respiratory tract infection |
| 325070014 | Aspiration pneumonia resulting from a procedure | lower respiratory tract infection |
| 350051017 | Atypical pneumonia | lower respiratory tract infection |
| 6013021000006110 | Bacterial lower respiratory infection | lower respiratory tract infection |
| 3690461000006120 | Bacterial pleurisy | lower respiratory tract infection |
| 141595017 | Bacterial pleurisy with effusion | lower respiratory tract infection |
| 301661011 | Bacterial pleurisy with effusion NOS | lower respiratory tract infection |
| 301376012 | Bacterial pneumonia | lower respiratory tract infection |
| 6012991000006110 | Bacterial respiratory infection | lower respiratory tract infection |
| 301650015 | Basal pleurisy | lower respiratory tract infection |
| 451130014 | Basal pneumonia | lower respiratory tract infection |
| 1772711000006120 | Basal pneumonia | lower respiratory tract infection |
| 1772721000006110 | Bilateral basal pneumonia | lower respiratory tract infection |
| 6566941000006110 | Bilateral bronchopneumonia | lower respiratory tract infection |
| 7052581000006120 | Bilateral lower lobe pneumonia | lower respiratory tract infection |
| 932231000006112 | Bilateral pneumonia | lower respiratory tract infection |
| 6750331000006110 | Bilateral pneumonia | lower respiratory tract infection |
| 5055881000006110 | Bronchial infection | lower respiratory tract infection |
| 6566921000006120 | Bronchial pneumonia | lower respiratory tract infection |
| 2564641000006110 | Bronchiolitis | lower respiratory tract infection |
| 7302821000006110 | Bronchiolitis due to Human metapneumovirus | lower respiratory tract infection |
| 492443010 | Bronchiolitis obliterans | lower respiratory tract infection |
| 474828010 | Bronchiolitis obliterans organising pneumonia | lower respiratory tract infection |
| 4426671000006110 | Bronchiolitis with interstitial lung disease | lower respiratory tract infection |
| 13651601000006100 | Bronchitis co-occurrent with acute wheeze | lower respiratory tract infection |
| 396105015 | Bronchopneumonia | lower respiratory tract infection |
| 885231000006119 | Bronchopneumonia | lower respiratory tract infection |
| 1772771000006110 | Bronchopneumonia | lower respiratory tract infection |
| 9309521000006110 | Bronchopneumonia due to virus | lower respiratory tract infection |
| 2595971000006110 | Bronchopulmonary aspergillosis | lower respiratory tract infection |
| 219761000006111 | Candidal pneumonia | lower respiratory tract infection |
| 5153171000006110 | Capillaria aerophila chest infection | lower respiratory tract infection |
| 1576341000006120 | Cause of Death- Bronchopneumonia | lower respiratory tract infection |
| 7475011000006120 | Cavitary pneumonia | lower respiratory tract infection |
| 301143015 | Chest cold | lower respiratory tract infection |
| 546421000006115 | Chest infection - influenza with pneumonia | lower respiratory tract infection |
| 1222332017 | Chest infection - other bacterial pneumonia | lower respiratory tract infection |
| 546441000006110 | Chest infection - pnemonia due to unspecified organism | lower respiratory tract infection |
| 546451000006112 | Chest infection - pneumococcal pneumonia | lower respiratory tract infection |
| 1222333010 | Chest infection - pneumonia organism OS | lower respiratory tract infection |
| 546481000006116 | Chest infection - unspecified bronchopneumonia | lower respiratory tract infection |
| 546491000006118 | Chest infection - viral pneumonia | lower respiratory tract infection |
| 546511000006112 | Chest infection with infectious disease EC | lower respiratory tract infection |
| 220251000006118 | Chickenpox pneumonia | lower respiratory tract infection |
| 350054013 | Chlamydial pneumonia | lower respiratory tract infection |
| 8033221000006120 | Chronic obstructive lung disease co-occurrent with acute bronchitis | lower respiratory tract infection |
| 555461000006119 | Chronic obstructive pulmonary disease with acute lower respiratory infection | lower respiratory tract infection |
| 8031971000006110 | Chronic pneumonia | lower respiratory tract infection |
| 1479355018 | Community acquired pneumonia | lower respiratory tract infection |
| 1787121000006120 | Community acquired pneumonia | lower respiratory tract infection |
| 316898015 | Congenital bacterial pneumonia | lower respiratory tract infection |
| 316320014 | Congenital pneumonia due to pseudomonas | lower respiratory tract infection |
| 316324017 | Congenital pneumonia NOS | lower respiratory tract infection |
| 5589281000006110 | Congenital viral pneumonia | lower respiratory tract infection |
| 2653731000006110 | Consolidation | lower respiratory tract infection |
| 13948281000006100 | Consolidation of lung present | lower respiratory tract infection |
| 13486761000006100 | COVID-19 lower respiratory infection | lower respiratory tract infection |
| 13484091000006100 | COVID-19 pneumonia | lower respiratory tract infection |
| 474819014 | Cryptogenic organising pneumonia | lower respiratory tract infection |
| 2622071000006120 | Cytomegaloviral pneumonia | lower respiratory tract infection |
| 1234028011 | Cytomegaloviral pneumonitis | lower respiratory tract infection |
| 22395010 | Diaphragmatic pleurisy | lower respiratory tract infection |
| 5873881000006110 | Drainage of empyema | lower respiratory tract infection |
| 5053821000006110 | Dry pleurisy | lower respiratory tract infection |
| 633531000006116 | E.coli pneumonia | lower respiratory tract infection |
| 285100019 | Emphysematous bronchitis | lower respiratory tract infection |
| 456436010 | Empyema | lower respiratory tract infection |
| 640711000006112 | Empyema NOS | lower respiratory tract infection |
| 301635012 | Empyema of pleura | lower respiratory tract infection |
| 50946012 | Empyema with bronchocutaneous fistula | lower respiratory tract infection |
| 47025013 | Empyema with bronchopleural fistula | lower respiratory tract infection |
| 406305010 | Empyema with fistula | lower respiratory tract infection |
| 640771000006115 | Empyema with fistula NOS | lower respiratory tract infection |
| 57249012 | Empyema with hepatopleural fistula | lower respiratory tract infection |
| 84488013 | Empyema with mediastinal fistula | lower respiratory tract infection |
| 640801000006118 | Empyema with no fistula | lower respiratory tract infection |
| 640811000006115 | Empyema with no fistula NOS | lower respiratory tract infection |
| 301625018 | Empyema with pleural fistula NOS | lower respiratory tract infection |
| 301626017 | Empyema with thoracic fistula NOS | lower respiratory tract infection |
| 46467017 | Encysted pleurisy | lower respiratory tract infection |
| 6298461000006110 | Eosinophilic pneumonia | lower respiratory tract infection |
| 2671161016 | Escherichia coli pneumonia | lower respiratory tract infection |
| 14133001000006100 | Exacerbation of bronchiectasis caused by infection | lower respiratory tract infection |
| 301669013 | Exudative pleurisy NOS | lower respiratory tract infection |
| 128069014 | Fibrinous pleurisy | lower respiratory tract infection |
| 6438611000006120 | Follicular bronchiolitis | lower respiratory tract infection |
| 3536621000006110 | Fungal infection of lung | lower respiratory tract infection |
| 5053491000006110 | Fungal pneumonia | lower respiratory tract infection |
| 6013321000006110 | Fungal respiratory infection | lower respiratory tract infection |
| 12652015 | Gangrenous pneumonia | lower respiratory tract infection |
| 4781141000006110 | Group B streptococcal pneumonia | lower respiratory tract infection |
| 219841000006117 | Haemophilus influenzae pneumonia | lower respiratory tract infection |
| 1814961000006120 | Hantavirus cardio-pulmonary syndrome | lower respiratory tract infection |
| 179040014 | Hantavirus pulmonary syndrome | lower respiratory tract infection |
| 7044731000006110 | HAP - hospital acquired pneumonia | lower respiratory tract infection |
| 6765531000006120 | Healthcare associated pneumonia | lower respiratory tract infection |
| 350073012 | Herpes simplex pneumonia | lower respiratory tract infection |
| 287618016 | Histoplasma capsulatum with pneumonia | lower respiratory tract infection |
| 287630010 | Histoplasma duboisii with pneumonia | lower respiratory tract infection |
| 287639011 | Histoplasmosis with pneumonia | lower respiratory tract infection |
| 1814921000006110 | HIV disease resulting in Pneumocystis jirovecii pneumonia | lower respiratory tract infection |
| 2674072012 | Hospital acquired pneumonia | lower respiratory tract infection |
| 1787131000006120 | Hospital acquired pneumonia | lower respiratory tract infection |
| 5573811000006120 | Infection of lower respiratory tract and mediastinum | lower respiratory tract infection |
| 5053611000006120 | Infectious mononucleosis pneumonia | lower respiratory tract infection |
| 6041771000006120 | Infective pleurisy | lower respiratory tract infection |
| 301408011 | Infective pneumonia | lower respiratory tract infection |
| 3669971000006110 | Infective pneumonia acquired prenatally | lower respiratory tract infection |
| 885251000006114 | Influenza + pneumonia | lower respiratory tract infection |
| 1229740013 | Influenza with bronchopneumonia | lower respiratory tract infection |
| 778871000006116 | Influenza with pneumonia | lower respiratory tract infection |
| 2609031000000110 | Influenza with pneumonia due to seasonal influenza virus | lower respiratory tract infection |
| 301414016 | Influenza with pneumonia NOS | lower respiratory tract infection |
| 301413010 | Influenza with pneumonia, influenza virus identified | lower respiratory tract infection |
| 3164871000006120 | Influenzal bronchopneumonia | lower respiratory tract infection |
| 6421012 | Invasive pulmonary aspergillosis | lower respiratory tract infection |
| 5887701000006110 | Left basal pneumonia | lower respiratory tract infection |
| 5887721000006120 | Left lower lobe pneumonia | lower respiratory tract infection |
| 1772731000006110 | Left lower zone pneumonia | lower respiratory tract infection |
| 5887821000006110 | Left upper lobe pneumonia | lower respiratory tract infection |
| 1772691000006120 | Left upper zone pneumonia | lower respiratory tract infection |
| 219771000006116 | Legionella pneumonia | lower respiratory tract infection |
| 1772701000006120 | Lingular pneumonia | lower respiratory tract infection |
| 6899331000006110 | Lipoid pneumonia | lower respiratory tract infection |
| 742901000006119 | Lipoid pneumonia (exogenous) | lower respiratory tract infection |
| 5887711000006110 | LLL - Left lower lobe pneumonia | lower respiratory tract infection |
| 5887731000006120 | LLZ - Left lower zone pneumonia | lower respiratory tract infection |
| 885191000006112 | Lobar -pneumococcal -pneumonia | lower respiratory tract infection |
| 301409015 | Lobar pneumonia | lower respiratory tract infection |
| 1772681000006120 | Lobar pneumonia | lower respiratory tract infection |
| 5887841000006120 | Lobar pneumonia left upper lobe | lower respiratory tract infection |
| 5887931000006120 | Lobar pneumonia right upper lobe | lower respiratory tract infection |
| 6566931000006120 | Lobular pneumonia | lower respiratory tract infection |
| 4782121000006110 | Loculated empyema | lower respiratory tract infection |
| 5887681000006110 | Lower lobe pneumonia | lower respiratory tract infection |
| 733471000006110 | Lower resp tract infection | lower respiratory tract infection |
| 3316381000006110 | Lower respiratory infection | lower respiratory tract infection |
| 13486741000006100 | Lower respiratory infection caused by SARS-CoV-2 (severe acute respiratory syndrome coronavirus 2) | lower respiratory tract infection |
| 13486771000006100 | Lower respiratory infection caused by Severe acute respiratory syndrome coronavirus 2 | lower respiratory tract infection |
| 396090018 | Lower respiratory tract infection | lower respiratory tract infection |
| 3316401000006110 | LRTI - Lower respiratory tract infection | lower respiratory tract infection |
| 512123013 | Lung consolidation | lower respiratory tract infection |
| 4411771000006110 | Lung infection | lower respiratory tract infection |
| 301386013 | Measles pneumonia | lower respiratory tract infection |
| 78272011 | Mycoplasma pneumonia | lower respiratory tract infection |
| 3259121000006120 | Mycoplasma pneumoniae pneumonia | lower respiratory tract infection |
| 2164029012 | Mycoplasmal pneumonia | lower respiratory tract infection |
| 2612661000006110 | Necrotising pneumonia | lower respiratory tract infection |
| 5589311000006120 | Neonatal aspiration pneumonia | lower respiratory tract infection |
| 5053461000006120 | Neonatal chlamydial pneumonia | lower respiratory tract infection |
| 5053571000006110 | Neonatal pneumonia | lower respiratory tract infection |
| 4781271000006110 | Nocardial pneumonia | lower respiratory tract infection |
| 4125731000006110 | Non-infectious pneumonia | lower respiratory tract infection |
| 5605771000006120 | Non-tuberculous mycobacterial pneumonia | lower respiratory tract infection |
| 7044721000006120 | Nosocomial pneumonia | lower respiratory tract infection |
| 253996010 | O/E - consolidation | lower respiratory tract infection |
| 253995014 | O/E - consolidation present | lower respiratory tract infection |
| 301127010 | Obliterating fibrous bronchiolitis | lower respiratory tract infection |
| 3145221000006110 | Obliterative bronchiolitis | lower respiratory tract infection |
| 3613101000006110 | Organised pneumonia | lower respiratory tract infection |
| 134668017 | Ornithosis with pneumonia | lower respiratory tract infection |
| 301368010 | Other bacterial pneumonia | lower respiratory tract infection |
| 316323011 | Other specified congenital pneumonia | lower respiratory tract infection |
| 301430012 | Other specified pneumonia or influenza | lower respiratory tract infection |
| 286221019 | Other specified pulmonary tuberculosis | lower respiratory tract infection |
| 286451010 | Other whooping cough NOS | lower respiratory tract infection |
| 3555901000006110 | Parainfluenza pneumonia | lower respiratory tract infection |
| 3555881000006120 | Parainfluenzal pneumonia | lower respiratory tract infection |
| 6870511000006110 | PCP - Pneumocystis pneumonia | lower respiratory tract infection |
| 3818781000006110 | Pleural effusion associated with pulmonary infection | lower respiratory tract infection |
| 8033171000006110 | Pleural effusion caused by bacterial infection | lower respiratory tract infection |
| 218821000006118 | Pleural empyema | lower respiratory tract infection |
| 5525211000006120 | Pleural empyema with fistula | lower respiratory tract infection |
| 301644013 | Pleurisy | lower respiratory tract infection |
| 301646010 | Pleurisy without effusion or active tuberculosis | lower respiratory tract infection |
| 301656014 | Pleurisy without effusion or active tuberculosis NOS | lower respiratory tract infection |
| 739941000006111 | Pneumococcal lobar pneumonia | lower respiratory tract infection |
| 5407016 | Pneumococcal pleurisy | lower respiratory tract infection |
| 40785018 | Pneumococcal pleurisy with effusion | lower respiratory tract infection |
| 5053421000006110 | Pneumococcal pneumonia | lower respiratory tract infection |
| 220181000006118 | Pneumocystosis pneumonia | lower respiratory tract infection |
| 301817018 | Pneumonia | lower respiratory tract infection |
| 301357010 | Pneumonia and influenza | lower respiratory tract infection |
| 13484101000006100 | Pneumonia caused by 2019 novel coronavirus | lower respiratory tract infection |
| 12990651000006100 | Pneumonia caused by 2019-nCoV (novel coronavirus) | lower respiratory tract infection |
| 13484121000006100 | Pneumonia caused by 2019-nCoV (novel coronavirus) | lower respiratory tract infection |
| 7699541000006120 | Pneumonia caused by Human coronavirus | lower respiratory tract infection |
| 13012271000006100 | Pneumonia caused by SARS-CoV-2 (severe acute respiratory syndrome coronavirus 2) | lower respiratory tract infection |
| 13484081000006100 | Pneumonia caused by SARS-CoV-2 (severe acute respiratory syndrome coronavirus 2) | lower respiratory tract infection |
| 219801000006119 | Pneumonia due to adenovirus | lower respiratory tract infection |
| 301375011 | Pneumonia due to bacteria NOS | lower respiratory tract infection |
| 219821000006112 | Pneumonia due to Eaton's agent | lower respiratory tract infection |
| 3334901000006110 | Pneumonia due to Escherichia coli | lower respiratory tract infection |
| 2765492018 | Pneumonia due to Gram negative bacteria | lower respiratory tract infection |
| 219851000006115 | Pneumonia due to haemophilus influenzae | lower respiratory tract infection |
| 2872480010 | Pneumonia due to Human metapneumovirus | lower respiratory tract infection |
| 107173015 | Pneumonia due to Klebsiella pneumoniae | lower respiratory tract infection |
| 219881000006111 | Pneumonia due to other aerobic gram-negative bacteria | lower respiratory tract infection |
| 301370018 | Pneumonia due to other specified bacteria | lower respiratory tract infection |
| 301377015 | Pneumonia due to other specified organisms | lower respiratory tract infection |
| 1232627018 | Pneumonia due to parainfluenza virus | lower respiratory tract infection |
| 219921000006115 | Pneumonia due to pleuropneumonia like organisms | lower respiratory tract infection |
| 4781201000006120 | Pneumonia due to pleuropneumonia-like organism | lower respiratory tract infection |
| 219931000006117 | Pneumonia due to proteus | lower respiratory tract infection |
| 3130351000006120 | Pneumonia due to Proteus mirabilis | lower respiratory tract infection |
| 3166801000006110 | Pneumonia due to Pseudomonas | lower respiratory tract infection |
| 301362011 | Pneumonia due to respiratory syncytial virus | lower respiratory tract infection |
| 301382010 | Pneumonia due to specified organism NOS | lower respiratory tract infection |
| 7253251000006110 | Pneumonia due to Staphylococcus aureus | lower respiratory tract infection |
| 3047811000006120 | Pneumonia due to Streptococcus | lower respiratory tract infection |
| 219991000006118 | Pneumonia due to streptococcus, group B | lower respiratory tract infection |
| 4200921000006120 | Pneumonia in aspergillosis | lower respiratory tract infection |
| 3465961000006110 | Pneumonia in pertussis | lower respiratory tract infection |
| 885241000006112 | Pneumonia NOS | lower respiratory tract infection |
| 301431011 | Pneumonia or influenza NOS | lower respiratory tract infection |
| 220021000006118 | Pneumonia with actinomycosis | lower respiratory tract infection |
| 301389018 | Pneumonia with anthrax | lower respiratory tract infection |
| 1219703018 | Pneumonia with aspergillosis | lower respiratory tract infection |
| 220051000006110 | Pneumonia with candidiasis | lower respiratory tract infection |
| 301391014 | Pneumonia with coccidioidomycosis | lower respiratory tract infection |
| 220071000006117 | Pneumonia with cytomegalic inclusion disease | lower respiratory tract infection |
| 301392019 | Pneumonia with histoplasmosis | lower respiratory tract infection |
| 396104016 | Pneumonia with infectious diseases EC | lower respiratory tract infection |
| 301404013 | Pneumonia with infectious diseases EC NOS | lower respiratory tract infection |
| 220111000006113 | Pneumonia with measles | lower respiratory tract infection |
| 301397013 | Pneumonia with nocardiasis | lower respiratory tract infection |
| 220131000006119 | Pneumonia with ornithosis | lower respiratory tract infection |
| 301394018 | Pneumonia with other infectious diseases EC | lower respiratory tract infection |
| 301403019 | Pneumonia with other infectious diseases EC NOS | lower respiratory tract infection |
| 301390010 | Pneumonia with other systemic mycoses | lower respiratory tract infection |
| 1231962012 | Pneumonia with pertussis | lower respiratory tract infection |
| 220191000006115 | Pneumonia with Q-fever | lower respiratory tract infection |
| 301393012 | Pneumonia with systemic mycosis NOS | lower respiratory tract infection |
| 494463016 | Pneumonia with toxoplasmosis | lower respiratory tract infection |
| 1216638013 | Pneumonia with tularaemia | lower respiratory tract infection |
| 220241000006115 | Pneumonia with typhoid fever | lower respiratory tract infection |
| 1231963019 | Pneumonia with whooping cough | lower respiratory tract infection |
| 885271000006116 | Pneumonia/influenza NOS | lower respiratory tract infection |
| 286339014 | Pneumonic plague | lower respiratory tract infection |
| 4764761000006110 | Post measles pneumonia | lower respiratory tract infection |
| 6348731000006110 | Post obstructive pneumonia | lower respiratory tract infection |
| 294997017 | Postmeasles pneumonia | lower respiratory tract infection |
| 216551000006114 | Postoperative lower respiratory tract infection | lower respiratory tract infection |
| 459416013 | Postoperative pneumonia | lower respiratory tract infection |
| 58969011 | Primary pneumonic plague | lower respiratory tract infection |
| 287649014 | Primary pulmonary blastomycosis | lower respiratory tract infection |
| 145932018 | Primary pulmonary coccidioidomycosis | lower respiratory tract infection |
| 4781151000006110 | Proteus pneumonia | lower respiratory tract infection |
| 69026013 | Pseudomonal pneumonia | lower respiratory tract infection |
| 36656012 | Pulmonary actinomycosis | lower respiratory tract infection |
| 195367012 | Pulmonary anthrax | lower respiratory tract infection |
| 193491000006112 | Pulmonary aspergillosis | lower respiratory tract infection |
| 350063010 | Pulmonary blastomycosis | lower respiratory tract infection |
| 35229011 | Pulmonary cryptococcosis | lower respiratory tract infection |
| 193801000006119 | Pulmonary disease due to Mycobacteria | lower respiratory tract infection |
| 7967281000006120 | Pulmonary embolism with pulmonary infarction | lower respiratory tract infection |
| 287632019 | Pulmonary histoplasmosis | lower respiratory tract infection |
| 4411761000006110 | Pulmonary infection | lower respiratory tract infection |
| 350061012 | Pulmonary mucormycosis | lower respiratory tract infection |
| 4741561000006110 | Pulmonary Mycobacterium avium complex infection | lower respiratory tract infection |
| 1229247010 | Pulmonary Mycobacterium avium-intracellulare infection | lower respiratory tract infection |
| 4591015 | Pulmonary nocardiosis | lower respiratory tract infection |
| 122874011 | Pulmonary paracoccidioidomycosis | lower respiratory tract infection |
| 155111000006115 | Pulmonary schistosomiasis | lower respiratory tract infection |
| 75466014 | Pulmonary sporotrichosis | lower respiratory tract infection |
| 239551014 | Pulmonary tuberculosis | lower respiratory tract infection |
| 286222014 | Pulmonary tuberculosis NOS | lower respiratory tract infection |
| 1231843013 | Purulent pleurisy | lower respiratory tract infection |
| 7129221000006120 | Recurrent aspiration pneumonia | lower respiratory tract infection |
| 451444011 | Recurrent lower respiratory tract infection | lower respiratory tract infection |
| 7515921000006110 | Recurrent pneumonia | lower respiratory tract infection |
| 183191000006115 | Recurrent wheezy bronchitis | lower respiratory tract infection |
| 1786171000006120 | Respiratory bronchiolitis associated interstitial lung disease | lower respiratory tract infection |
| 1856531000006120 | Respiratory infection aggravates symptom | lower respiratory tract infection |
| 175471000006116 | Respiratory infection NOS | lower respiratory tract infection |
| 3427851000006110 | Respiratory syncytial virus bronchiolitis | lower respiratory tract infection |
| 411488017 | Respiratory tract infection | lower respiratory tract infection |
| 13455018 | Rheumatic pneumonia | lower respiratory tract infection |
| 5887761000006110 | Right basal pneumonia | lower respiratory tract infection |
| 5887791000006110 | Right lower lobe pneumonia | lower respiratory tract infection |
| 1772741000006120 | Right lower zone pneumonia | lower respiratory tract infection |
| 5887871000006110 | Right middle lobe pneumonia | lower respiratory tract infection |
| 1772751000006120 | Right middle zone pneumonia | lower respiratory tract infection |
| 5887921000006110 | Right upper lobe pneumonia | lower respiratory tract infection |
| 1772761000006110 | Right upper zone pneumonia | lower respiratory tract infection |
| 5887781000006120 | RLL - Right lower lobe pneumonia | lower respiratory tract infection |
| 5887771000006120 | RLZ - Right lower zone pneumonia | lower respiratory tract infection |
| 5573831000006110 | RTI - Respiratory tract infection | lower respiratory tract infection |
| 5887901000006110 | RUL - Right upper lobe pneumonia | lower respiratory tract infection |
| 5303018 | Salmonella pneumonia | lower respiratory tract infection |
| 5983281000006110 | Secondary bacterial pneumonia | lower respiratory tract infection |
| 110441014 | Secondary pneumonic plague | lower respiratory tract infection |
| 301670014 | Serofibrinous pleurisy NOS | lower respiratory tract infection |
| 301671013 | Serous pleurisy NOS | lower respiratory tract infection |
| 6320012 | Staphylococcal pleurisy | lower respiratory tract infection |
| 39802016 | Staphylococcal pleurisy with effusion | lower respiratory tract infection |
| 219971000006119 | Staphylococcal pneumonia | lower respiratory tract infection |
| 301652011 | Sterile pleurisy | lower respiratory tract infection |
| 141587016 | Streptococcal pleurisy | lower respiratory tract infection |
| 12711015 | Streptococcal pleurisy with effusion | lower respiratory tract infection |
| 56816017 | Streptococcal pneumonia | lower respiratory tract infection |
| 2724991000000110 | Suspected whooping cough | lower respiratory tract infection |
| 4514611000006120 | TB - Pulmonary tuberculosis | lower respiratory tract infection |
| 4781831000006110 | Toxic bronchiolitis obliterans | lower respiratory tract infection |
| 287864011 | Toxoplasma pneumonitis | lower respiratory tract infection |
| 5055021000006120 | Traumatic pneumonia | lower respiratory tract infection |
| 24683016 | Tuberculous empyema | lower respiratory tract infection |
| 2730071000006110 | Tuberculous pleural empyema | lower respiratory tract infection |
| 286225011 | Tuberculous pleurisy | lower respiratory tract infection |
| 286214019 | Tuberculous pleurisy in primary progressive tuberculosis | lower respiratory tract infection |
| 286230010 | Tuberculous pleurisy NOS | lower respiratory tract infection |
| 83211000006113 | Tuberculous pleurisy, confirmed bacteriologically and histologically | lower respiratory tract infection |
| 132731014 | Tuberculous pneumonia | lower respiratory tract infection |
| 3231041000006110 | Typhoid pneumonia | lower respiratory tract infection |
| 3580251000006110 | Unresolved lobar pneumonia | lower respiratory tract infection |
| 3437661000006110 | Unresolved pneumonia | lower respiratory tract infection |
| 7100861000006110 | VAP - ventilator associated pneumonia | lower respiratory tract infection |
| 4781311000006110 | Varicella pneumonia | lower respiratory tract infection |
| 301400016 | Varicella pneumonitis | lower respiratory tract infection |
| 7100831000006120 | Ventilator associated pneumonia | lower respiratory tract infection |
| 7100851000006110 | Ventilator-acquired pneumonia | lower respiratory tract infection |
| 6013161000006120 | Viral lower respiratory infection | lower respiratory tract infection |
| 6041781000006120 | Viral pleurisy | lower respiratory tract infection |
| 125510013 | Viral pneumonia | lower respiratory tract infection |
| 301363018 | Viral pneumonia NEC | lower respiratory tract infection |
| 301364012 | Viral pneumonia NOS | lower respiratory tract infection |
| 6013151000006120 | Viral respiratory infection | lower respiratory tract infection |
| 46589015 | Whooping cough | lower respiratory tract infection |
| 286448015 | Whooping cough - other specified organism | lower respiratory tract infection |
| 286452015 | Whooping cough NOS | lower respiratory tract infection |
| 885221000006117 | Whooping cough pneumonia | lower respiratory tract infection |
| 5526131000006110 | Whooping cough-like syndrome | lower respiratory tract infection |
| 3483291000006120 | Whooping respiration | lower respiratory tract infection |
| 299070012 | Recurrent acute otitis media | acute otitis media |
| 399498017 | Otitis media | acute otitis media |
| 299531012 | Viral ear infection | acute otitis media |
| 7628271000006110 | acute exacerbation of extrinsic asthma | asthma exacerbation |

**Appendix 2.** ICD-10 codes for infections of interest

| **ICD code** | **Chapter** | **Block** | **Category/sub-category** | **Infection of interest** |
| --- | --- | --- | --- | --- |
| H65.0 | Mastoiditis and other ear infection complications | Diseases of middle ear and mastoid | Acute serous otitis media | Otitis media |
| H65.1 | Mastoiditis and other ear infection complications | Diseases of middle ear and mastoid | Other acute nonsuppurative otitis media | Otitis media |
| H65.9 | Mastoiditis and other ear infection complications | Diseases of middle ear and mastoid | Nonsuppurative otitis media, unspecified | Otitis media |
| H66.0 | Mastoiditis and other ear infection complications | Diseases of middle ear and mastoid | Acute suppurative otitis media | Otitis media |
| H66.4 | Mastoiditis and other ear infection complications | Diseases of middle ear and mastoid | Suppurative otitis media, unspecified | Otitis media |
| H66.9 | Mastoiditis and other ear infection complications | Diseases of middle ear and mastoid | Otitis media, unspecified | Otitis media |
| H67.0 | Mastoiditis and other ear infection complications | Diseases of middle ear and mastoid | Otitis media, unspecified | Otitis media |
| H67.8 | Mastoiditis and other ear infection complications | Diseases of middle ear and mastoid | Otitis media in bacterial diseases classified elsewhere | Otitis media |
| H70 | Mastoiditis and other ear infection complications | Diseases of middle ear and mastoid | Mastoiditis and related conditions | Otitis media |
| H74 | Mastoiditis and other ear infection complications | Diseases of middle ear and mastoid | Other disorders of middle ear mastoid | Otitis media |
| H75 | Mastoiditis and other ear infection complications | Diseases of middle ear and mastoid | Other disorders of middle ear and mastoid in diseases classified elsewhere | Otitis media |
| A36.0 | Certain infectious and parasitic diseases | Other bacterial diseases | Pharyngeal diphtheria | Respiratory tract infections |
| A36.1 | Certain infectious and parasitic diseases | Other bacterial diseases | Nasopharyngeal diphtheria | Respiratory tract infections |
| A36.2 | Certain infectious and parasitic diseases | Other bacterial diseases | Laryngeal diphtheria | Respiratory tract infections |
| J00 | Diseases of the respiratory system | Acute upper respiratory tract infections | Acute nasopharyngitis | Respiratory tract infections |
| J01 | Diseases of the respiratory system | Acute upper respiratory tract infections | Acute sinusitis | Respiratory tract infections |
| J02 | Diseases of the respiratory system | Acute upper respiratory tract infections | Acute pharyngitis | Respiratory tract infections |
| J03 | Diseases of the respiratory system | Acute upper respiratory tract infections | Acute tonsillitis | Respiratory tract infections |
| J04 | Diseases of the respiratory system | Acute upper respiratory tract infections | Acute laryngitis and tracheitis | Respiratory tract infections |
| J05 | Diseases of the respiratory system | Acute upper respiratory tract infections | Acute obstructive laryngitis [croup] and epiglottitis | Respiratory tract infections |
| J06 | Diseases of the respiratory system | Acute upper respiratory tract infections | Acute upper respiratory infections of multiple and unspecified sites | Respiratory tract infections |
| J12 | Diseases of the respiratory system | Influenza and pneumonia | Viral pneumonia, not elsewhere classified | Respiratory tract infections |
| J13 | Diseases of the respiratory system | Influenza and pneumonia | Pneumonia due to Streptococcus pneumoniae | Respiratory tract infections |
| J14 | Diseases of the respiratory system | Influenza and pneumonia | Pneumonia due to Haemophilus influenzae | Respiratory tract infections |
| J15 | Diseases of the respiratory system | Influenza and pneumonia | Bacterial pneumonia, not elsewhere classified | Respiratory tract infections |
| J16 | Diseases of the respiratory system | Influenza and pneumonia | Pneumonia due to other infectious organisms, not elsewhere classified | Respiratory tract infections |
| J17 | Diseases of the respiratory system | Influenza and pneumonia | Pneumonia in diseases classified elsewhere | Respiratory tract infections |
| J18 | Diseases of the respiratory system | Influenza and pneumonia | Pneumonia, organism unspecified | Respiratory tract infections |
| J20 | Diseases of the respiratory system | Other acute lower respiratory infections | Acute bronchitis | Respiratory tract infections |
| J21 | Diseases of the respiratory system | Other acute lower respiratory infections | Acute bronchiolitis | Respiratory tract infections |
| J22 | Diseases of the respiratory system | Other acute lower respiratory infections | Unspecified acute lower respiratory infection | Respiratory tract infections |
| J36 | Diseases of the respiratory system | Other diseases of upper respiratory tract | Peritonsillar abscess | Respiratory tract infections |
| J39.0 | Diseases of the respiratory system | Other diseases of upper respiratory tract | Retropharyngeal and parapharyngeal abscess | Respiratory tract infections |
| J39.1 | Diseases of the respiratory system | Other diseases of upper respiratory tract | Other abscess of pharynx | Respiratory tract infections |
| J41 | Diseases of the respiratory system | Chronic lower respiratory diseases | Simple and mucopurulent chronic bronchitis | Respiratory tract infections |
| J42 | Diseases of the respiratory system | Chronic lower respiratory diseases | Unspecified chronic bronchitis | Respiratory tract infections |
| J43 | Diseases of the respiratory system | Chronic lower respiratory diseases | Emphysema | Respiratory tract infections |
| J44 | Diseases of the respiratory system | Chronic lower respiratory diseases | COPD | Respiratory tract infections |
| J45 | Diseases of the respiratory system | Chronic lower respiratory diseases | Asthma | Respiratory tract infections |
| J98.7 | Diseases of the respiratory system | Other diseases of the respiratory system | Respiratory infections, not elsewhere classified (Respiratory (tract) infections not specified as acute, chronic, lower, or upper) | Respiratory tract infections |
| J98.8 | Diseases of the respiratory system | Other diseases of the respiratory system | Other specified respiratory disorders | Respiratory tract infections |
| R06 | Symptoms, signs and abnormal clinical and laboratory findings, not elsewhere classified | Symptoms and signs involving the circulatory and respiratory systems | Abnormalities of breathing | Respiratory tract infections |
| R05 | Symptoms, signs and abnormal clinical and laboratory findings, not elsewhere classified | Symptoms and signs involving the circulatory and respiratory systems | Cough | Respiratory tract infections |
| R07.0 | Symptoms, signs and abnormal clinical and laboratory findings, not elsewhere classified | Pain in throat and chest | Pain in throat | Respiratory tract infections |
| R09.3 | Symptoms, signs and abnormal clinical and laboratory findings, not elsewhere classified | Other symptoms and signs involving the circulatory and respiratory systems | Abnormal sputum | Respiratory tract infections |
| A36.3 | Certain infectious and parasitic diseases | Other bacterial diseases | Cutaneous diphtheria | Skin infections |
| A46 | Certain infectious and parasitic diseases | Other bacterial diseases | Erysipelas | Skin infections |
| K12.2 | Diseases of the digestive system | Diseases of oral cavity, salivary glands and jaws | Cellulitis and abscess of mouth | Skin infections |
| L00 | Diseases of the skin and subcutaneous tissue | Infections of the skin and subcutaneous tissue | Staphylococcal scalded skin syndrome | Skin infections |
| L01 | Diseases of the skin and subcutaneous tissue | Infections of the skin and subcutaneous tissue | Impetigo | Skin infections |
| L02 | Diseases of the skin and subcutaneous tissue | Infections of the skin and subcutaneous tissue | Cutaneous abscess, furuncle and carbuncle | Skin infections |
| L03 | Diseases of the skin and subcutaneous tissue | Infections of the skin and subcutaneous tissue | Cellulitis | Skin infections |
| L04 | Diseases of the skin and subcutaneous tissue | Infections of the skin and subcutaneous tissue | Acute lymphadenitis | Skin infections |
| L08.0 | Diseases of the skin and subcutaneous tissue | Infections of the skin and subcutaneous tissue | Pyoderma (excl pyoderma gangrenosum) | Skin infections |
| L08.8 | Diseases of the skin and subcutaneous tissue | Infections of the skin and subcutaneous tissue | Other spec local infections of skin and subcutaneous tissue | Skin infections |
| L08.9 | Diseases of the skin and subcutaneous tissue | Infections of the skin and subcutaneous tissue | Local infection of skin and subcutaneous tissue unspecified | Skin infections |
| N10 | Diseases of the genitourinary system | Renal tubulo-interstitial diseases | Acute tubulo-interstitial nephritis | Urinary tract infections |
| N12 | Diseases of the genitourinary system | Renal tubulo-interstitial diseases | Tubulo-interstitial nephritis, not specified as acute or chronic | Urinary tract infections |
| N13.6 | Diseases of the genitourinary system | Renal tubulo-interstitial diseases | Pyenephrosis | Urinary tract infections |
| N15.1 | Diseases of the genitourinary system | Renal tubulo-interstitial diseases | Renal and perinephric abscess | Urinary tract infections |
| N15.9 | Diseases of the genitourinary system | Renal tubulo-interstitial diseases | Renal tubule-interstitial disease, unspecified | Urinary tract infections |
| N16.0 | Diseases of the genitourinary system | Renal tubulo-interstitial diseases | Renal tubule-interstitial disorders in infectious and parasitic diseases | Urinary tract infections |
| N30.0 | Diseases of the genitourinary system | Other diseases of urinary system | Cystitis | Urinary tract infections |
| N30.8 | Diseases of the genitourinary system | Other diseases of urinary system | Other cystitis - abscess of bladder | Urinary tract infections |
| N30.9 | Diseases of the genitourinary system | Other diseases of urinary system | Cystitis, unspecified | Urinary tract infections |
| N39.0 | Diseases of the genitourinary system | Other diseases of urinary system | Urinary tract infection, site not specified | Urinary tract infections |
| N41.0 | Diseases of the genitourinary system | Diseases of male genital organs | Acute prostatitis | Urinary tract infections |
| N41.2 | Diseases of the genitourinary system | Diseases of male genital organs | Abscess of prostate | Urinary tract infections |
| N41.3 | Diseases of the genitourinary system | Diseases of male genital organs | Prostatocystitis | Urinary tract infections |
| N41.9 | Diseases of the genitourinary system | Diseases of male genital organs | Inflammatory disease of prostate, unspecified | Urinary tract infections |
| N45 | Diseases of the genitourinary system | Diseases of male genital organs | Orchitis and epidymitis | Urinary tract infections |
| A38 | Certain infectious and parasitic diseases | Other bacterial diseases | Scarlet fever | Respiratory tract infections |

**Appendix 3.** List of systematic antibiotics

| **ProdCodeId** | **Product Name** | **Drug subtance name** | **Pharmacological class** | **ATC code** | **AWaRe** | **EML** |
| --- | --- | --- | --- | --- | --- | --- |
| 3079441000033110 | Amoxicillin 250mg powder for solution for injection vials | Amoxicillin sodium | Penicillins | J01CA04 | Access | Yes |
| 3079541000033110 | Amoxicillin 500mg powder for solution for injection vials | Amoxicillin sodium | Penicillins | J01CA04 | Access | Yes |
| 48441000033113 | Amoxil 1g powder for solution for injection vials | Amoxicillin sodium | Penicillins | J01CA04 | Access | Yes |
| 48541000033114 | Amoxil 250mg powder for solution for injection vials | Amoxicillin sodium | Penicillins | J01CA04 | Access | Yes |
| 48641000033110 | Amoxil 500mg powder for solution for injection vials | Amoxicillin sodium | Penicillins | J01CA04 | Access | Yes |
| 3079341000033110 | Amoxicillin 1g powder for solution for injection vials | Amoxicillin sodium | Penicillins | J01CA04 | Access | Yes |
| 37241000033112 | Almodan 125mg/5ml syrup | Amoxicillin trihydrate | Penicillins | J01CA04 | Access | Yes |
| 44041000033113 | Amoxil 250mg capsules | Amoxicillin trihydrate | Penicillins | J01CA04 | Access | Yes |
| 44141000033112 | Amoxil 500mg capsules | Amoxicillin trihydrate | Penicillins | J01CA04 | Access | Yes |
| 45441000033117 | Amoram 250mg capsules | Amoxicillin trihydrate | Penicillins | J01CA04 | Access | Yes |
| 45541000033116 | Amoram 500mg capsules | Amoxicillin trihydrate | Penicillins | J01CA04 | Access | Yes |
| 45641000033115 | Amix 500 capsules | Amoxicillin trihydrate | Penicillins | J01CA04 | Access | Yes |
| 46141000033118 | Amix 250 capsules | Amoxicillin trihydrate | Penicillins | J01CA04 | Access | Yes |
| 52841000033110 | Amoxil 125mg/1.25ml paediatric oral suspension | Amoxicillin trihydrate | Penicillins | J01CA04 | Access | Yes |
| 53941000033113 | Amoxil 3g oral powder sachets sucrose free | Amoxicillin trihydrate | Penicillins | J01CA04 | Access | Yes |
| 55641000033117 | Amix 125 oral suspension | Amoxicillin trihydrate | Penicillins | J01CA04 | Access | Yes |
| 55741000033114 | Amix 250 oral suspension | Amoxicillin trihydrate | Penicillins | J01CA04 | Access | Yes |
| 55941000033112 | Amoxil 125mg/5ml syrup sucrose free | Amoxicillin trihydrate | Penicillins | J01CA04 | Access | Yes |
| 56041000033119 | Amoxil 250mg/5ml syrup sucrose free | Amoxicillin trihydrate | Penicillins | J01CA04 | Access | Yes |
| 56841000033114 | Amoram 125mg/5ml oral suspension | Amoxicillin trihydrate | Penicillins | J01CA04 | Access | Yes |
| 56941000033118 | Amoram 250mg/5ml oral suspension | Amoxicillin trihydrate | Penicillins | J01CA04 | Access | Yes |
| 620241000033118 | Galenamox 250mg capsules | Amoxicillin trihydrate | Penicillins | J01CA04 | Access | Yes |
| 620341000033111 | Galenamox 500mg capsules | Amoxicillin trihydrate | Penicillins | J01CA04 | Access | Yes |
| 625841000033114 | Galenamox 250mg/5ml oral suspension | Amoxicillin trihydrate | Penicillins | J01CA04 | Access | Yes |
| 625941000033118 | Galenamox 125mg/5ml oral suspension | Amoxicillin trihydrate | Penicillins | J01CA04 | Access | Yes |
| 1671741000033110 | Respillin 250mg capsules | Amoxicillin trihydrate | Penicillins | J01CA04 | Access | Yes |
| 1671841000033110 | Respillin 500mg capsules | Amoxicillin trihydrate | Penicillins | J01CA04 | Access | Yes |
| 3079141000033110 | Amoxicillin 250mg capsules | Amoxicillin trihydrate | Penicillins | J01CA04 | Access | Yes |
| 3079241000033110 | Amoxicillin 500mg capsules | Amoxicillin trihydrate | Penicillins | J01CA04 | Access | Yes |
| 3079641000033110 | Amoxicillin 125mg/5ml oral suspension | Amoxicillin trihydrate | Penicillins | J01CA04 | Access | Yes |
| 3079741000033110 | Amoxicillin 250mg/5ml oral suspension | Amoxicillin trihydrate | Penicillins | J01CA04 | Access | Yes |
| 3079841000033110 | Amoxicillin 125mg/1.25ml oral suspension paediatric | Amoxicillin trihydrate | Penicillins | J01CA04 | Access | Yes |
| 3079941000033110 | Amoxicillin 3g oral powder sachets sugar free | Amoxicillin trihydrate | Penicillins | J01CA04 | Access | Yes |
| 3080041000033110 | Amoxicillin 125mg/5ml oral suspension sugar free | Amoxicillin trihydrate | Penicillins | J01CA04 | Access | Yes |
| 3080141000033110 | Amoxicillin 250mg/5ml oral suspension sugar free | Amoxicillin trihydrate | Penicillins | J01CA04 | Access | Yes |
| 13707941000033100 | Amoxicillin 1g dispersible tablets sugar free | Amoxicillin trihydrate | Penicillins | J01CA04 | Access | Yes |
| 13827741000033100 | Amoxicillin 500mg/5ml oral suspension sugar free | Amoxicillin trihydrate | Penicillins | J01CA04 | Access | Yes |
| 1051841000033110 | Penbritin 500mg powder for solution for injection vials | Ampicillin sodium | Penicillins | J01CA01 | Access | Yes |
| 50641000033110 | Ampicillin 500mg powder for solution for injection vials | Ampicillin sodium | Penicillins | J01CA01 | Access | Yes |
| 44441000033116 | Ampicillin 250mg capsules | Ampicillin | Penicillins | J01CA01 | Access | Yes |
| 44541000033115 | Ampicillin 500mg capsules | Ampicillin | Penicillins | J01CA01 | Access | Yes |
| 52041000033115 | Ampicillin 125mg/5ml oral suspension | Ampicillin | Penicillins | J01CA01 | Access | Yes |
| 52141000033116 | Ampicillin 250mg/5ml oral suspension | Ampicillin | Penicillins | J01CA01 | Access | Yes |
| 1044041000033110 | Penbritin 250mg capsules | Ampicillin | Penicillins | J01CA01 | Access | Yes |
| 1044141000033110 | Penbritin 500mg capsules | Ampicillin | Penicillins | J01CA01 | Access | Yes |
| 1063241000033110 | Penbritin 125mg/5ml syrup | Ampicillin | Penicillins | J01CA01 | Access | Yes |
| 1063341000033110 | Penbritin Forte 250mg/5ml syrup | Ampicillin | Penicillins | J01CA01 | Access | Yes |
| 8884841000033110 | Azithromycin 500mg powder for solution for infusion vials | Azithromycin dihydrate | Macrolides | J01FA10 | Watch | Yes |
| 96941000033113 | Azithromycin 250mg capsules | Azithromycin dihydrate | Macrolides | J01FA10 | Watch | Yes |
| 98441000033116 | Azithromycin 200mg/5ml oral suspension | Azithromycin | Macrolides | J01FA10 | Watch | Yes |
| 1553141000033110 | Zithromax 250mg capsules | Azithromycin dihydrate | Macrolides | J01FA10 | Watch | Yes |
| 1557141000033110 | Zithromax 200mg/5ml oral suspension | Azithromycin | Macrolides | J01FA10 | Watch | Yes |
| 1572841000033110 | Azithromycin 500mg tablets | Azithromycin | Macrolides | J01FA10 | Watch | Yes |
| 1713741000033110 | Zithromax 500mg tablets | Azithromycin | Macrolides | J01FA10 | Watch | Yes |
| 3955441000033110 | Azithromycin 250mg tablets | Azithromycin | Macrolides | J01FA10 | Watch | Yes |
| 4823141000033110 | Clamelle 500mg tablets | Azithromycin | Macrolides | J01FA10 | Watch | Yes |
| 5836141000033110 | Aztreonam 75mg powder and solvent for nebuliser solution vials with device | Aztreonam lysine | Monobactams | J01DF01 | Reserve | No |
| 5836241000033110 | Cayston 75mg powder and solvent for nebuliser solution vials with Altera Nebuliser Handset | Aztreonam lysine | Monobactams | J01DF01 | Reserve | No |
| 97841000033119 | Aztreonam 1g powder for solution for injection vials | Aztreonam | Monobactams | J01DF01 | Reserve | No |
| 97941000033110 | Aztreonam 2g powder for solution for injection vials | Aztreonam | Monobactams | J01DF01 | Reserve | No |
| 98041000033113 | Aztreonam 500mg powder for solution for injection vials | Aztreonam | Monobactams | J01DF01 | Reserve | No |
| 5369941000033110 | Benzathine benzylpenicillin 2.4million unit powder and solvent for suspension for injection vials | Benzathine benzylpenicillin | Penicillins | J01CE08 | Access | Yes |
| 13418641000033100 | Benzathine benzylpenicillin 1.2million unit powder and solvent for suspension for injection vials | Benzathine benzylpenicillin | Penicillins | J01CE08 | Access | Yes |
| 4456141000033110 | Extencilline 2.4million unit powder and solvent for suspension for injection vials | Benzathine benzylpenicillin | Penicillins | J01CE08 | Access | Yes |
| 130941000033113 | Benzylpenicillin 600mg powder for solution for injection vials | Benzylpenicillin sodium | Penicillins | J01CE01 | Access | Yes |
| 1769641000033110 | Benzylpenicillin 1.2g powder for solution for injection vials | Benzylpenicillin sodium | Penicillins | J01CE01 | Access | Yes |
| 1769741000033110 | Crystapen 1.2g powder for solution for injection vials | Benzylpenicillin sodium | Penicillins | J01CE01 | Access | Yes |
| 386341000033112 | Crystapen 600mg powder for solution for injection vials | Benzylpenicillin sodium | Penicillins | J01CE01 | Access | Yes |
| 219741000033111 | Cefaclor 500mg capsules | Cefaclor monohydrate | Second-generation-cephalosporins | J01DC04 | Watch | No |
| 220441000033115 | Cefaclor 250mg capsules | Cefaclor monohydrate | Second-generation-cephalosporins | J01DC04 | Watch | No |
| 227741000033114 | Cefaclor 375mg modified-release tablets | Cefaclor monohydrate | Second-generation-cephalosporins | J01DC04 | Watch | No |
| 230741000033112 | Cefaclor 125mg/5ml oral suspension sugar free | Cefaclor monohydrate | Second-generation-cephalosporins | J01DC04 | Watch | No |
| 230841000033119 | Cefaclor 250mg/5ml oral suspension sugar free | Cefaclor monohydrate | Second-generation-cephalosporins | J01DC04 | Watch | No |
| 230941000033110 | Cefaclor 125mg/5ml oral suspension | Cefaclor monohydrate | Second-generation-cephalosporins | J01DC04 | Watch | No |
| 231741000033119 | Cefaclor 250mg/5ml oral suspension | Cefaclor monohydrate | Second-generation-cephalosporins | J01DC04 | Watch | No |
| 433941000033116 | Distaclor 500mg capsules | Cefaclor monohydrate | Second-generation-cephalosporins | J01DC04 | Watch | No |
| 452341000033110 | Distaclor MR 375mg tablets | Cefaclor monohydrate | Second-generation-cephalosporins | J01DC04 | Watch | No |
| 460341000033118 | Distaclor 125mg/5ml oral suspension | Cefaclor monohydrate | Second-generation-cephalosporins | J01DC04 | Watch | No |
| 460441000033112 | Distaclor 250mg/5ml oral suspension | Cefaclor monohydrate | Second-generation-cephalosporins | J01DC04 | Watch | No |
| 797641000033116 | Keftid 250mg capsules | Cefaclor monohydrate | Second-generation-cephalosporins | J01DC04 | Watch | No |
| 797741000033113 | Keftid 500mg capsules | Cefaclor monohydrate | Second-generation-cephalosporins | J01DC04 | Watch | No |
| 803141000033116 | Keftid 125mg/5ml oral suspension | Cefaclor monohydrate | Second-generation-cephalosporins | J01DC04 | Watch | No |
| 803241000033111 | Keftid 250mg/5ml oral suspension | Cefaclor monohydrate | Second-generation-cephalosporins | J01DC04 | Watch | No |
| 2617541000033110 | Bacticlor MR 375mg tablets | Cefaclor monohydrate | Second-generation-cephalosporins | J01DC04 | Watch | No |
| 104041000033114 | Baxan 500mg capsules | Cefadroxil monohydrate | First-generation-cephalosporins | J01DB05 | Access | No |
| 115841000033118 | Baxan 125mg/5ml oral suspension | Cefadroxil monohydrate | First-generation-cephalosporins | J01DB05 | Access | No |
| 115941000033114 | Baxan 250mg/5ml oral suspension | Cefadroxil monohydrate | First-generation-cephalosporins | J01DB05 | Access | No |
| 116041000033116 | Baxan 500mg/5ml oral suspension | Cefadroxil monohydrate | First-generation-cephalosporins | J01DB05 | Access | No |
| 220541000033119 | Cefadroxil 500mg capsules | Cefadroxil monohydrate | First-generation-cephalosporins | J01DB05 | Access | No |
| 231041000033117 | Cefadroxil 125mg/5ml oral suspension | Cefadroxil monohydrate | First-generation-cephalosporins | J01DB05 | Access | No |
| 231541000033110 | Cefadroxil 250mg/5ml oral suspension | Cefadroxil monohydrate | First-generation-cephalosporins | J01DB05 | Access | No |
| 231641000033111 | Cefadroxil 500mg/5ml oral suspension | Cefadroxil monohydrate | First-generation-cephalosporins | J01DB05 | Access | No |
| 220941000033113 | Ceporex 250mg capsules | Cefalexin | First-generation-cephalosporins | J01DB01 | Access | Yes |
| 221041000033115 | Ceporex 500mg capsules | Cefalexin | First-generation-cephalosporins | J01DB01 | Access | Yes |
| 232341000033112 | Ceporex 125mg/5ml syrup | Cefalexin | First-generation-cephalosporins | J01DB01 | Access | Yes |
| 232441000033118 | Ceporex 250mg/5ml syrup | Cefalexin | First-generation-cephalosporins | J01DB01 | Access | Yes |
| 232541000033117 | Ceporex 500mg/5ml syrup | Cefalexin | First-generation-cephalosporins | J01DB01 | Access | Yes |
| 235141000033115 | Ceporex 250mg tablets | Cefalexin | First-generation-cephalosporins | J01DB01 | Access | Yes |
| 235241000033110 | Ceporex 500mg tablets | Cefalexin | First-generation-cephalosporins | J01DB01 | Access | Yes |
| 796841000033118 | Keflex 250mg capsules | Cefalexin | First-generation-cephalosporins | J01DB01 | Access | Yes |
| 796941000033114 | Keflex 500mg capsules | Cefalexin | First-generation-cephalosporins | J01DB01 | Access | Yes |
| 802741000033110 | Keflex 125mg/5ml oral suspension | Cefalexin | First-generation-cephalosporins | J01DB01 | Access | Yes |
| 802841000033117 | Keflex 250mg/5ml oral suspension | Cefalexin | First-generation-cephalosporins | J01DB01 | Access | Yes |
| 803341000033118 | Keflex 250mg tablets | Cefalexin | First-generation-cephalosporins | J01DB01 | Access | Yes |
| 803441000033112 | Keflex 500mg tablets | Cefalexin | First-generation-cephalosporins | J01DB01 | Access | Yes |
| 3084041000033110 | Cefalexin 250mg capsules | Cefalexin | First-generation-cephalosporins | J01DB01 | Access | Yes |
| 3084141000033110 | Cefalexin 500mg capsules | Cefalexin | First-generation-cephalosporins | J01DB01 | Access | Yes |
| 3084241000033110 | Cefalexin 250mg tablets | Cefalexin | First-generation-cephalosporins | J01DB01 | Access | Yes |
| 3084341000033110 | Cefalexin 500mg tablets | Cefalexin | First-generation-cephalosporins | J01DB01 | Access | Yes |
| 3084441000033110 | Cefalexin 125mg/5ml oral suspension | Cefalexin | First-generation-cephalosporins | J01DB01 | Access | Yes |
| 3084541000033110 | Cefalexin 250mg/5ml oral suspension | Cefalexin | First-generation-cephalosporins | J01DB01 | Access | Yes |
| 3084641000033110 | Cefalexin 500mg/5ml oral suspension | Cefalexin | First-generation-cephalosporins | J01DB01 | Access | Yes |
| 4373141000033110 | Cefalexin 250mg/5ml oral suspension sugar free | Cefalexin | First-generation-cephalosporins | J01DB01 | Access | Yes |
| 5747941000033110 | Cefalexin 125mg/5ml oral suspension sugar free | Cefalexin | First-generation-cephalosporins | J01DB01 | Access | Yes |
| 799041000033118 | Kefadol 1g powder for solution for injection vials | Cefamandole nafate | Second-generation-cephalosporins | J01DC03 | Watch | No |
| 12880241000033100 | Cefazolin 2g powder for solution for injection vials | Cefazolin sodium | First-generation-cephalosporins | J01DB04 | Access | Yes |
| 799441000033110 | Kefzol 1g powder for solution for injection vials | Cefazolin sodium | First-generation-cephalosporins | J01DB04 | Access | Yes |
| 799541000033111 | Kefzol 500mg powder for solution for injection vials | Cefazolin sodium | First-generation-cephalosporins | J01DB04 | Access | Yes |
| 230041000033114 | Cefixime 100mg/5ml oral suspension | Cefixime | Third-generation-cephalosporins | J01DD08 | Watch | Yes |
| 232741000033113 | Cefixime 200mg tablets | Cefixime | Third-generation-cephalosporins | J01DD08 | Watch | Yes |
| 1393841000033110 | Suprax Paediatric 100mg/5ml oral suspension | Cefixime | Third-generation-cephalosporins | J01DD08 | Watch | Yes |
| 1398741000033110 | Suprax 200mg tablets | Cefixime | Third-generation-cephalosporins | J01DD08 | Watch | Yes |
| 12395841000033100 | Cefixime 400mg tablets | Cefixime | Third-generation-cephalosporins | J01DD08 | Watch | Yes |
| 224041000033118 | Cefotaxime 1g powder for solution for injection vials | Cefotaxime sodium | Third-generation-cephalosporins | J01DD01 | Watch | Yes |
| 225741000033119 | Cefotaxime 2g powder for solution for injection vials | Cefotaxime sodium | Third-generation-cephalosporins | J01DD01 | Watch | Yes |
| 225841000033112 | Cefotaxime 500mg powder for solution for injection vials | Cefotaxime sodium | Third-generation-cephalosporins | J01DD01 | Watch | Yes |
| 275741000033118 | Claforan 1g powder for solution for injection vials | Cefotaxime sodium | Third-generation-cephalosporins | J01DD01 | Watch | Yes |
| 275841000033111 | Claforan 2g powder for solution for injection vials | Cefotaxime sodium | Third-generation-cephalosporins | J01DD01 | Watch | Yes |
| 275941000033115 | Claforan 500mg powder for solution for injection vials | Cefotaxime sodium | Third-generation-cephalosporins | J01DD01 | Watch | Yes |
| 226541000033116 | Cefoxitin 2g powder for solution for injection vials | Cefoxitin sodium | Second-generation-cephalosporins | J01DC01 | Watch | No |
| 224141000033119 | Cefoxitin 1g powder for solution for injection vials | Cefoxitin sodium | Second-generation-cephalosporins | J01DC01 | Watch | No |
| 223441000033113 | Cefrom 1g powder for solution for injection vials | Cefpirome sulfate | Fourth-generation-cephalosporins | J01DE02 | Watch | No |
| 228441000033118 | Cefpodoxime 40mg/5ml oral suspension | Cefpodoxime proxetil | Third-generation-cephalosporins | J01DD13 | Watch | No |
| 233141000033119 | Cefpodoxime 100mg tablets | Cefpodoxime proxetil | Third-generation-cephalosporins | J01DD13 | Watch | No |
| 1015141000033110 | Orelox 40mg/5ml oral suspension paediatric | Cefpodoxime proxetil | Third-generation-cephalosporins | J01DD13 | Watch | No |
| 1017041000033110 | Orelox 100mg tablets | Cefpodoxime proxetil | Third-generation-cephalosporins | J01DD13 | Watch | No |
| 1581841000033110 | Cefzil 250mg/5ml oral suspension | Cefprozil | Second-generation-cephalosporins | J01DC10 | Watch | No |
| 1581941000033110 | Cefprozil 250mg/5ml oral suspension | Cefprozil | Second-generation-cephalosporins | J01DC10 | Watch | No |
| 1582041000033110 | Cefzil 250mg tablets | Cefprozil | Second-generation-cephalosporins | J01DC10 | Watch | No |
| 1582141000033110 | Cefzil 500mg tablets | Cefprozil | Second-generation-cephalosporins | J01DC10 | Watch | No |
| 1582241000033110 | Cefprozil 250mg tablets | Cefprozil | Second-generation-cephalosporins | J01DC10 | Watch | No |
| 1582341000033110 | Cefprozil 500mg tablets | Cefprozil | Second-generation-cephalosporins | J01DC10 | Watch | No |
| 3084941000033110 | Cefradine 1g powder for solution for injection vials | Cefradine | First-generation-cephalosporins | J01DB09 | Access | No |
| 1507841000033110 | Velosef 1g powder for solution for injection vials | Cefradine | First-generation-cephalosporins | J01DB09 | Access | No |
| 1507941000033110 | Velosef 500mg powder for solution for injection vials | Cefradine | First-generation-cephalosporins | J01DB09 | Access | No |
| 3085041000033110 | Cefradine 500mg powder for solution for injection vials | Cefradine | First-generation-cephalosporins | J01DB09 | Access | No |
| 967441000033110 | Nicef 250mg capsules | Cefradine | First-generation-cephalosporins | J01DB09 | Access | No |
| 967541000033111 | Nicef 500mg capsules | Cefradine | First-generation-cephalosporins | J01DB09 | Access | No |
| 1506041000033110 | Velosef 250mg capsules | Cefradine | First-generation-cephalosporins | J01DB09 | Access | No |
| 1506141000033110 | Velosef 500mg capsules | Cefradine | First-generation-cephalosporins | J01DB09 | Access | No |
| 1511941000033110 | Velosef 250mg/5ml syrup | Cefradine | First-generation-cephalosporins | J01DB09 | Access | No |
| 3084741000033110 | Cefradine 250mg capsules | Cefradine | First-generation-cephalosporins | J01DB09 | Access | No |
| 3084841000033110 | Cefradine 500mg capsules | Cefradine | First-generation-cephalosporins | J01DB09 | Access | No |
| 3085141000033110 | Cefradine 250mg/5ml oral solution | Cefradine | First-generation-cephalosporins | J01DB09 | Access | No |
| 224341000033116 | Ceftazidime 1g powder for solution for injection vials | Ceftazidime pentahydrate | Third-generation-cephalosporins | J01DD02 | Watch | Yes |
| 1610641000033110 | Fortum Monovial 2g powder for solution for injection vials | Ceftazidime pentahydrate | Third-generation-cephalosporins | J01DD02 | Watch | Yes |
| 225941000033116 | Ceftazidime 2g powder for solution for injection vials | Ceftazidime pentahydrate | Third-generation-cephalosporins | J01DD02 | Watch | Yes |
| 226041000033114 | Ceftazidime 250mg powder for solution for injection vials | Ceftazidime pentahydrate | Third-generation-cephalosporins | J01DD02 | Watch | Yes |
| 226141000033113 | Ceftazidime 500mg powder for solution for injection vials | Ceftazidime pentahydrate | Third-generation-cephalosporins | J01DD02 | Watch | Yes |
| 600141000033115 | Fortum 1g powder for solution for injection vials | Ceftazidime pentahydrate | Third-generation-cephalosporins | J01DD02 | Watch | Yes |
| 600241000033110 | Fortum 2g powder for solution for injection vials | Ceftazidime pentahydrate | Third-generation-cephalosporins | J01DD02 | Watch | Yes |
| 600341000033117 | Fortum 250mg powder for solution for injection vials | Ceftazidime pentahydrate | Third-generation-cephalosporins | J01DD02 | Watch | Yes |
| 600441000033111 | Fortum 500mg powder for solution for injection vials | Ceftazidime pentahydrate | Third-generation-cephalosporins | J01DD02 | Watch | Yes |
| 800041000033111 | Kefadim 2g powder for solution for injection vials | Ceftazidime pentahydrate | Third-generation-cephalosporins | J01DD02 | Watch | Yes |
| 1581541000033110 | Ceftazidime 3g powder for solution for injection vials | Ceftazidime pentahydrate | Third-generation-cephalosporins | J01DD02 | Watch | Yes |
| 222741000033111 | Ceftriaxone 1g powder for solution for injection vials | Ceftriaxone sodium | Third-generation-cephalosporins | J01DD04 | Watch | Yes |
| 222841000033118 | Ceftriaxone 2g powder for solution for injection vials | Ceftriaxone sodium | Third-generation-cephalosporins | J01DD04 | Watch | Yes |
| 222941000033114 | Ceftriaxone 250mg powder for solution for injection vials | Ceftriaxone sodium | Third-generation-cephalosporins | J01DD04 | Watch | Yes |
| 1180641000033110 | Rocephin 1g powder for solution for injection vials | Ceftriaxone sodium | Third-generation-cephalosporins | J01DD04 | Watch | Yes |
| 1180741000033110 | Rocephin 2g powder for solution for injection vials | Ceftriaxone sodium | Third-generation-cephalosporins | J01DD04 | Watch | Yes |
| 1180841000033110 | Rocephin 250mg powder for solution for injection vials | Ceftriaxone sodium | Third-generation-cephalosporins | J01DD04 | Watch | Yes |
| 8349741000033110 | Cefuroxime 50mg powder for solution for injection vials | Cefuroxime sodium | Second-generation-cephalosporins | J01DC02 | Watch | Yes |
| 8349841000033110 | Aprokam 50mg powder for solution for injection vials | Cefuroxime sodium | Second-generation-cephalosporins | J01DC02 | Watch | Yes |
| 224541000033111 | Cefuroxime 1.5g powder for solution for injection vials | Cefuroxime sodium | Second-generation-cephalosporins | J01DC02 | Watch | Yes |
| 226741000033112 | Cefuroxime 250mg powder for solution for injection vials | Cefuroxime sodium | Second-generation-cephalosporins | J01DC02 | Watch | Yes |
| 226841000033119 | Cefuroxime 750mg powder for solution for injection vials | Cefuroxime sodium | Second-generation-cephalosporins | J01DC02 | Watch | Yes |
| 1554741000033110 | Zinacef 1.5g powder for solution for injection vials | Cefuroxime sodium | Second-generation-cephalosporins | J01DC02 | Watch | Yes |
| 1554841000033110 | Zinacef 250mg powder for solution for injection vials | Cefuroxime sodium | Second-generation-cephalosporins | J01DC02 | Watch | Yes |
| 1554941000033110 | Zinacef 750mg powder for solution for injection vials | Cefuroxime sodium | Second-generation-cephalosporins | J01DC02 | Watch | Yes |
| 228941000033111 | Cefuroxime 125mg granules sachets | Cefuroxime axetil | Second-generation-cephalosporins | J01DC02 | Watch | Yes |
| 230141000033113 | Cefuroxime 125mg/5ml oral suspension | Cefuroxime axetil | Second-generation-cephalosporins | J01DC02 | Watch | Yes |
| 233041000033118 | Cefuroxime 250mg tablets | Cefuroxime axetil | Second-generation-cephalosporins | J01DC02 | Watch | Yes |
| 235441000033111 | Cefuroxime 125mg tablets | Cefuroxime axetil | Second-generation-cephalosporins | J01DC02 | Watch | Yes |
| 1556241000033110 | Zinnat Suspension 125mg granules sachets | Cefuroxime axetil | Second-generation-cephalosporins | J01DC02 | Watch | Yes |
| 1557041000033110 | Zinnat 125mg/5ml oral suspension | Cefuroxime axetil | Second-generation-cephalosporins | J01DC02 | Watch | Yes |
| 1557341000033110 | Zinnat 250mg tablets | Cefuroxime axetil | Second-generation-cephalosporins | J01DC02 | Watch | Yes |
| 1558241000033110 | Zinnat 125mg tablets | Cefuroxime axetil | Second-generation-cephalosporins | J01DC02 | Watch | Yes |
| 241541000033111 | Chloramphenicol 1g powder for solution for injection vials | Chloramphenicol sodium succinate | Amphenicols | J01BA01 | Access | Yes |
| 800341000033113 | Kemicetine 1g powder for solution for injection vials | Chloramphenicol sodium succinate | Amphenicols | J01BA01 | Access | Yes |
| 237441000033116 | Chloramphenicol 250mg capsules | Chloramphenicol | Amphenicols | J01BA01 | Access | Yes |
| 253541000033110 | Ciproxin 200mg/100ml solution for infusion bottles | Ciprofloxacin lactate | Fluoroquinolones | J01MA02 | Watch | Yes |
| 253641000033111 | Ciproxin 400mg/200ml solution for infusion bottles | Ciprofloxacin lactate | Fluoroquinolones | J01MA02 | Watch | Yes |
| 253741000033119 | Ciprofloxacin 200mg/100ml solution for infusion bottles | Ciprofloxacin lactate | Fluoroquinolones | J01MA02 | Watch | Yes |
| 253841000033112 | Ciprofloxacin 400mg/200ml solution for infusion bottles | Ciprofloxacin lactate | Fluoroquinolones | J01MA02 | Watch | Yes |
| 254241000033110 | Ciprofloxacin 200mg/100ml infusion bags | Ciprofloxacin lactate | Fluoroquinolones | J01MA02 | Watch | Yes |
| 254341000033117 | Ciprofloxacin 400mg/200ml infusion bags | Ciprofloxacin lactate | Fluoroquinolones | J01MA02 | Watch | Yes |
| 254441000033111 | Ciproxin I.V. Flexibag 200mg/100ml infusion | Ciprofloxacin lactate | Fluoroquinolones | J01MA02 | Watch | Yes |
| 255341000033116 | Ciproxin 100mg/50ml solution for infusion bottles | Ciprofloxacin lactate | Fluoroquinolones | J01MA02 | Watch | Yes |
| 255641000033112 | Ciprofloxacin 100mg/50ml solution for infusion bottles | Ciprofloxacin lactate | Fluoroquinolones | J01MA02 | Watch | Yes |
| 4039741000033110 | Ciprofloxacin 200mg/100ml solution for infusion vials | Ciprofloxacin lactate | Fluoroquinolones | J01MA02 | Watch | Yes |
| 4039841000033110 | Ciprofloxacin 400mg/200ml solution for infusion vials | Ciprofloxacin lactate | Fluoroquinolones | J01MA02 | Watch | Yes |
| 5052541000033110 | Ciprofloxacin 100mg/50ml solution for infusion vials | Ciprofloxacin lactate | Fluoroquinolones | J01MA02 | Watch | Yes |
| 258741000033114 | Ciproxin 500mg tablets | Ciprofloxacin hydrochloride | Fluoroquinolones | J01MA02 | Watch | Yes |
| 258841000033116 | Ciprofloxacin 500mg tablets | Ciprofloxacin hydrochloride | Fluoroquinolones | J01MA02 | Watch | Yes |
| 258941000033112 | Ciproxin 750mg tablets | Ciprofloxacin hydrochloride | Fluoroquinolones | J01MA02 | Watch | Yes |
| 259041000033115 | Ciprofloxacin 750mg tablets | Ciprofloxacin hydrochloride | Fluoroquinolones | J01MA02 | Watch | Yes |
| 259341000033118 | Ciprofloxacin 100mg tablets | Ciprofloxacin hydrochloride | Fluoroquinolones | J01MA02 | Watch | Yes |
| 259441000033112 | Ciproxin 100mg tablets | Ciprofloxacin hydrochloride | Fluoroquinolones | J01MA02 | Watch | Yes |
| 259641000033114 | Ciproxin 250mg tablets | Ciprofloxacin hydrochloride | Fluoroquinolones | J01MA02 | Watch | Yes |
| 260241000033112 | Ciprofloxacin 250mg tablets | Ciprofloxacin hydrochloride | Fluoroquinolones | J01MA02 | Watch | Yes |
| 1582841000033110 | Ciproxin 250mg/5ml oral suspension | Ciprofloxacin | Fluoroquinolones | J01MA02 | Watch | Yes |
| 1582941000033110 | Ciprofloxacin 250mg/5ml oral suspension | Ciprofloxacin | Fluoroquinolones | J01MA02 | Watch | Yes |
| 274641000033111 | Clarithromycin 500mg powder for solution for infusion vials | Clarithromycin | Macrolides | J01FA09 | Watch | Yes |
| 804841000033112 | Klaricid IV 500mg powder for solution for infusion vials | Clarithromycin | Macrolides | J01FA09 | Watch | Yes |
| 277841000033115 | Clarithromycin 500mg modified-release tablets | Clarithromycin | Macrolides | J01FA09 | Watch | Yes |
| 278241000033118 | Clarithromycin 125mg/5ml oral suspension | Clarithromycin | Macrolides | J01FA09 | Watch | Yes |
| 282141000033118 | Clarithromycin 250mg granules sachets | Clarithromycin | Macrolides | J01FA09 | Watch | Yes |
| 285341000033110 | Clarithromycin 500mg tablets | Clarithromycin | Macrolides | J01FA09 | Watch | Yes |
| 287441000033111 | Clarithromycin 250mg tablets | Clarithromycin | Macrolides | J01FA09 | Watch | Yes |
| 804941000033116 | Klaricid XL 500mg tablets | Clarithromycin | Macrolides | J01FA09 | Watch | Yes |
| 805141000033117 | Klaricid Paediatric 125mg/5ml oral suspension | Clarithromycin | Macrolides | J01FA09 | Watch | Yes |
| 805341000033119 | Klaricid Adult 250mg granules sachets | Clarithromycin | Macrolides | J01FA09 | Watch | Yes |
| 805441000033113 | Klaricid 500 tablets | Clarithromycin | Macrolides | J01FA09 | Watch | Yes |
| 805741000033118 | Klaricid 250mg tablets | Clarithromycin | Macrolides | J01FA09 | Watch | Yes |
| 1830541000033110 | Clarithromycin 250mg/5ml oral suspension | Clarithromycin | Macrolides | J01FA09 | Watch | Yes |
| 1830641000033110 | Klaricid Paediatric 250mg/5ml oral suspension | Clarithromycin | Macrolides | J01FA09 | Watch | Yes |
| 3838541000033110 | Clarithromycin 125mg granules straws | Clarithromycin | Macrolides | J01FA09 | Watch | Yes |
| 3838641000033110 | Clarithromycin 187.5mg granules straws | Clarithromycin | Macrolides | J01FA09 | Watch | Yes |
| 3838741000033110 | Clarithromycin 250mg granules straws | Clarithromycin | Macrolides | J01FA09 | Watch | Yes |
| 3838841000033110 | Clarosip 125mg granules for oral suspension straws | Clarithromycin | Macrolides | J01FA09 | Watch | Yes |
| 3838941000033110 | Clarosip 187.5mg granules for oral suspension straws | Clarithromycin | Macrolides | J01FA09 | Watch | Yes |
| 3839041000033110 | Clarosip 250mg granules for oral suspension straws | Clarithromycin | Macrolides | J01FA09 | Watch | Yes |
| 6030241000033110 | Clarie XL 500mg tablets | Clarithromycin | Macrolides | J01FA09 | Watch | Yes |
| 3873041000033110 | Clindamycin 300mg/2ml solution for injection ampoules | Clindamycin phosphate | Lincosamides | J01FF01 | Access | Yes |
| 3873141000033110 | Clindamycin 600mg/4ml solution for injection ampoules | Clindamycin phosphate | Lincosamides | J01FF01 | Access | Yes |
| 13419541000033100 | Clindamycin 300mg/50ml infusion bags | Clindamycin phosphate | Lincosamides | J01FF01 | Access | Yes |
| 13419641000033100 | Clindamycin 600mg/50ml infusion bags | Clindamycin phosphate | Lincosamides | J01FF01 | Access | Yes |
| 3873241000033110 | Dalacin C Phosphate 300mg/2ml solution for injection ampoules | Clindamycin phosphate | Lincosamides | J01FF01 | Access | Yes |
| 3873341000033110 | Dalacin C Phosphate 600mg/4ml solution for injection ampoules | Clindamycin phosphate | Lincosamides | J01FF01 | Access | Yes |
| 261541000033115 | Clindamycin 150mg capsules | Clindamycin hydrochloride | Lincosamides | J01FF01 | Access | Yes |
| 262741000033117 | Clindamycin 75mg capsules | Clindamycin hydrochloride | Lincosamides | J01FF01 | Access | Yes |
| 396041000033114 | Dalacin C 150mg capsules | Clindamycin hydrochloride | Lincosamides | J01FF01 | Access | Yes |
| 396141000033113 | Dalacin C 75mg capsules | Clindamycin hydrochloride | Lincosamides | J01FF01 | Access | Yes |
| 8243241000033110 | Clindamycin 300mg capsules | Clindamycin hydrochloride | Lincosamides | J01FF01 | Access | Yes |
| 8495241000033110 | Clindamycin 75mg/5ml oral suspension | Clindamycin hydrochloride | Lincosamides | J01FF01 | Access | Yes |
| 368641000033119 | Colomycin 250,000units/5ml syrup | Colistin sulfate | Polymyxins | A07AA10 | Reserve | No |
| 368841000033118 | Colistin 250,000units/5ml oral solution | Colistin sulfate | Polymyxins | A07AA10 | Reserve | No |
| 371541000033119 | Colomycin 1.5million unit tablets | Colistin sulfate | Polymyxins | A07AA10 | Reserve | No |
| 372641000033113 | Colistin 1.5million unit tablets | Colistin sulfate | Polymyxins | A07AA10 | Reserve | No |
| 12208041000033100 | Dalbavancin 500mg powder for solution for infusion vials | Dalbavancin hydrochloride | Glycopeptides | J01XA04 | Reserve | No |
| 3892141000033110 | Daptomycin 350mg powder for solution for infusion vials | Daptomycin | Lipopeptides | J01XX09 | Reserve | No |
| 3892241000033110 | Cubicin 350mg powder for concentrate for solution for infusion vials | Daptomycin | Lipopeptides | J01XX09 | Reserve | No |
| 3993741000033110 | Daptomycin 500mg powder for solution for infusion vials | Daptomycin | Lipopeptides | J01XX09 | Reserve | No |
| 13722841000033100 | Delafloxacin 450mg tablets | Delafloxacin meglumine | Fluoroquinolones | J01MA23 | Watch | No |
| 416041000033117 | Demeclocycline 150mg capsules | Demeclocycline hydrochloride | Tetracyclines | J01AA01 | Watch | No |
| 820141000033117 | Ledermycin 150mg capsules | Demeclocycline hydrochloride | Tetracyclines | J01AA01 | Watch | No |
| 9702841000033110 | Demeclocycline 150mg tablets | Demeclocycline hydrochloride | Tetracyclines | J01AA01 | Watch | No |
| 415841000033119 | Demix 100 capsules | Doxycycline hyclate | Tetracyclines | J01AA02 | Access | Yes |
| 415941000033110 | Demix 50 capsules | Doxycycline hyclate | Tetracyclines | J01AA02 | Access | Yes |
| 1514941000033110 | Vibramycin 100mg capsules | Doxycycline hyclate | Tetracyclines | J01AA02 | Access | Yes |
| 1515041000033110 | Vibramycin 50 capsules | Doxycycline hyclate | Tetracyclines | J01AA02 | Access | Yes |
| 1523841000033110 | Vibramycin-D 100mg dispersible tablets | Doxycycline monohydrate | Tetracyclines | J01AA02 | Access | Yes |
| 1603241000033110 | Doxycycline 50mg capsules | Doxycycline hyclate | Tetracyclines | J01AA02 | Access | Yes |
| 1705841000033110 | Vibramycin Acne Pack 50mg capsules | Doxycycline hyclate | Tetracyclines | J01AA02 | Access | Yes |
| 2104241000033110 | Doxylar 50mg capsules | Doxycycline hyclate | Tetracyclines | J01AA02 | Access | Yes |
| 2104341000033110 | Doxylar 100mg capsules | Doxycycline hyclate | Tetracyclines | J01AA02 | Access | Yes |
| 2193041000033110 | Vibrox 100mg capsules | Doxycycline hyclate | Tetracyclines | J01AA02 | Access | Yes |
| 2637141000033110 | Periostat 20mg tablets | Doxycycline hyclate | Tetracyclines | J01AA02 | Access | Yes |
| 3089341000033110 | Doxycycline 100mg capsules | Doxycycline hyclate | Tetracyclines | J01AA02 | Access | Yes |
| 3089441000033110 | Doxycycline 100mg dispersible tablets sugar free | Doxycycline monohydrate | Tetracyclines | J01AA02 | Access | Yes |
| 3089541000033110 | Doxycycline 20mg tablets | Doxycycline hyclate | Tetracyclines | J01AA02 | Access | Yes |
| 5150141000033110 | Doxycycline 40mg modified-release capsules | Doxycycline monohydrate | Tetracyclines | J01AA02 | Access | Yes |
| 5150241000033110 | Efracea 40mg modified-release capsules | Doxycycline monohydrate | Tetracyclines | J01AA02 | Access | Yes |
| 7871241000033110 | Doxycycline 50mg/5ml oral suspension | Doxycycline hyclate | Tetracyclines | J01AA02 | Access | Yes |
| 2839241000033110 | Ertapenem 1g powder for solution for infusion vials | Ertapenem sodium | Carbapenems | J01DH03 | Watch | No |
| 2839341000033110 | Invanz 1g powder for solution for infusion vials | Ertapenem sodium | Carbapenems | J01DH03 | Watch | No |
| 535241000033119 | Erythrocin IV lactobionate 1g powder for solution for infusion vials | Erythromycin lactobionate | Macrolides | J01FA01 | Watch | No |
| 535441000033118 | Erythromycin 1g powder for solution for infusion vials | Erythromycin lactobionate | Macrolides | J01FA01 | Watch | No |
| 534141000033113 | Erymax 250mg gastro-resistant capsules | Erythromycin | Macrolides | J01FA01 | Watch | No |
| 534241000033118 | Erythromycin 250mg gastro-resistant capsules | Erythromycin | Macrolides | J01FA01 | Watch | No |
| 536141000033119 | Erythroped PI SF 125mg/5ml oral suspension | Erythromycin ethyl succinate | Macrolides | J01FA01 | Watch | No |
| 536941000033117 | Erythromycin stearate 250mg tablets | Erythromycin stearate | Macrolides | J01FA01 | Watch | No |
| 537241000033112 | Erythroped Forte SF 500mg/5ml oral suspension | Erythromycin ethyl succinate | Macrolides | J01FA01 | Watch | No |
| 537341000033119 | Erythroped SF 250mg/5ml oral suspension | Erythromycin ethyl succinate | Macrolides | J01FA01 | Watch | No |
| 537441000033113 | Erythromycin ethyl succinate 125mg/5ml oral suspension sugar free | Erythromycin ethyl succinate | Macrolides | J01FA01 | Watch | No |
| 537541000033114 | Erythromycin ethyl succinate 250mg/5ml oral suspension sugar free | Erythromycin ethyl succinate | Macrolides | J01FA01 | Watch | No |
| 537641000033110 | Erythromycin ethyl succinate 125mg/5ml oral suspension | Erythromycin ethyl succinate | Macrolides | J01FA01 | Watch | No |
| 537741000033118 | Erythromycin ethyl succinate 250mg/5ml oral suspension | Erythromycin ethyl succinate | Macrolides | J01FA01 | Watch | No |
| 537841000033111 | Erythromycin ethyl succinate 500mg/5ml oral suspension | Erythromycin ethyl succinate | Macrolides | J01FA01 | Watch | No |
| 537941000033115 | Erythromycin ethyl succinate 500mg/5ml oral suspension sugar free | Erythromycin ethyl succinate | Macrolides | J01FA01 | Watch | No |
| 538841000033112 | Erythromycin ethyl succinate 500mg tablets | Erythromycin ethyl succinate | Macrolides | J01FA01 | Watch | No |
| 539541000033115 | Erythrocin 250 tablets | Erythromycin stearate | Macrolides | J01FA01 | Watch | No |
| 539641000033119 | Erythrocin 500 tablets | Erythromycin stearate | Macrolides | J01FA01 | Watch | No |
| 540141000033115 | Erythroped A 500mg tablets | Erythromycin ethyl succinate | Macrolides | J01FA01 | Watch | No |
| 1183741000033110 | Rommix 250 EC tablets | Erythromycin | Macrolides | J01FA01 | Watch | No |
| 1436941000033110 | Tiloryth 250mg gastro-resistant capsules | Erythromycin | Macrolides | J01FA01 | Watch | No |
| 1924941000033110 | Erythromycin 250mg gastro-resistant tablets | Erythromycin | Macrolides | J01FA01 | Watch | No |
| 2890541000033110 | Kerymax 250mg gastro-resistant capsules | Erythromycin | Macrolides | J01FA01 | Watch | No |
| 13300741000033100 | Erythromycin stearate 500mg tablets | Erythromycin stearate | Macrolides | J01FA01 | Watch | No |
| 584241000033113 | Floxapen 1g powder for solution for injection vials | Flucloxacillin sodium | Penicillins | J01CF05 | Access | No |
| 584341000033115 | Floxapen 250mg powder for solution for injection vials | Flucloxacillin sodium | Penicillins | J01CF05 | Access | No |
| 584441000033114 | Floxapen 500mg powder for solution for injection vials | Flucloxacillin sodium | Penicillins | J01CF05 | Access | No |
| 587941000033111 | Flucloxacillin 250mg powder for solution for injection vials | Flucloxacillin sodium | Penicillins | J01CF05 | Access | No |
| 12395741000033100 | Flucloxacillin 2g powder for solution for injection vials | Flucloxacillin sodium | Penicillins | J01CF05 | Access | No |
| 587341000033112 | Flucloxacillin 1g powder for solution for injection vials | Flucloxacillin sodium | Penicillins | J01CF05 | Access | No |
| 587841000033115 | Flucloxacillin 500mg powder for solution for injection vials | Flucloxacillin sodium | Penicillins | J01CF05 | Access | No |
| 577941000033118 | Floxapen 250mg capsules | Flucloxacillin sodium | Penicillins | J01CF05 | Access | No |
| 578041000033115 | Floxapen 500mg capsules | Flucloxacillin sodium | Penicillins | J01CF05 | Access | No |
| 578141000033116 | Flucloxacillin 250mg capsules | Flucloxacillin sodium | Penicillins | J01CF05 | Access | No |
| 578241000033111 | Flucloxacillin 500mg capsules | Flucloxacillin sodium | Penicillins | J01CF05 | Access | No |
| 579541000033118 | Fluclomix 500 capsules | Flucloxacillin sodium | Penicillins | J01CF05 | Access | No |
| 579741000033114 | Fluclomix 250 capsules | Flucloxacillin sodium | Penicillins | J01CF05 | Access | No |
| 590241000033119 | Flucloxacillin 125mg/5ml oral solution | Flucloxacillin sodium | Penicillins | J01CF05 | Access | No |
| 590441000033118 | Flucloxacillin 250mg/5ml oral suspension | Flucloxacillin magnesium | Penicillins | J01CF05 | Access | No |
| 592341000033113 | Floxapen 125mg/5ml syrup | Flucloxacillin magnesium | Penicillins | J01CF05 | Access | No |
| 592441000033119 | Floxapen 250mg/5ml syrup | Flucloxacillin magnesium | Penicillins | J01CF05 | Access | No |
| 808441000033117 | Ladropen 250mg capsules | Flucloxacillin sodium | Penicillins | J01CF05 | Access | No |
| 808541000033116 | Ladropen 500mg capsules | Flucloxacillin sodium | Penicillins | J01CF05 | Access | No |
| 4086141000033110 | Flucloxacillin 250mg/5ml oral solution | Flucloxacillin sodium | Penicillins | J01CF05 | Access | No |
| 5292941000033110 | Flucloxacillin 125mg/5ml oral solution sugar free | Flucloxacillin sodium | Penicillins | J01CF05 | Access | No |
| 5293041000033110 | Flucloxacillin 250mg/5ml oral solution sugar free | Flucloxacillin sodium | Penicillins | J01CF05 | Access | No |
| 9302641000033110 | Fosfomycin 2g powder for solution for infusion vials | Fosfomycin sodium | Phosphonics | J01XX01 | Reserve | Yes |
| 9995241000033110 | Fosfomycin 4g powder for solution for infusion vials | Fosfomycin sodium | Phosphonics | J01XX01 | Reserve | Yes |
| 599941000033116 | Fosfomycin 3g granules sachets | Fosfomycin trometamol | Phosphonics | J01XX01 | Watch | No |
| 925241000033114 | Monuril 3g granules sachets | Fosfomycin trometamol | Phosphonics | J01XX01 | Watch | No |
| 8946441000033110 | Fosfomycin 500mg capsules | Fosfomycin calcium | Phosphonics | J01XX01 | Watch | No |
| 618641000033113 | Fucidin 250mg/5ml oral suspension | Fusidic acid | Steroid antibacterials | J01XC01 | Watch | No |
| 618841000033114 | Fusidic acid 250mg/5ml oral suspension | Fusidic acid | Steroid antibacterials | J01XC01 | Watch | No |
| 11524841000033100 | Levofloxacin 100mg/ml nebuliser liquid ampoules | Levofloxacin hemihydrate | Fluoroquinolones | J01MA12 | Watch | No |
| 11525041000033100 | Quinsair 240mg nebuliser solution ampoules | Levofloxacin hemihydrate | Fluoroquinolones | J01MA12 | Watch | No |
| 1626141000033110 | Levofloxacin 500mg/100ml solution for infusion vials | Levofloxacin hemihydrate | Fluoroquinolones | J01MA12 | Watch | No |
| 1698641000033110 | Tavanic 500mg/100ml solution for infusion vials | Levofloxacin hemihydrate | Fluoroquinolones | J01MA12 | Watch | No |
| 6427241000033110 | Levofloxacin 500mg/100ml infusion bags | Levofloxacin hemihydrate | Fluoroquinolones | J01MA12 | Watch | No |
| 8618641000033110 | Levofloxacin 500mg/100ml solution for infusion bottles | Levofloxacin hemihydrate | Fluoroquinolones | J01MA12 | Watch | No |
| 1626241000033110 | Levofloxacin 250mg tablets | Levofloxacin hemihydrate | Fluoroquinolones | J01MA12 | Watch | No |
| 1626341000033110 | Levofloxacin 500mg tablets | Levofloxacin hemihydrate | Fluoroquinolones | J01MA12 | Watch | No |
| 1698741000033110 | Tavanic 250mg tablets | Levofloxacin hemihydrate | Fluoroquinolones | J01MA12 | Watch | No |
| 1698841000033110 | Tavanic 500mg tablets | Levofloxacin hemihydrate | Fluoroquinolones | J01MA12 | Watch | No |
| 10220241000033100 | Evoxil 500mg tablets | Levofloxacin hemihydrate | Fluoroquinolones | J01MA12 | Watch | No |
| 854441000033117 | Lymecycline 408mg capsules | Lymecycline | Tetracyclines | J01AA04 | Watch | No |
| 1414641000033110 | Tetralysal 300 capsules | Lymecycline | Tetracyclines | J01AA04 | Watch | No |
| 882841000033116 | Meropenem 1g powder for solution for injection vials | Meropenem trihydrate | Carbapenems | J01DH02 | Watch | Yes |
| 883041000033119 | Meropenem 500mg powder for solution for injection vials | Meropenem trihydrate | Carbapenems | J01DH02 | Watch | Yes |
| 883441000033111 | Meronem 1g powder for solution for injection vials | Meropenem trihydrate | Carbapenems | J01DH02 | Watch | Yes |
| 883641000033113 | Meronem 500mg powder for solution for injection vials | Meropenem trihydrate | Carbapenems | J01DH02 | Watch | Yes |
| 2923441000033110 | Metronidazole 100mg/20ml solution for infusion ampoules | Metronidazole | Imidazoles | J01XD01 | Access | Yes |
| 2923541000033110 | Metronidazole 500mg/100ml infusion 100ml bags | Metronidazole | Imidazoles | J01XD01 | Access | Yes |
| 2923641000033110 | Flagyl 500mg/100ml infusion 100ml bags | Metronidazole | Imidazoles | J01XD01 | Access | Yes |
| 12356041000033100 | Metronidazole 500mg/100ml solution for infusion bottles | Metronidazole | Imidazoles | J01XD01 | Access | Yes |
| 592041000033111 | Flagyl-S 200mg/5ml oral suspension | Metronidazole benzoate | Imidazoles | P01AB01 | Access | Yes |
| 593041000033119 | Flagyl 200mg tablets | Metronidazole | Imidazoles | P01AB01 | Access | Yes |
| 593141000033115 | Flagyl 400mg tablets | Metronidazole | Imidazoles | P01AB01 | Access | Yes |
| 891941000033116 | Metronidazole 100mg/5ml oral suspension | Metronidazole benzoate | Imidazoles | P01AB01 | Access | Yes |
| 893141000033119 | Metronidazole 200mg/5ml oral suspension | Metronidazole benzoate | Imidazoles | P01AB01 | Access | Yes |
| 897941000033114 | Metronidazole 200mg tablets | Metronidazole | Imidazoles | P01AB01 | Access | Yes |
| 898041000033112 | Metronidazole 400mg tablets | Metronidazole | Imidazoles | P01AB01 | Access | Yes |
| 899441000033113 | Metronidazole 500mg tablets | Metronidazole | Imidazoles | P01AB01 | Access | Yes |
| 591941000033117 | Flagyl 500mg suppositories | Metronidazole | Imidazoles | J01XD01 | Access | Yes |
| 592241000033115 | Flagyl 1g suppositories | Metronidazole | Imidazoles | J01XD01 | Access | Yes |
| 892441000033118 | Metronidazole 500mg suppositories | Metronidazole | Imidazoles | J01XD01 | Access | Yes |
| 892741000033113 | Metrolyl 1g suppositories | Metronidazole | Imidazoles | J01XD01 | Access | Yes |
| 892841000033115 | Metrolyl 500mg suppositories | Metronidazole | Imidazoles | J01XD01 | Access | Yes |
| 893041000033118 | Metronidazole 1g suppositories | Metronidazole | Imidazoles | J01XD01 | Access | Yes |
| 23141000033112 | Aknemin 100mg capsules | Minocycline hydrochloride | Tetracyclines | J01AA08 | Watch | No |
| 23241000033117 | Aknemin 50 capsules | Minocycline hydrochloride | Tetracyclines | J01AA08 | Watch | No |
| 153241000033111 | Blemix 50mg tablets | Minocycline hydrochloride | Tetracyclines | J01AA08 | Watch | No |
| 153341000033118 | Blemix 100mg tablets | Minocycline hydrochloride | Tetracyclines | J01AA08 | Watch | No |
| 906241000033112 | Minocycline 100mg capsules | Minocycline hydrochloride | Tetracyclines | J01AA08 | Watch | No |
| 906341000033119 | Minocycline 50mg capsules | Minocycline hydrochloride | Tetracyclines | J01AA08 | Watch | No |
| 916041000033119 | Minocin MR 100mg capsules | Minocycline hydrochloride | Tetracyclines | J01AA08 | Watch | No |
| 916141000033115 | Minocycline 100mg modified-release capsules | Minocycline hydrochloride | Tetracyclines | J01AA08 | Watch | No |
| 918641000033112 | Minocin 100mg tablets | Minocycline hydrochloride | Tetracyclines | J01AA08 | Watch | No |
| 918741000033115 | Minocin 50mg tablets | Minocycline hydrochloride | Tetracyclines | J01AA08 | Watch | No |
| 918841000033113 | Minocycline 100mg tablets | Minocycline hydrochloride | Tetracyclines | J01AA08 | Watch | No |
| 918941000033117 | Minocycline 50mg tablets | Minocycline hydrochloride | Tetracyclines | J01AA08 | Watch | No |
| 2993941000033110 | Sebomin MR 100mg capsules | Minocycline hydrochloride | Tetracyclines | J01AA08 | Watch | No |
| 3855741000033110 | Sebren MR 100mg capsules | Minocycline hydrochloride | Tetracyclines | J01AA08 | Watch | No |
| 3980541000033110 | Acnamino MR 100mg capsules | Minocycline hydrochloride | Tetracyclines | J01AA08 | Watch | No |
| 5454241000033110 | Moxifloxacin 400mg/250ml solution for infusion bottles | Moxifloxacin hydrochloride | Fluoroquinolones | J01MA14 | Watch | No |
| 5454341000033110 | Avelox 400mg/250ml solution for infusion bottles | Moxifloxacin hydrochloride | Fluoroquinolones | J01MA14 | Watch | No |
| 2890241000033110 | Moxifloxacin 400mg tablets | Moxifloxacin hydrochloride | Fluoroquinolones | J01MA14 | Watch | No |
| 2890341000033110 | Avelox 400mg tablets | Moxifloxacin hydrochloride | Fluoroquinolones | J01MA14 | Watch | No |
| 953941000033117 | Nalidixic acid 300mg/5ml oral suspension | Nalidixic acid | Quinolones | J01MB02 | Watch | No |
| 954741000033117 | Nalidixic acid 500mg tablets | Nalidixic acid | Quinolones | J01MB02 | Watch | No |
| 964941000033112 | Negram 300mg/5ml oral suspension | Nalidixic acid | Quinolones | J01MB02 | Watch | No |
| 965141000033111 | Negram 500mg tablets | Nalidixic acid | Quinolones | J01MB02 | Watch | No |
| 1494941000033110 | Uriben 300mg/5ml oral suspension | Nalidixic acid | Quinolones | J01MB02 | Watch | No |
| 976441000033117 | Nivemycin 500mg tablets | Neomycin sulfate | Aminoglycosides | A07AA01 | Watch | No |
| 1659541000033110 | Neomycin 500mg tablets | Neomycin sulfate | Aminoglycosides | A07AA01 | Watch | No |
| 618541000033112 | Furadantin 25mg/5ml oral suspension | Nitrofurantoin | Nitrofuran-derivatives | J01XE01 | Access | Yes |
| 619141000033114 | Furadantin 100mg tablets | Nitrofurantoin | Nitrofuran-derivatives | J01XE01 | Access | Yes |
| 619241000033119 | Furadantin 50mg tablets | Nitrofurantoin | Nitrofuran-derivatives | J01XE01 | Access | Yes |
| 857841000033111 | Macrodantin 100mg capsules | Nitrofurantoin | Nitrofuran-derivatives | J01XE01 | Access | Yes |
| 857941000033115 | Macrodantin 50mg capsules | Nitrofurantoin | Nitrofuran-derivatives | J01XE01 | Access | Yes |
| 863441000033113 | Macrobid 100mg modified-release capsules | Nitrofurantoin | Nitrofuran-derivatives | J01XE01 | Access | Yes |
| 967941000033117 | Nitrofurantoin 100mg capsules | Nitrofurantoin | Nitrofuran-derivatives | J01XE01 | Access | Yes |
| 968141000033115 | Nitrofurantoin 50mg capsules | Nitrofurantoin | Nitrofuran-derivatives | J01XE01 | Access | Yes |
| 971641000033114 | Nitrofurantoin 100mg modified-release capsules | Nitrofurantoin | Nitrofuran-derivatives | J01XE01 | Access | Yes |
| 974441000033110 | Nitrofurantoin 25mg/5ml oral suspension sugar free | Nitrofurantoin | Nitrofuran-derivatives | J01XE01 | Access | Yes |
| 975441000033114 | Nitrofurantoin 100mg tablets | Nitrofurantoin | Nitrofuran-derivatives | J01XE01 | Access | Yes |
| 975541000033110 | Nitrofurantoin 50mg tablets | Nitrofurantoin | Nitrofuran-derivatives | J01XE01 | Access | Yes |
| 1495641000033110 | Urantoin 50mg tablets | Nitrofurantoin | Nitrofuran-derivatives | J01XE01 | Access | Yes |
| 2955441000033110 | Urantoin 100mg tablets | Nitrofurantoin | Nitrofuran-derivatives | J01XE01 | Access | Yes |
| 5997141000033110 | Nitrofurantoin 25mg/5ml oral solution | Nitrofurantoin | Nitrofuran-derivatives | J01XE01 | Access | Yes |
| 5997241000033110 | Nitrofurantoin 25mg/5ml oral suspension | Nitrofurantoin | Nitrofuran-derivatives | J01XE01 | Access | Yes |
| 11603941000033100 | Genfura 100mg tablets | Nitrofurantoin | Nitrofuran-derivatives | J01XE01 | Access | Yes |
| 11604041000033100 | Genfura 50mg tablets | Nitrofurantoin | Nitrofuran-derivatives | J01XE01 | Access | Yes |
| 986441000033114 | Norfloxacin 400mg tablets | Norfloxacin | Fluoroquinolones | J01MA06 | Watch | No |
| 1496041000033110 | Utinor 400mg tablets | Norfloxacin | Fluoroquinolones | J01MA06 | Watch | No |
| 9290241000033110 | Ofloxacin 200mg/100ml solution for infusion bottles | Ofloxacin hydrochloride | Fluoroquinolones | J01MA01 | Watch | No |
| 1002541000033110 | Ofloxacin 200mg tablets | Ofloxacin | Fluoroquinolones | J01MA01 | Watch | No |
| 1002641000033110 | Ofloxacin 400mg tablets | Ofloxacin | Fluoroquinolones | J01MA01 | Watch | No |
| 1407541000033110 | Tarivid 400mg tablets | Ofloxacin | Fluoroquinolones | J01MA01 | Watch | No |
| 1409341000033110 | Tarivid 200mg tablets | Ofloxacin | Fluoroquinolones | J01MA01 | Watch | No |
| 1025441000033110 | Oxytetramix 250 tablets | Oxytetracycline dihydrate | Tetracyclines | J01AA06 | Watch | No |
| 1026541000033110 | Oxytetracycline 250mg tablets | Oxytetracycline dihydrate | Tetracyclines | J01AA06 | Watch | No |
| 1026741000033110 | Oxymycin 250mg tablets | Oxytetracycline dihydrate | Tetracyclines | J01AA06 | Watch | No |
| 1427741000033110 | Terramycin 250mg tablets | Oxytetracycline dihydrate | Tetracyclines | J01AA06 | Watch | No |
| 3267441000033110 | Paromomycin 125mg/5ml oral solution | Paromomycin sulfate | Aminoglycosides | A07AA06 | Unclassified | No |
| 8495141000033110 | Paromomycin 250mg tablets | Paromomycin sulfate | Aminoglycosides | A07AA06 | Unclassified | No |
| 1076141000033110 | Phenoxymethylpenicillin 125mg/5ml oral solution | Phenoxymethylpenicillin potassium | Penicillins | J01CE02 | Access | Yes |
| 1076241000033110 | Phenoxymethylpenicillin 250mg/5ml oral solution | Phenoxymethylpenicillin potassium | Penicillins | J01CE02 | Access | Yes |
| 1081941000033110 | Phenoxymethylpenicillin 250mg tablets | Phenoxymethylpenicillin potassium | Penicillins | J01CE02 | Access | Yes |
| 2130541000033110 | Tenkicin 250mg tablets | Phenoxymethylpenicillin potassium | Penicillins | J01CE02 | Access | Yes |
| 3923541000033110 | Phenoxymethylpenicillin 125mg/5ml oral solution sugar free | Phenoxymethylpenicillin potassium | Penicillins | J01CE02 | Access | Yes |
| 3923641000033110 | Phenoxymethylpenicillin 250mg/5ml oral solution sugar free | Phenoxymethylpenicillin potassium | Penicillins | J01CE02 | Access | Yes |
| 1091741000033110 | Pivmecillinam 200mg tablets | Pivmecillinam hydrochloride | Penicillins | J01CA08 | Access | No |
| 1275241000033110 | Selexid 200mg tablets | Pivmecillinam hydrochloride | Penicillins | J01CA08 | Access | No |
| 2746541000033110 | Pristinamycin 500mg tablets | Pristinamycin | Streptogramins | J01FG01 | Watch | No |
| 3105141000033110 | Sulfadiazine 1g/4ml solution for injection ampoules | Sodium sulfadiazine | Sulfonamides | J01EC02 | Access | No |
| 1370641000033110 | Spectinomycin 2g powder and solvent for suspension for injection vials | Spectinomycin hydrochloride | Aminocyclitols | J01XX04 | Access | Yes |
| 9302541000033110 | Streptomycin 1g powder for solution for injection vials | Streptomycin sulfate | Aminoglycosides | J01GA01 | Watch | No |
| 3105241000033110 | Sulfadiazine 500mg tablets | Sulfadiazine | Sulfonamides | J01EC02 | Access | No |
| 5975741000033110 | Sulfadiazine 250mg/5ml oral suspension | Sulfadiazine | Sulfonamides | J01EC02 | Access | No |
| 3104641000033110 | Sulfamethoxypyridazine 500mg tablets | Sulfamethoxypyridazine | Sulfonamides | J01ED05 | Access | No |
| 6035241000033110 | Sulfapyridine 250mg capsules | Sulfapyridine | Sulfonamides | J01EB04 | Access | No |
| 1420841000033110 | Teicoplanin 200mg powder and solvent for solution for injection vials | Teicoplanin | Glycopeptides | J01XA02 | Watch | No |
| 1420941000033110 | Teicoplanin 400mg powder and solvent for solution for injection vials | Teicoplanin | Glycopeptides | J01XA02 | Watch | No |
| 1406741000033110 | Targocid 200mg powder and solvent for solution for injection vials | Teicoplanin | Glycopeptides | J01XA02 | Watch | No |
| 1406841000033110 | Targocid 400mg powder and solvent for solution for injection vials | Teicoplanin | Glycopeptides | J01XA02 | Watch | No |
| 2676841000033110 | Telithromycin 400mg tablets | Telithromycin | Macrolides | J01FA15 | Watch | No |
| 2676941000033110 | Ketek 400mg tablets | Telithromycin | Macrolides | J01FA15 | Watch | No |
| 1421041000033110 | Temocillin 1g powder for solution for injection vials | Temocillin sodium | Penicillins | J01CA17 | Watch | No |
| 3541000033113 | Achromycin 250mg capsules | Tetracycline hydrochloride | Tetracyclines | J01AA07 | Access | No |
| 1414841000033110 | Tetracycline 250mg capsules | Tetracycline hydrochloride | Tetracyclines | J01AA07 | Access | No |
| 1428041000033110 | Tetracycline 250mg tablets | Tetracycline hydrochloride | Tetracyclines | J01AA07 | Access | No |
| 3923141000033110 | Tigecycline 50mg powder for solution for infusion vials | Tigecycline | Glycylcyclines | J01AA12 | Reserve | No |
| 3923241000033110 | Tygacil 50mg powder for solution for infusion vials | Tigecycline | Glycylcyclines | J01AA12 | Reserve | No |
| 563241000033113 | Fasigyn 500mg tablets | Tinidazole | Imidazoles | P01AB02 | Access | No |
| 1444641000033110 | Tinidazole 500mg tablets | Tinidazole | Imidazoles | P01AB02 | Access | No |
| 1924741000033110 | Tobramycin 300mg/5ml nebuliser liquid ampoules | Tobramycin | Aminoglycosides | J01GB01 | Watch | No |
| 1924841000033110 | Tobi 300mg/5ml nebuliser solution 5ml ampoules | Tobramycin | Aminoglycosides | J01GB01 | Watch | No |
| 4898541000033110 | Tobramycin 300mg/4ml nebuliser liquid ampoules | Tobramycin | Aminoglycosides | J01GB01 | Watch | No |
| 4898641000033110 | Bramitob 300mg/4ml nebuliser solution 4ml ampoules | Tobramycin | Aminoglycosides | J01GB01 | Watch | No |
| 6454141000033110 | Tobramycin 28mg inhalation powder capsules with device | Tobramycin | Aminoglycosides | J01GB01 | Watch | No |
| 6454241000033110 | Tobi Podhaler 28mg inhalation powder capsules with device | Tobramycin | Aminoglycosides | J01GB01 | Watch | No |
| 9118041000033110 | Tymbrineb 300mg/5ml nebuliser solution 5ml ampoules | Tobramycin | Aminoglycosides | J01GB01 | Watch | No |
| 11717241000033100 | Tobramycin 170mg/1.7ml nebuliser liquid ampoules | Tobramycin | Aminoglycosides | J01GB01 | Watch | No |
| 9730341000033110 | Tobramycin 240mg/6ml solution for injection vials | Tobramycin | Aminoglycosides | J01GB01 | Watch | No |
| 961341000033111 | Nebcin 80mg/2ml solution for injection vials | Tobramycin | Aminoglycosides | J01GB01 | Watch | No |
| 961941000033110 | Nebcin 20mg/2ml solution for injection vials | Tobramycin sulfate | Aminoglycosides | J01GB01 | Watch | No |
| 1446541000033110 | Tobramycin 80mg/2ml solution for injection vials | Tobramycin | Aminoglycosides | J01GB01 | Watch | No |
| 1447041000033110 | Tobramycin 20mg/2ml solution for injection vials | Tobramycin sulfate | Aminoglycosides | J01GB01 | Watch | No |
| 1447141000033110 | Tobramycin 40mg/1ml solution for injection vials | Tobramycin sulfate | Aminoglycosides | J01GB01 | Watch | No |
| 928541000033118 | Monotrim 100mg/5ml solution for injection ampoules | Trimethoprim | Trimethoprim-derivatives | J01EA01 | Access | Yes |
| 1460641000033110 | Trimethoprim 100mg/5ml solution for injection ampoules | Trimethoprim | Trimethoprim-derivatives | J01EA01 | Access | Yes |
| 933741000033116 | Monotrim 50mg/5ml oral suspension | Trimethoprim | Trimethoprim-derivatives | J01EA01 | Access | Yes |
| 936041000033112 | Monotrim 100mg tablets | Trimethoprim | Trimethoprim-derivatives | J01EA01 | Access | Yes |
| 936141000033111 | Monotrim 200mg tablets | Trimethoprim | Trimethoprim-derivatives | J01EA01 | Access | Yes |
| 1467541000033110 | Trimethoprim 50mg/5ml oral suspension sugar free | Trimethoprim | Trimethoprim-derivatives | J01EA01 | Access | Yes |
| 1471141000033110 | Trimethoprim 100mg tablets | Trimethoprim | Trimethoprim-derivatives | J01EA01 | Access | Yes |
| 1471241000033110 | Trimethoprim 200mg tablets | Trimethoprim | Trimethoprim-derivatives | J01EA01 | Access | Yes |
| 1472641000033110 | Trimogal 100mg tablets | Trimethoprim | Trimethoprim-derivatives | J01EA01 | Access | Yes |
| 1472741000033110 | Trimogal 200mg tablets | Trimethoprim | Trimethoprim-derivatives | J01EA01 | Access | Yes |
| 1472841000033110 | Trimopan 100mg tablets | Trimethoprim | Trimethoprim-derivatives | J01EA01 | Access | Yes |
| 1472941000033110 | Trimopan 200mg tablets | Trimethoprim | Trimethoprim-derivatives | J01EA01 | Access | Yes |
| 1501541000033110 | Vancomycin 1g powder for solution for infusion vials | Vancomycin hydrochloride | Glycopeptides | A07AA09 | Watch | Yes |
| 1498741000033110 | Vancocin Matrigel 250mg capsules | Vancomycin hydrochloride | Glycopeptides | A07AA09 | Watch | Yes |
| 1498841000033110 | Vancomycin 125mg capsules | Vancomycin hydrochloride | Glycopeptides | A07AA09 | Watch | Yes |
| 1498941000033110 | Vancomycin 250mg capsules | Vancomycin hydrochloride | Glycopeptides | A07AA09 | Watch | Yes |
| 3865941000033110 | Vancocin Matrigel 125mg capsules | Vancomycin hydrochloride | Glycopeptides | A07AA09 | Watch | Yes |
| 8945541000033110 | Vancomycin 250mg/5ml oral solution | Vancomycin hydrochloride | Glycopeptides | A07AA09 | Watch | Yes |
| 2800841000033110 | Vancomycin 500mg powder for solution for infusion vials | Vancomycin hydrochloride | Glycopeptides | A07AA09 | Watch | Yes |
| 1501341000033110 | Vancocin 1g powder for solution for infusion vials | Vancomycin hydrochloride | Glycopeptides | A07AA09 | Watch | Yes |
| 2800741000033110 | Vancocin 500mg powder for solution for infusion vials | Vancomycin hydrochloride | Glycopeptides | A07AA09 | Watch | Yes |
| 92641000033117 | Augmentin Intravenous 1.2g powder for solution for injection vials | Amoxicillin sodium/ Potassium clavulanate | Beta-lactam/beta-lactamase-inhibitor | J01CR02 | Access | Yes |
| 92741000033114 | Augmentin Intravenous 600mg powder for solution for injection vials | Amoxicillin sodium/ Potassium clavulanate | Beta-lactam/beta-lactamase-inhibitor | J01CR02 | Access | Yes |
| 330441000033114 | Co-amoxiclav 500mg/100mg powder for solution for injection vials | Amoxicillin sodium/ Potassium clavulanate | Beta-lactam/beta-lactamase-inhibitor | J01CR02 | Access | Yes |
| 330541000033110 | Co-amoxiclav 1000mg/200mg powder for solution for injection vials | Amoxicillin sodium/ Potassium clavulanate | Beta-lactam/beta-lactamase-inhibitor | J01CR02 | Access | Yes |
| 93141000033115 | Augmentin 375mg dispersible tablets | Amoxicillin trihydrate/ Potassium clavulanate | Beta-lactam/beta-lactamase-inhibitor | J01CR02 | Access | Yes |
| 94641000033110 | Augmentin 125/31 SF oral suspension | Amoxicillin trihydrate/ Potassium clavulanate | Beta-lactam/beta-lactamase-inhibitor | J01CR02 | Access | Yes |
| 94741000033118 | Augmentin 250/62 SF oral suspension | Amoxicillin trihydrate/ Potassium clavulanate | Beta-lactam/beta-lactamase-inhibitor | J01CR02 | Access | Yes |
| 94841000033111 | Augmentin-Duo 400/57 oral suspension | Amoxicillin trihydrate/ Potassium clavulanate | Beta-lactam/beta-lactamase-inhibitor | J01CR02 | Access | Yes |
| 94941000033115 | Augmentin 375mg tablets | Amoxicillin trihydrate/ Potassium clavulanate | Beta-lactam/beta-lactamase-inhibitor | J01CR02 | Access | Yes |
| 95041000033115 | Augmentin 625mg tablets | Amoxicillin trihydrate/ Potassium clavulanate | Beta-lactam/beta-lactamase-inhibitor | J01CR02 | Access | Yes |
| 317741000033113 | Co-amoxiclav 250mg/125mg dispersible tablets sugar free | Amoxicillin trihydrate/ Potassium clavulanate | Beta-lactam/beta-lactamase-inhibitor | J01CR02 | Access | Yes |
| 366941000033113 | Co-amoxiclav 125mg/31mg/5ml oral suspension sugar free | Amoxicillin trihydrate/ Potassium clavulanate | Beta-lactam/beta-lactamase-inhibitor | J01CR02 | Access | Yes |
| 367041000033114 | Co-amoxiclav 250mg/62mg/5ml oral suspension sugar free | Amoxicillin trihydrate/ Potassium clavulanate | Beta-lactam/beta-lactamase-inhibitor | J01CR02 | Access | Yes |
| 367541000033116 | Co-amoxiclav 400mg/57mg/5ml oral suspension sugar free | Amoxicillin trihydrate/ Potassium clavulanate | Beta-lactam/beta-lactamase-inhibitor | J01CR02 | Access | Yes |
| 370041000033115 | Co-amoxiclav 250mg/125mg tablets | Amoxicillin trihydrate/ Potassium clavulanate | Beta-lactam/beta-lactamase-inhibitor | J01CR02 | Access | Yes |
| 370141000033116 | Co-amoxiclav 500mg/125mg tablets | Amoxicillin trihydrate/ Potassium clavulanate | Beta-lactam/beta-lactamase-inhibitor | J01CR02 | Access | Yes |
| 1594141000033110 | Co-amoxiclav 875mg/125mg tablets | Amoxicillin trihydrate/ Potassium clavulanate | Beta-lactam/beta-lactamase-inhibitor | J01CR02 | Access | Yes |
| 2186941000033110 | Amiclav 250mg/125mg tablets | Amoxicillin trihydrate/ Potassium clavulanate | Beta-lactam/beta-lactamase-inhibitor | J01CR02 | Access | Yes |
| 2290741000033110 | Ranclav 375mg tablets | Amoxicillin trihydrate/ Potassium clavulanate | Beta-lactam/beta-lactamase-inhibitor | J01CR02 | Access | Yes |
| 2290941000033110 | Ranclav 125mg/31mg/5ml SF oral suspension | Amoxicillin trihydrate/ Potassium clavulanate | Beta-lactam/beta-lactamase-inhibitor | J01CR02 | Access | Yes |
| 5697041000033110 | Co-amoxiclav 125mg/31mg/5ml oral suspension | Amoxicillin trihydrate/ Potassium clavulanate | Beta-lactam/beta-lactamase-inhibitor | J01CR02 | Access | Yes |
| 5697141000033110 | Co-amoxiclav 250mg/62mg/5ml oral suspension | Amoxicillin trihydrate/ Potassium clavulanate | Beta-lactam/beta-lactamase-inhibitor | J01CR02 | Access | Yes |
| 329941000033113 | Co-fluampicil 250mg/250mg powder for solution for injection vials | Ampicillin sodium/ Flucloxacillin sodium | Combinations of penicillins | J01CR50 | Not-recommended | No |
| 860041000033112 | Magnapen 500mg powder for solution for injection vials | Ampicillin sodium/ Flucloxacillin sodium | Combinations of penicillins | J01CR50 | Not-recommended | No |
| 368341000033110 | Co-fluampicil 125mg/125mg/5ml oral suspension | Ampicillin trihydrate/ Flucloxacillin magnesium | Combinations of penicillins | J01CR50 | Not-recommended | No |
| 867741000033114 | Magnapen syrup | Ampicillin trihydrate/ Flucloxacillin magnesium | Combinations of penicillins | J01CR50 | Not-recommended | No |
| 294741000033111 | Co-fluampicil 250mg/250mg capsules | Ampicillin trihydrate/ Flucloxacillin sodium | Combinations of penicillins | J01CR50 | Not-recommended | No |
| 857741000033118 | Magnapen 250mg/250mg capsules | Ampicillin trihydrate/ Flucloxacillin sodium | Combinations of penicillins | J01CR50 | Not-recommended | No |
| 10740641000033100 | Ceftolozane 1g / Tazobactam 500mg powder for solution for infusion vials | Ceftolozane sulfate/ Tazobactam sodium | Fifth-generation cephalosporins | J01DI54 | Reserve | No |
| 10740741000033100 | Zerbaxa 1g/0.5g powder for concentrate for solution for infusion vials | Ceftolozane sulfate/ Tazobactam sodium | Fifth-generation cephalosporins | J01DI54 | Reserve | No |
| 429741000033115 | Deteclo 300mg tablets | Chlortetracycline hydrochloride/ Demeclocycline hydrochloride/ Tetracycline hydrochloride | Combinations of tetracyclines | J01AA20 | Watch | No |
| 753141000033112 | Imipenem 500mg / Cilastatin 500mg powder for solution for infusion vials | Cilastatin sodium/ Imipenem monohydrate | Carbapenems | J01DH51 | Watch | No |
| 1120741000033110 | Primaxin IV 500mg powder for solution for infusion vials | Cilastatin sodium/ Imipenem monohydrate | Carbapenems | J01DH51 | Watch | No |
| 911041000033113 | Mictral granules 7g sachets | Citric acid/ Nalidixic acid/ Sodium bicarbonate/ Sodium citrate | Quinolones | J01MB02 | Watch | No |
| 1405141000033110 | Tazocin 2g/0.25g powder for solution for infusion vials | Piperacillin sodium/ Tazobactam sodium | Beta-lactam/beta-lactamase-inhibitor_anti-pseudomonal | J01CR05 | Watch | Yes |
| 1405241000033110 | Tazocin 4g/0.5g powder for solution for infusion vials | Piperacillin sodium/ Tazobactam sodium | Beta-lactam/beta-lactamase-inhibitor_anti-pseudomonal | J01CR05 | Watch | Yes |
| 5006741000033110 | Piperacillin 2g / Tazobactam 250mg powder for solution for infusion vials | Piperacillin sodium/ Tazobactam sodium | Beta-lactam/beta-lactamase-inhibitor_anti-pseudomonal | J01CR05 | Watch | Yes |
| 5006841000033110 | Piperacillin 4g / Tazobactam 500mg powder for solution for infusion vials | Piperacillin sodium/ Tazobactam sodium | Beta-lactam/beta-lactamase-inhibitor_anti-pseudomonal | J01CR05 | Watch | Yes |
| 1441341000033110 | Timentin 3.2g powder for solution for infusion vials | Potassium clavulanate/ Ticarcillin sodium | Beta-lactam/beta-lactamase-inhibitor | J01CR03 | Unclassified | No |
| 333441000033116 | Co-trimoxazole 80mg/400mg/5ml solution for infusion ampoules | Sulfamethoxazole/ Trimethoprim | Sulfonamide-trimethoprim-combinations | J01EE01 | Access | Yes |
| 1271441000033110 | Septrin for Infusion 80mg/400mg/5ml solution for infusion ampoules | Sulfamethoxazole/ Trimethoprim | Sulfonamide-trimethoprim-combinations | J01EE02 | Access | Yes |
| 289441000033119 | Co-trimoxazole 80mg/400mg/5ml oral suspension | Sulfamethoxazole/ Trimethoprim | Sulfonamide-trimethoprim-combinations | J01EE03 | Access | Yes |
| 366841000033117 | Co-trimoxazole 40mg/200mg/5ml oral suspension sugar free | Sulfamethoxazole/ Trimethoprim | Sulfonamide-trimethoprim-combinations | J01EE04 | Access | Yes |
| 373441000033119 | Co-trimoxazole 80mg/400mg tablets | Sulfamethoxazole/ Trimethoprim | Sulfonamide-trimethoprim-combinations | J01EE05 | Access | Yes |
| 373541000033118 | Co-trimoxazole 160mg/800mg tablets | Sulfamethoxazole/ Trimethoprim | Sulfonamide-trimethoprim-combinations | J01EE06 | Access | Yes |
| 1267541000033110 | Septrin Adult 80mg/400mg/5ml oral suspension | Sulfamethoxazole/ Trimethoprim | Sulfonamide-trimethoprim-combinations | J01EE07 | Access | Yes |
| 1272641000033110 | Septrin Paediatric 40mg/200mg/5ml oral suspension | Sulfamethoxazole/ Trimethoprim | Sulfonamide-trimethoprim-combinations | J01EE08 | Access | Yes |
| 1275641000033110 | Septrin tablets | Sulfamethoxazole/ Trimethoprim | Sulfonamide-trimethoprim-combinations | J01EE09 | Access | Yes |
| 1275741000033110 | Septrin Forte 160mg/800mg tablets | Sulfamethoxazole/ Trimethoprim | Sulfonamide-trimethoprim-combinations | J01EE10 | Access | Yes |

**Appendix 4.**

Steps for estimating how total variance in antibiotic prescribing could be attributed to different variance components

---------------------------------------------------------------------------------

1. **Estimating patient case-mix variance**

Patient case-mix variance was estimated using the following steps.

First, a fixed-effects logistic regression model was fitted with antibiotic prescribing decision (yes/no) as the binary outcome, including GP practice identifier and all patient-level case-mix covariates as predictors. This model was used to generate, for each patient, predicted probabilities of antibiotic prescribing based on their covariates and the GP practice attended. Patient case-mix covariates included sociodemographic factors (age, sex, ethnicity, and deprivation status^1^), smoking status, comorbidities (Elixhauser comorbidity score^2^, diabetes, chronic kidney disease, and chronic lung disease), previous use of immunosuppressants within the past 6 months, history of hospitalisation and/or outpatient visits in the preceding year, and vaccination status (influenza and pneumococcal vaccines). We selected case-mix covariates for each model based on clinical relevance; for example, chronic lung disease and influenza and pneumococcal vaccination were included only in models for respiratory infections and not for UTIs or impetigo. Associations between these covariates and the antibiotic prescribing decision are shown in *Appendix 5*.

Second, a multinomial regression was fitted with the practice identifier as the outcome and the same set of patient-level covariates as predictors. This model was subsequently used to generate, for each patient, the predicted probability of attending each GP practice given their characteristics. Let *Z* denote the GP practice attended, taking values $z\mathcal{\in Z} = \{1,...,m\} ,$and let *X_i_* denote the vector of patient-level covariates for patient *i*. For each patient *i*, the predicted prescribing probabilities across all practices, $E(Y_{i}(z)|X_{i})$ were multiplied by the corresponding predicted attendance probabilities, $P{(Z}_{i}=z | X_{i})$ and summed over all practices to obtain the patient’s expected probability of antibiotic prescribing:

$$\overline{Y}_{i}= \sum_{z\in\mathcal{Z}} E{(Y}_{i}(z) | X_{i}) P(Z_{i}=z | X_{i})$$

The quantity $\overline{Y}_{i}$ represents the expected probability of antibiotic prescribing for patient *i,* averaged over all GP practices according to the probability that a patient with similar characteristics would attend each practice. It therefore captures prescribing probability attributable solely to patient case-mix.

The patient case-mix variance component was then defined as the variance of these expected probabilities across patients, $V_{case-mix}=Var_{i}\left( \overline{Y}_{i} \right).$ This represents the portion of total variance in antibiotic prescribing explained by differences in patient characteristics rather than by the practice themselves.

2. **Estimating between-practice variance**

Between-practice variance was estimated as the average, across all patients, of the variability in predicted prescribing probabilities between GP practices after accounting for patient case-mix. For each patient *i*, the predicted prescribing probabilities across all practices, $EY_{\left( Z=z \right)|x_{i}}$ , were combined with the corresponding attendance probabilities, $P(Z=z|X_{i})$, to calculate how much the predicted probability of receiving an antibiotic would vary if that same patient were seen by different practices. For each patient, the variability across practices was calculated as:

$Var_{Z|X_{i}}\left( E{(Y}_{i}(z) |X_{i} \right))= \sum_{z \in\mathcal{Z}} E\left( Y_{i}\left( z \right) \right|X_{i})^{2} P\left( Z_{i}=z | X_{i} \right)- \bar{Y}_{i}^{2}$

The between-practice variance component was then defined as

$$V_{practice}= \frac{1}{N}\sum_{i=1}^{N} Var_{Z|X_{i}}(E(Y_{i}\left( z \right)| X_{i}))$$

This component reflects the proportion of total variance in antibiotic prescribing that remains attributable to systematic differences between GP practices, after adjusting for case-mix.

3. **Estimating residual unexplained variance**

Residual variance was then obtained by subtracting the patient and GP-level components from the empirical total variance in antibiotic prescribing.

**Appendix 5.**

Patient case-mix factors included in the decomposition analysis and their associations with antibiotic prescribing decisions for each condition

---------------------------------------------------------------------------------

We used mixed-effects logistic regression models to estimate odds ratios (ORs) and 95% confidence intervals (CIs). Fixed effects included sociodemographic factors (age, sex, ethnicity, and deprivation status^1^), smoking status, comorbidities (Elixhauser comorbidity score^2^, diabetes, chronic kidney disease, and chronic lung disease), previous use of medications (immunosuppressants and antibiotics), history of hospitalisation and/or outpatient visits in the preceding year, and vaccination status (influenza and pneumococcal vaccines). We selected fixed effects for each model based on clinical relevance; for example, chronic lung disease and influenza and pneumococcal vaccination were included only in models for respiratory infections and not for UTIs, or impetigo. GP identifier was included as a random effect to account for clustering within practices.

The results show that several patient-level factors were associated with the odds of receiving an antibiotic in a multivariable analysis including all covariates of interest. Compared with patients aged 25 to 39 years, those younger than 25 years (including the ≤5, 6–9, 10–17, and 18–24 age groups) had lower odds of receiving antibiotics, while those aged 40 to 74 years (including the 40–59 and 60–74 groups) had higher odds across most infections (except for sore throat and impetigo). Female patients had higher odds than males in seven infections: URTI, acute cough, rhinosinusitis, sore throat, LRTI and UTI. Patients who had ever smoked had higher odds than non-smokers across 8 of 11 infections. Those who had received antibiotics within the previous six months also had higher odds compared with those who had not, across all infections; a similar pattern was observed for patients who had received immunosuppressants (across 10 of 11 infections). Receiving an influenza vaccination within the past year and a pneumococcal vaccination within the past ten years was also associated with higher odds of prescribing in most respiratory tract infections.

Some of these findings should be interpreted with caution. For example, consistent with Palin et al.,^3^ we found that patients who had received vaccines had a higher risk of antibiotic prescribing than those who had not. Although these associations persisted after adjustment for other covariates, they were observed among patients who consulted and should not be interpreted as causal effects of vaccination on antibiotic use, a pattern likely explained by collider bias. Conditioning on consultation can induce associations between patient characteristics and prescribing that do not exist in the general population at risk of infection. For example, vaccinated individuals may be less likely to consult when experiencing mild illness, meaning that among consulters, vaccinated patients represent a group with more severe or persistent symptoms and therefore a higher likelihood of receiving antibiotics.

However, collider bias is less relevant for the variance decomposition, which by design examines variation in antibiotic prescribing among patients who consulted. The analysis is conditional on consultation and does not estimate the overall risk of receiving an antibiotic after infection, many of which never lead to a GP visit.

**UPPER RESPIRATORY TRACT INFECTION**

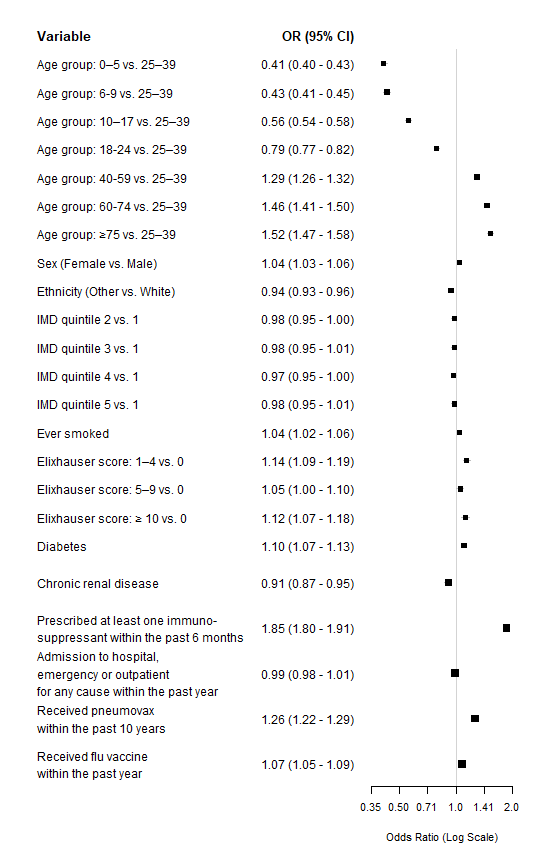

**ACUTE COUGH**

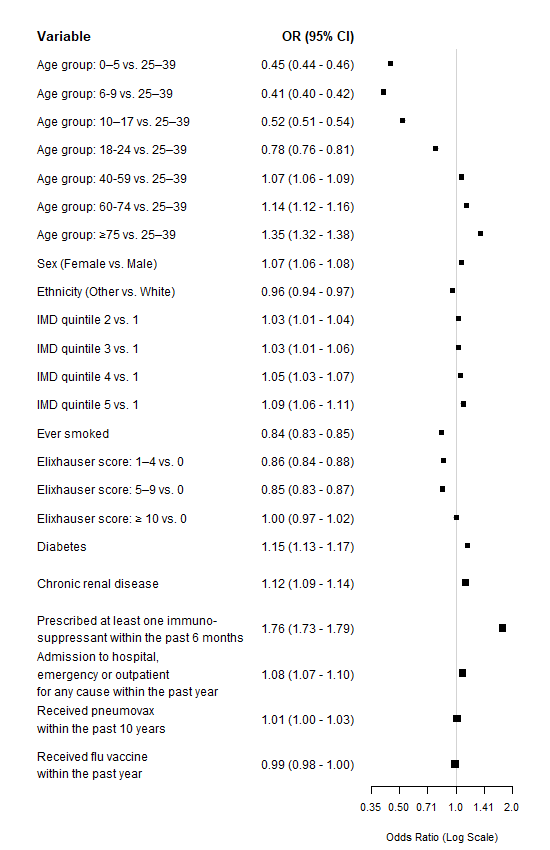

**ACUTE RHINOSINUSITIS**

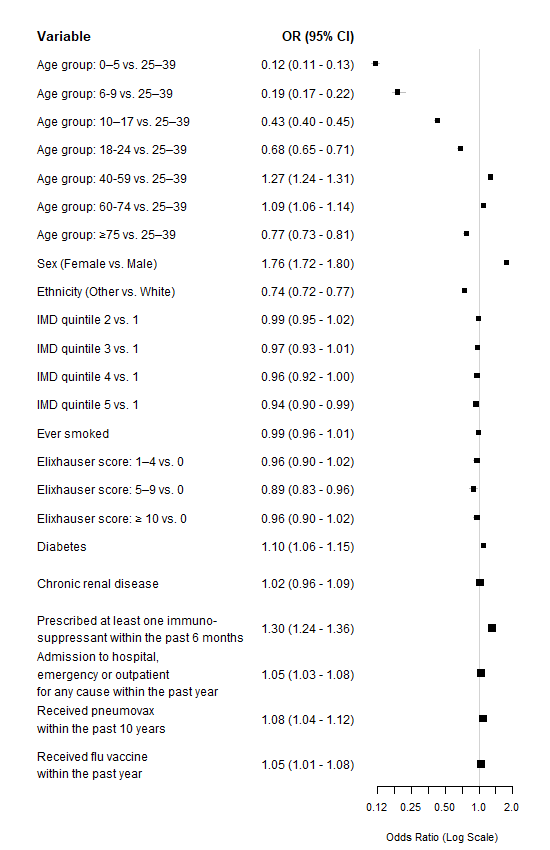

**ACUTE SORE THROAT**

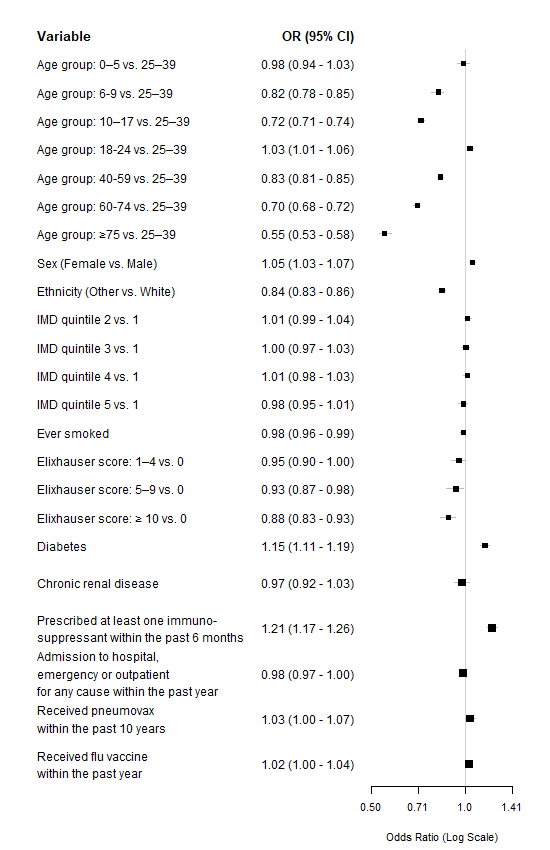

**LOWER RESPIRATORY INFECTION**

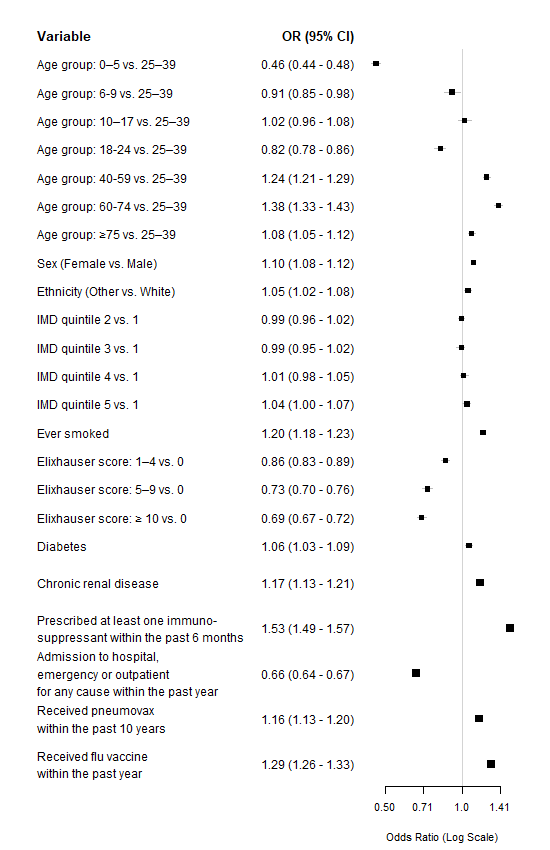

**ACUTE BRONCHITIS**

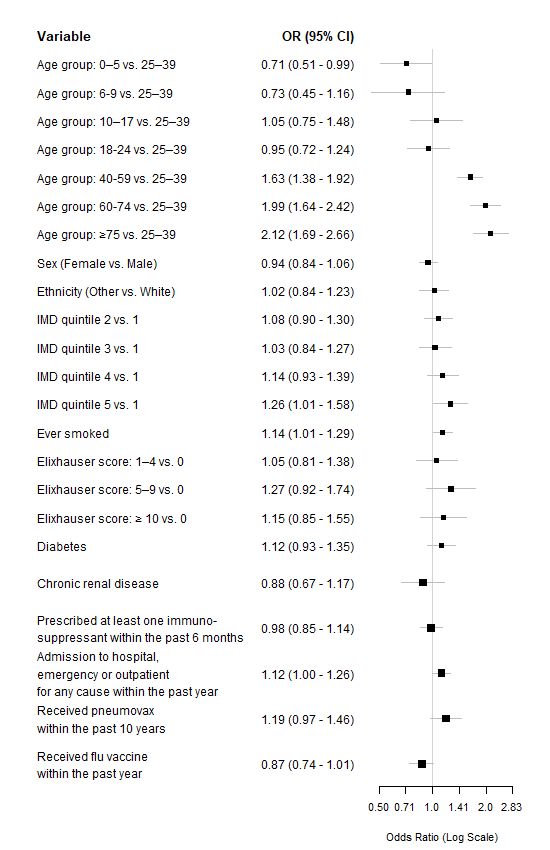

**ASTHMA EXACERBATION**

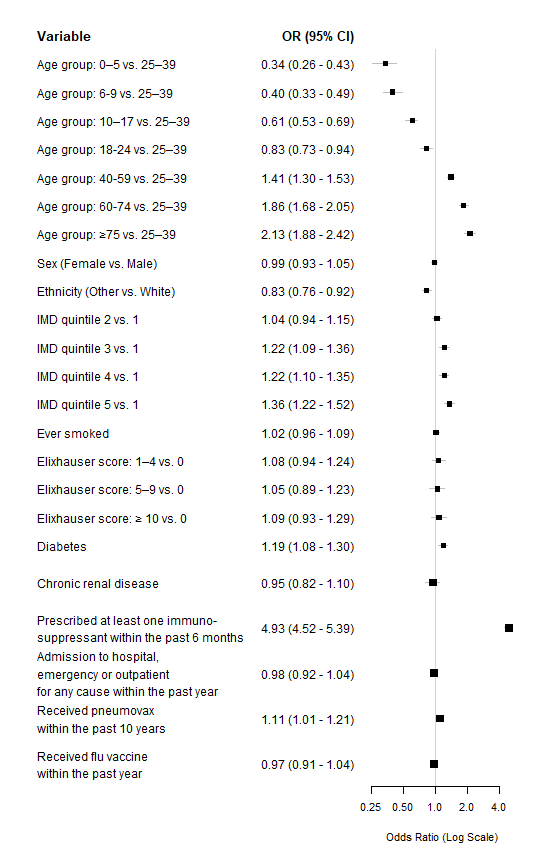

**COPD EXACERBATION**

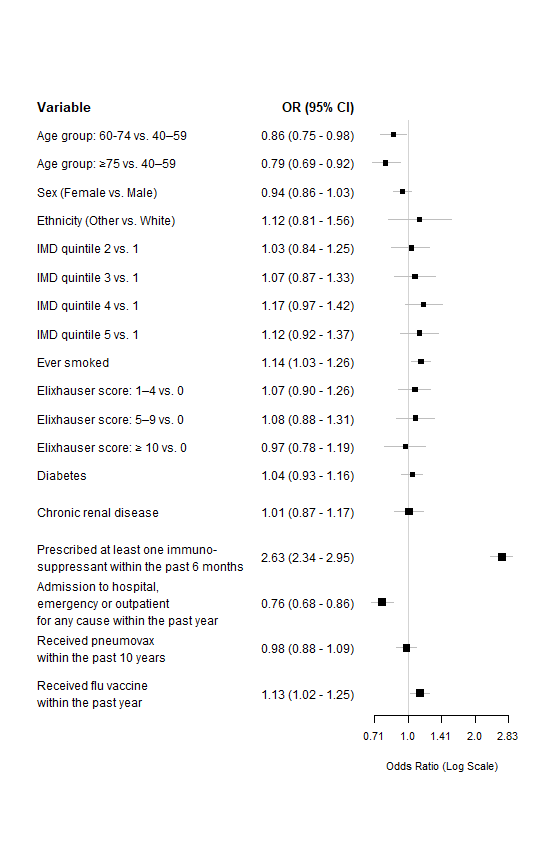

**LOWER URINARY TRACT INFECTION**

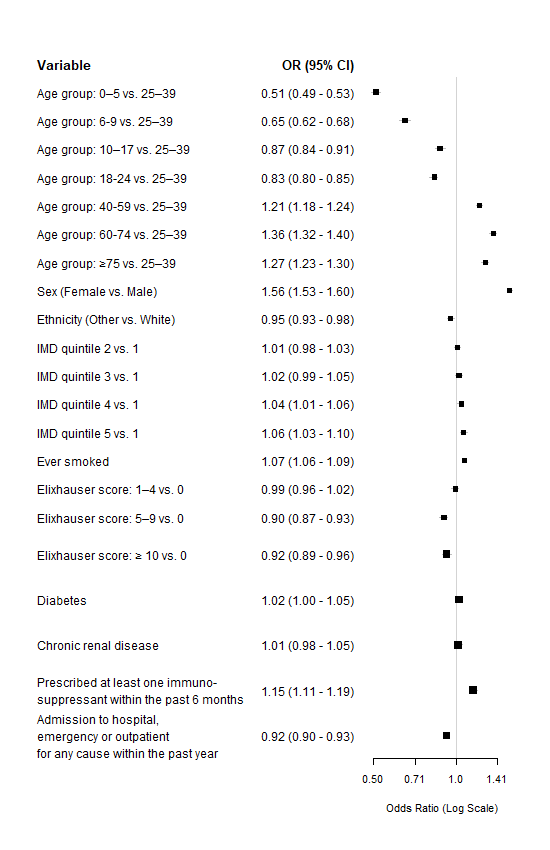

**IMPETIGO**

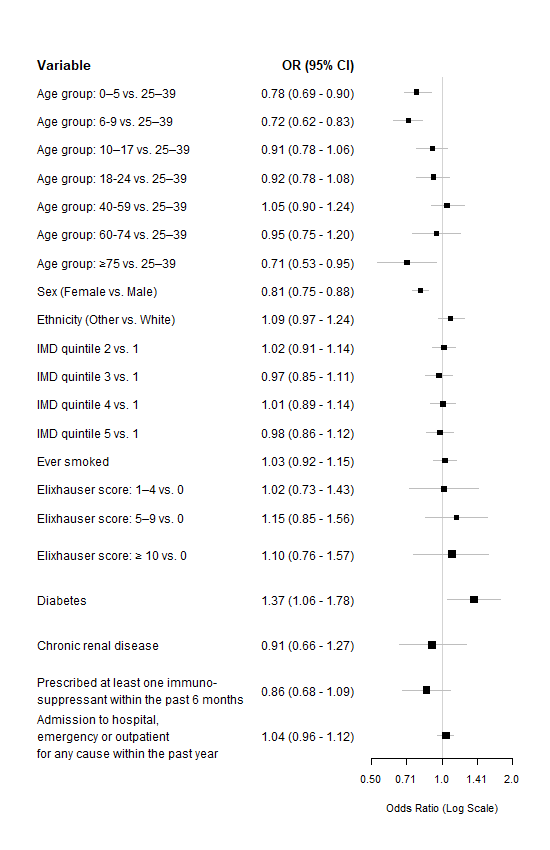

**Appendix 6**

Distribution of infection severity conditional on GP practice prescribing quartile and prescribing decision in patient aged ≥16 years with acute sore throat and lower respiratory tract infection^4^

---------------------------------------------------------------------------------

**Table.** Distribution of GP consultations for acute sore throat by FeverPAIN score across GP practices grouped by quartile of antibiotic prescribing proportion

| **Antibiotic prescribing proportion quartile** | **FeverPAIN 0/1 (%)** | **FeverPAIN 2/3 (%)** | **FeverPAIN 4/5 (%)** |
| --- | --- | --- | --- |
| 1 (lowest) | 58.2 | 35.7 | 6.1 |
| 2 | 55.6 | 36.2 | 8.2 |
| 3 | 51.5 | 39.4 | 9.1 |
| 4(highest) | 39.7 | 51 | 9.3 |

**Table.** Distribution of GP consultations for acute sore throat by FeverPAIN score, stratified by antibiotic prescribing status across GP practices

| **Antibiotic prescribing status** | **FeverPAIN 0/1 (%)** | **FeverPAIN 2/3 (%)** | **FeverPAIN 4/5 (%)** |
| --- | --- | --- | --- |
| Received antibiotic | 35.0 | 49.4 | 15.6 |
| Not received antibiotic | 71.2 | 28.2 | 0.6 |

**Table.** Distribution of GP consultations for LRTI by Pneumonia risk score score across GP practices grouped by quartile of antibiotic prescribing proportion

| **Antibiotic prescribing proportion quartile** | **No signs for pneumonia (%)** | **1+ signs for pneumonia**  **(%)** |
| --- | --- | --- |
| 1 (lowest) | 63 | 37 |
| 2 | 54 | 46 |
| 3 | 45.3 | 54.7 |
| 4(highest) | 30.9 | 60.1 |

**Table.** Distribution of GP consultations for acute sore throat by FeverPAIN score, stratified by antibiotic prescribing status across GP practices

| **Antibiotic prescribing status** | **No signs for pneumonia (%)** | **1+ signs for pneumonia**  **(%)** |
| --- | --- | --- |
| Received antibiotic | 27.3 | 72.7 |
| Not received antibiotic | 84.3 | 15.7 |

**Appendix 7.** Detailed results of the three-way variance decomposition analysis showing how different sources of variance contributed to the overall variance in antibiotic prescribing for each infection, and how the contributions of the explained variance components (between-practice variance and patient case-mix) changed across three models with increasing levels of case-mix adjustment.

|  | **Model 1 (full case-mix adjustment)** | | | **Model 2 (age-sex adjustment)** | | | **Model 3 (no adjustment)** | |
| --- | --- | --- | --- | --- | --- | --- | --- | --- |
|  | Source of variance | | | Source of variance | | | Source of variance | |
|  | Patient case-mix | Between-practice | Residual unexplained | Patient case-mix | Between-practice | Residual unexplained | Between-practice | Residual unexplained |
| **Upper respiratory tract infection** |  |  |  |  |  |  |  |  |
| Variance | 0.006 (0.006 – 0.006) | 0.015 (0.014 – 0.015) | 0.122 (0.121 – 0.122) | <0.001 (<0.001 – <0.001) | 0.015 (0.015 – 0.016) | 0.127 (0.126 – 0.128) | 0.015 (0.015 – 0.016) | 0.127 (0.126 – 0.128) |
| Proportion of total variance attributable to each component (%) | 4.1 (3.9 – 4.3) | 10.3 (10 – 10.9) | 85.6 (84.9 – 85.9) | <0.1 (<0.1 – <0.1) | 10.6 (10.3 – 11.2) | 89.4 (88.8 – 89.7) | 10.6 (10.4 – 11.2) | 89.4 (88.8 – 89.6) |
| **Acute cough** |  |  |  |  |  |  |  |  |
| Variance | 0.008 (0.008 – 0.008) | 0.014 (0.014 – 0.015) | 0.201 (0.2 – 0.202) | <0.001  (<0.001 – 0.001) | 0.014 (0.014 – 0.015) | 0.208 (0.207 – 0.208) | 0.014 (0.014 – 0.015) | 0.209 (0.208 – 0.209) |
| Proportion of total variance attributable to each component (%) | 3.6 (3.4 – 3.7) | 6.2 (6.1 – 6.6) | 90.2 (89.8 – 90.4) | 0.4 (0.4 – 0.5) | 6.3 (6.3 – 6.7) | 93.3 (92.9 – 93.3) | 6.3 (6.2 – 6.6) | 93.7 (93.4 – 93.8) |
| **Acute rhino sinusitis** |  |  |  |  |  |  |  |  |
| Variance | 0.02 (0.019 – 0.021) | 0.017 (0.017 – 0.019) | 0.213 (0.21 – 0.213) | <0.001 (<0.001 – 0.001) | 0.02 (0.02 – 0.023) | 0.229 (0.226 – 0.228) | 0.02 (0.02 – 0.023) | 0.23 (0.227 – 0.23) |
| Proportion of total variance attributable to each component (%) | 8.1 (7.6 – 8.5) | 6.8 (6.9 – 7.8) | 85.2 (84 – 85.3) | 0.4 (0.3 – 0.5) | 8.1 (8.1 – 9.2) | 91.5 (90.4 – 91.4) | 8.1 (8.1 – 9.2) | 91.9 (90.8 – 91.9) |
| **Acute sore throat** |  |  |  |  |  |  |  |  |
| Variance | 0.002 (0.002 – 0.002) | 0.015 (0.015 – 0.017) | 0.223 (0.221 – 0.223) | <0.001 (<0.001 – <0.001) | 0.015 (0.015 – 0.017) | 0.225 (0.223 – 0.225) | 0.015 (0.015 – 0.016) | 0.225 (0.223 – 0.224) |
| Proportion of total variance attributable to each component (%) | 0.8 (0.7 – 0.9) | 6.1 (6.1 – 6.9) | 93.1 (92.2 – 93.1) | <0.1 (<0.1 – <0.1) | 6.1 (6.1 – 7) | 93.9 (93 – 93.9) | 6.1 (6.2 – 6.9) | 93.9 (93.1 – 93.8) |
| **Lower respiratory tract infection** |  |  |  |  |  |  |  |  |
| Variance | 0.005 (0.004 – 0.005) | 0.015 (0.015 – 0.016) | 0.125 (0.124 – 0.125) | <0.001 (<0.001 – <0.001) | 0.016 (0.015 – 0.017) | 0.129 (0.128 – 0.129) | 0.015 (0.015 – 0.016) | 0.129 (0.128 – 0.129) |
| Proportion of total variance attributable to each component (%) | 3.1 (2.9 – 3.4) | 10.5 (10.5 – 11.2) | 86.4 (85.6 – 86.5) | <0.1 (<0.1 – <0.1) | 10.8 (10.7 – 11.5) | 89.2 (88.5 – 89.3) | 10.7 (10.6 – 11.3) | 89.3 (88.7 – 89.4) |
| **Acute bronchitis** |  |  |  |  |  |  |  |  |
| Variance | 0.017 (0.013 – 0.021) | 0.074 (0.042 – 0.078) | 0.136 (0.131 – 0.169) | <0.001 (<0.001 – <0.001) | 0.086 (0.046 – 0.091) | 0.14 (0.136 – 0.181) | 0.089 (0.046 – 0.094) | 0.137 (0.132 – 0.181) |
| Proportion of total variance attributable to each component (%) | 7.3 (5.7 – 9.2) | 32.6 (18.4 – 34.4) | 60.1 (57.6 – 74.5) | <0.1 (<0.1 – 0.4) | 38 (20.1 – 39.9) | 61.9 (60 – 79.8) | 39.3 (20.2 – 41.5) | 60.7 (58.5 – 79.8) |
| **Acute otitis media** |  |  |  |  |  |  |  |  |
| Variance | 0.002 (0.001 – 0.002) | 0.024 (0.023 – 0.026) | 0.161 (0.158 – 0.162) | 0.001 (<0.001 – 0.001) | 0.024 (0.023 – 0.027) | 0.162 (0.159 – 0.162) | 0.024 (0.024 – 0.027) | 0.162 (0.159 – 0.162) |
| Proportion of total variance attributable to each component (%) | 1 (0.7 – 1.3) | 12.6 (12.3 – 14.2) | 86.4 (84.7 – 86.7) | 0.5 (0.4 – 0.8) | 12.8 (12.3 – 14.4) | 86.7 (85 – 87.1) | 13 (12.7 – 14.4) | 87 (85.6 – 87.3) |
| **Asthma exacerbation** |  |  |  |  |  |  |  |  |
| Variance | 0.029 (0.026 – 0.032) | 0.022 (0.021 – 0.028) | 0.199 (0.191 – 0.202) | 0.006 (0.004 – 0.008) | 0.025 (0.024 – 0.03) | 0.219 (0.213 – 0.22) | 0.024 (0.021 – 0.031) | 0.226 (0.219 – 0.229) |
| Proportion of total variance attributable to each component (%) | 11.4 (10.3 – 12.8) | 9 (8.2 – 11.3) | 79.6 (76.6 – 80.7) | 2.3 (1.5 – 3.4) | 10 (9.6 – 12.2) | 87.7 (85.4 – 88.2) | 9.6 (8.4 – 12.3) | 90.4 (87.7 – 91.6) |
| **COPD exacerbation** |  |  |  |  |  |  |  |  |
| Variance | 0.006 (0.005 – 0.01) | 0.011 (0.01 – 0.018) | 0.141 (0.132 – 0.142) | <0.001 (<0.001 – 0.002) | 0.011 (0.009 – 0.017) | 0.147 (0.141 – 0.15) | 0.012 (0.009 – 0.018) | 0.147 (0.14 – 0.149) |
| Proportion of total variance attributable to each component (%) | 3.5 (3.1 – 6.5) | 7.2 (6.3 – 11.2) | 89.3 (83.4 – 90) | 0.2 (<0.1 – 1.1) | 7 (5.4 – 10.9) | 92.7 (88.9 – 94.5) | 7.4 (5.8 – 11.4) | 92.6 (88.6 – 94.2) |
| **Lower urinary tract infection** |  |  |  |  |  |  |  |  |
| Variance | 0.002 (0.002 – 0.002) | 0.015 (0.015 – 0.016) | 0.133 (0.132 – 0.133) | <0.001 (<0.001 – <0.001) | 0.016 (0.015 – 0.017) | 0.129 (0.128 – 0.129) | 0.015 (0.015 – 0.016) | 0.129 (0.128 – 0.129) |
| Proportion of total variance attributable to each component (%) | 1.4 (1.3 – 1.5) | 10 (9.9 – 10.4) | 88.6 (88.1 – 88.7) | <0.1 (<0.1 – <0.1) | 10.8 (10.7 – 11.5) | 89.2 (88.5 – 89.3) | 10.7 (10.6 – 11.3) | 89.3 (88.7 – 89.4) |
| **Impetigo** |  |  |  |  |  |  |  |  |
| Variance | 0.002 (0.002 – 0.005) | 0.014 (0.013 – 0.02) | 0.227 (0.219 – 0.227) | <0.001 (<0.001 – <0.001) | 0.014 (0.013 – 0.021) | 0.229 (0.221 – 0.229) | 0.013 (0.011 – 0.021) | 0.231 (0.223 – 0.232) |
| Proportion of total variance attributable to each component (%) | 0.8 (0.7 – 2.2) | 5.8 (5.3 – 8.3) | 93.4 (90.3 – 93.4) | <0.1 (<0.1 – 0.4) | 5.7 (5.4 – 8.5) | 94.2 (91.2 – 94.5) | 5.4 (4.7 – 8.5) | 94.6 (91.5 – 95.3) |

For each condition, results are presented as variance estimates for each component (patient case-mix, between-practice, and residual unexplained variation), together with the corresponding proportion of total variance attributable to each component. All estimates are reported as point estimates with 95% confidence intervals using 100 posterior simulations based on the estimated coefficients and variance-covariance matrices. ^a^: patient case-mix factors included sociodemographic factors (age, sex, ethnicity, and deprivation status), smoking status, comorbidities (Elixhauser comorbidity score, diabetes, chronic kidney disease, and chronic lung disease), use of immunosuppressants within the previous 6 months, history of hospitalisation and/or outpatient visits in the preceding year, vaccination status (influenza and pneumococcal), and practice identifiers. Inclusion of case-mix covariates for each condition was based on clinical relevance; for example, chronic lung disease and influenza and pneumococcal vaccination were included only in models for respiratory infections and not for UTIs, or impetigo.

**Model 1** (full case-mix adjustment) included sociodemographic factors (age, sex, ethnicity, and deprivation status), smoking status, comorbidities (Elixhauser comorbidity score, diabetes, chronic kidney disease, and chronic lung disease), use of immunosuppressants within the previous 6 months, history of hospitalisation and/or outpatient visits in the preceding year, vaccination status (influenza and pneumococcal), and practice identifiers. Inclusion of case-mix covariates for each condition was based on clinical relevance. **Model 2** (minimal case-mix adjustment) included only patient age and sex as covariates, consistent with the current STAR-PU methodology, and practice identifiers. **Model 3** (no case-mix adjustment) included practice identifiers only, with no case-mix adjustment. Panel **A** shows changes in contributions to total variance (the sum of patient case-mix variance, between-practice variance, and residual unexplained variance

REFERENCE

1.	Ministry of Housing Communities and Local Government. The English indices of deprivation 2019: Statistical release. London: MHCLG; 2019.

2.	van Walraven C, Austin PC, Jennings A, Quan H, Forster AJ. A modification of the Elixhauser comorbidity measures into a point system for hospital death using administrative data. *Med Care* 2009; **47**(6): 626-33.

3.	Palin V, Mölter A, Belmonte M, et al. Antibiotic prescribing for common infections in UK general practice: variability and drivers. *J Antimicrob Chemother* 2019; **74**(8): 2440-50.

4.	Stuart B, Brotherwood H, Van't Hoff C, et al. Exploring the appropriateness of antibiotic prescribing for common respiratory tract infections in UK primary care. *J Antimicrob Chemother* 2020; **75**(1): 236-42.
